# Counting Cells by Age Tells Us About How, and Why, and When, We Grow, and Become Old and Ill

**DOI:** 10.1101/2023.01.05.23284244

**Authors:** Luca Citi, Jessica Su, Luke Huang, James S Michaelson

**Affiliations:** School of Computer Science and Electronic Engineering, University of Essex, UK; Department of Biomedical Engineering, Johns Hopkins University, USA; Department of Pathology, Massachusetts General Hospital, USA; Department of Surgery, Massachusetts General Hospital, USA; Department of Pathology, Harvard Medical School, USA

## Abstract

Growth and aging are fundamental features of animal life. The march from fertilization to oblivion comes in enormous variety: days and hundreds of cells for nematodes, decades and trillions of cells for humans.^1-4^ Since Verhulst (1838^5^) proposed the Logistic Equation - exponential growth with a countervailing linear decline in rate – biologists have searched for ever better density-dependent growth equations,^6-12^ none of which accurately capture the relationship between size and time for real animals.^13-15^ Furthermore, while growth and aging run in parallel, whether the relationship is causal has yet to be determined. Similarly unknown has been the reason behind the exponential *Force of Mortality*, described by Gompertz in 1825 for all-cause mortality^16^ and reported by Levin et al. in 2020 for COVID-19.^17^ Here we report that examination in units of numbers of cells, *N, Cellular Phylodynamic Analysis*,^6^ reveals that growth, lifespan, and mortality, are linked to the reduction in the fraction of cells dividing, occurring by a simple expression, the *Universal Mitotic Fraction Equation*. Lifespan is correlated with an age when fewer than one-in-a-thousand cells are dividing, quantifying the long-appreciated mechanism of aging, the failure of cells to be rejuvenated by dilution with new materials made and DNA repaired at mitosis.^29-31^ These observations provide practical mathematical tools for comprehending and managing the challenges of growth and aging, for such tasks as deciphering COVID-19 lethality and its amelioration by vaccination.

## INTRODUCTION

### Counting Cells: *Cellular Phylodynamic Analysis*

As we grow older, we grow bigger, and we grow slower, and we grow frailer. Insight into the underlying processes at work can be gained by considering growth in units of numbers of cells, ***N***. We have called this approach ***Cellular Phylodynamic Analysis***,^6^ building from Stadler, Pybus, and Stumpf,^18^ who have shown how methods developed for studying the populations of organisms in nature can also be used in studying the populations of cells within us.^19^ Taking this perspective has provided a understanding of the growth and creation of the body and its parts,^6,20^ for quantifying the transformation, by somatic mutation, of normal growth into precancerous growth and then into cancer,^21^ for understanding how cancer cells spread,^21,22,23^ and for putting this knowledge to practical use.^24,25^ Here, we will explore how such a ***Cellular Phylodynamic Analysis*** can give us insight into growth and its link to aging.

### The Force of Mortality

Different species live to different lifespans - worms to 2 weeks, mice to 2 years, humans to a century – and different creatures in each species live to different ages. In 1835, Gompertz^16^ reported that the chance of death for humans, ***D***, doubles with each 5–6-year increase in age, ***t***, and in 2020 Levin et al^17^ reported the same relationship for COVID-19 lethality (FIGURE 1). This exponential increase in the chance of death, the ***Gompertzian Force of Mortality***, can be expressed mathematically as the ***Gompertz Mortality Equation***, captured by Equations #7 and #8 below, or graphically, by comparing the chance of death, ***D***, against age, ***t***, on log graphs, ***Gompertz Plots***, thus yielding straight lines, ***Gompertz Lines***, of slope, ***G***_***s***_ (***Gompertzian Slope***) and height ***G***_***H***_ (***Gompertzian Height***) (FIGURE 1).

**FIGURE 1:**
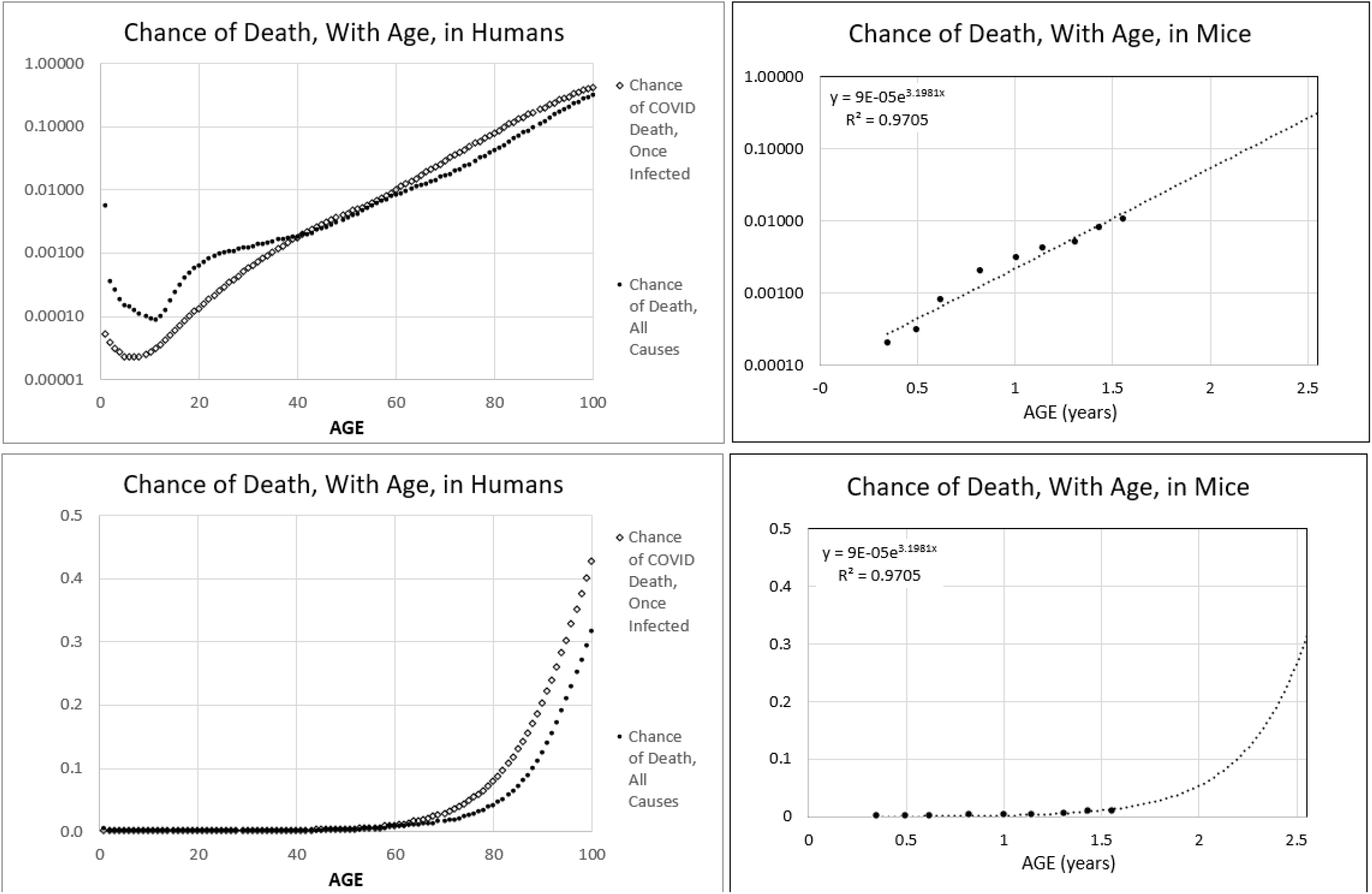

The ***Gompertzian Force of Mortality*** is such a familiar presence in our lives that we have trouble seeing it in plain view. Its destructive reality can be appreciated by considering what life would be like without it. The chance of death at age 60 is about 1%/year. If a thousand 60-year-olds continued with that 1% yearly chance of death -- that is, if there was no exponential ***Gompertzian Force of Mortality*** -- by age 100, more than six hundred would still be alive; by age 200 more than two hundred would survive; by age 300 more than a hundred would remain, and the last would die around age 750. Aging’s ***Gompertzian Force of Mortality*** is the most catastrophic, life-stealing disease we pay no attention to.

### COVID’s Force of Mortality

***Gompertz Plots*** for all-cause mortality in mice, all-cause mortality in humans, and COVID-19 mortality in humans at the beginning of the pandemic, can be seen in FIGURE 1. Graphs on the top row of this figure show the log-linear ***Gompertz Lines*** of the exponential ***Gompertz Mortality Equation***. When linearized, these data display the shocking, progressively more lethal outcome of COVID-19 that occurs as we age. Such a linear graph can be seen at the bottom of FIGURE 1, where we can hardly detect COVID-19 lethality (Deaths/Cases) in the first five decades of life when its ***Gompertzian Force of Mortality*** stealthily accumulates less than 1% chance of death. However, from age 60 on, we see the progressively more catastrophic, exponential, increase in COVID-19 lethality, with risk of death rising from about 1% at age 60, to about 3% at age 70, to almost 10% at age 80, to almost 20% at age 90, and to almost 50% at age 100. It is not COVID-19 that has caused all the death and suffering we have experienced in the last two years. It is growing old. We have seen this in the trivial lethality evident among those younger than 60 and the heartbreaking tragedy on display in our old-age homes.

### Counting Cells: Counting Lives

Here, we shall see that ***Cellular Phylodynamic Analysis***, counting cells, reveals that growth across the animal kingdom is driven by the reduction in the fraction of cells dividing, as captured by a simple expression, the ***Universal Mitotic Fraction Equation. Cellular Phylodynamic Analysis*** further reveals that growth of the body occurs by another expression, the ***Universal Growth Equation***, and growth of the parts of the body by yet another expression, the ***Cellular Allometric Growth Equation***. In this fashion, the reduction in cell division leads directly to aging by the ***Gompertz Mortality Equation***. These findings give us insight into the underlying causes of these features of multicellular organization, as well as practical mitigations for managing growth, development, and the afflictions of old age.

In the accompanying manuscript,^26^ we shall see that ***Gompertzian Analysis***, counting cases and deaths, sorted by age, and then fitting these data to the ***Gompertz Mortality Equation***, allowed us to tease out the impact of vaccination, and other considerations, on COVID-19 lethality and infection. Taken together, these findings of ***Gompertzian Analysis*** and ***Cellular Phylodynamic Analysis*** provide deeper insight and practical tools for such tasks as comprehending COVID-19 mortality, quantifying how vaccination, and other forces, have reduced COVID-19’s lethal burden, and how the toll of COVID-19 infection, and other illnesses of old age, might be reduced even further.

## RESULTS

### Growth

For over two centuries, it has been appreciated that growth begins exponentially but then declines in rate.^5^ Many equations to capture such density-dependent growth have been considered,^6-12^ none of which accurately fit the growth of real animals.^13-15^ Rather than searching for another equation that fits growth, we carried out a ***Cellular Phylodynamic Analysis*** to examine what animal size data, in units of numbers of cells, ***N***, can tell us about the nature of the growth.^6^ To do so, we assembled values of numbers of cells, ***N***, from birth to maturity, for nematodes, frogs, chickens, cows, geese, quail, turkeys, mice, rats, fish, mollusks and humans (FIGURE 2, APPENDIX, and Reference 6). These data revealed, as expected, that at the beginning of life almost all cells are dividing, displaying the ***Cell Cycle Time, c***. Soon, however, the embryo steps back from such exponential growth, and we could make a rough measure of this mitotic decline by calculating the fraction of cells that would have to divide in order to account for the amount of growth that occurs at each moment in time, a value we call the ***Mitotic Fraction, m. m*** does not consider cell death, cell size, etc., but we shall see that it is a good start for addressing the questions at hand. The practical execution of ***m***’s calculation can be a bit involved, but details may be found in the APPENDIX and Reference 6. Strikingly, when we graphed the ***Mitotic Fraction, m***, versus animal size, ***N***, all ten animals listed above displayed the same relationship:

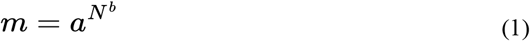

where the ***a*** and ***b*** parameters describe each animal’s specific growth qualities (FIGURE 2, APPENDIX, and Reference 6). We call Equation#1 the ***Universal Mitotic Fraction Equation***, as it describes the decline in the fraction of cells dividing, as measured by the ***Mitotic Fraction, m***, from ∼100% after conception, to very small numbers, as growth slows to adulthood. for all of the great range of animals we have examined.

**FIGURE 2:**
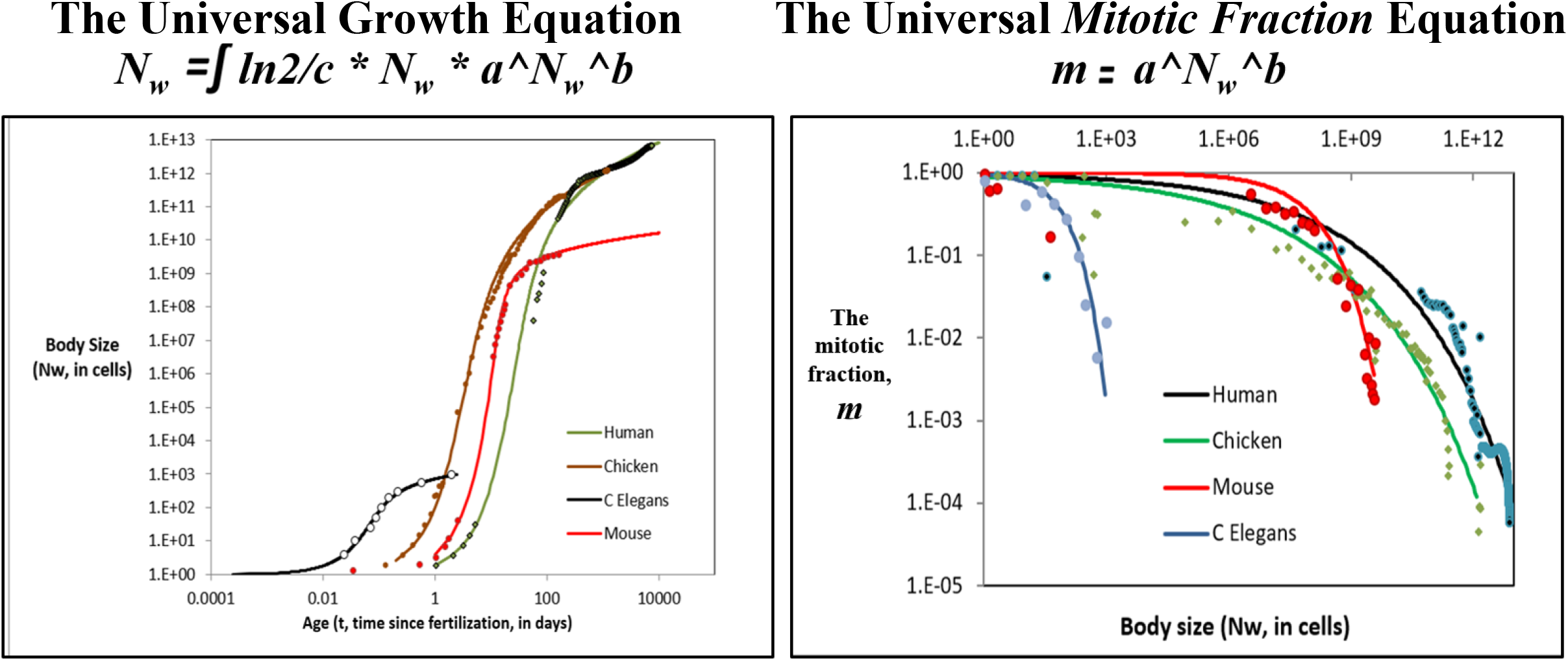
Growth data and the *Universal Growth Equation* and *Universal Mitotic Fraction Equation*. **for capturing growth from fertilization until maturity**. For additional data, see APPENDIX and Reference 6 **Left:** **Growth, in units of numbers of cells, *Nw*, in the whole body, from fertilization until maturity, in units of days, *t***. Datapoints for animal size’s (***N***_***x***_) versus time (***t***), for humans, chickens, ***C elegans*** nematode worms, and mice, and their fit to the numerically integrated form of the ***Universal Growth Equation*** (#2)^.6^ **Right**: **Estimate of the decline in the *Mitotic Fraction*, a measure of the fraction of cells dividing that occurs as animals increase in size, as estimated by the *Mitotic Fraction Method*, and captured by the *Universal Mitotic Fraction Equation***. Data points for animal size, ***N***_***w***_, in integer units of numbers of cells, vs. the ***Mitotic Fraction, m***, from fertilization, until maturity, for humans, chickens, mice, and ***C elegans*** nematode worms^.6^

It follows logically that the ***rate of growth*** of an animal reflects the ***Cell Cycle Time, c***, and the number of cells dividing, ***N*** and the ***Mitotic Fraction, m***, that is:

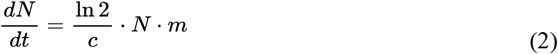

The relationship between size, ***N***, and age, ***t***, is captured by numerical integration of Equation #5, where the age, ***t***, at fertilization is ∼0, and the age at death is ***d***:

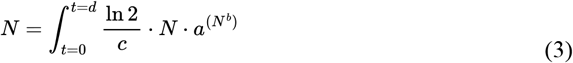

as well as by this closed-form integration:

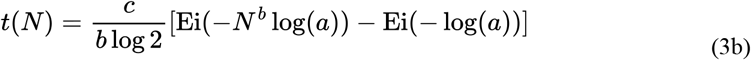

When tested against actual growth data, Equations #3 and #3b were found to closely capture the growth, from fertilization until maturity, for each of the species listed above, while none of the other widely used density dependent growth equations capture actual growth data as well (FIGURE 2, APPENDIX, and Reference 6). For these reasons, we call Equation #3 the ***Universal Growth Equation***, and Equation #3b the ***Inverse Universal Growth Equation***.

From a purely descriptive standpoint, the ***Universal Growth Equation***’s ***a*** and ***b*** parameters mold the “curviness” of the ***S-shaped*** growth curve, determining the sizes to which animals grow, while ***c*** expands or contracts the “***S*”** along the axis of time like an accordion, determining the speed at which they grow.

We do not know why growth occurs by this ***Universal Growth Equation***, but, curiously, for the idealized case of cells growing in a constant volume, such as an egg or a uterus, little more than the discrete allocation of inhibitory growth factor molecules binding to cells is enough to give rise to the ***Mitotic Fraction*** in the form of the ***Universal Mitotic Fraction Equation***. The mathematical demonstration of this finding can be seen in the APPENDIX and Reference 6, and the values of the ***a*** and ***b*** parameters reflect the thermodynamics of the growth factors at work.

The appearance of the ***Cell Cycle Time, c***, in the ***Universal Growth Equation*** is also a pleasant surprise. The length of time that it takes a cell to divide has long been known to reflect genome size,^6,27^ and ***c***’s appearance in the ***Universal Growth Equation***, reveals that junk DNA may have an unanticipated use in providing the speed-ballast for animal growth.

The same ***Cellular Phylodynamic Analysis***, that is to say, the examination of growth and development in units of numbers of cells, ***N***, has made it possible to identify several additional features of the formation of the animal body, which will aid in our analysis of aging (APPENDIX and Reference 6). Such an examination of the growth of many tissues, organs, and anatomical structures, in units of numbers of cells, ***N***, revealed that parts of the body frequently arise and grow by an expression we call the ***Cellular Allometric Growth Equation***^6^:

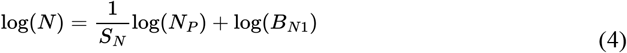

where ***N***_***p***_ is the number of cells in a cell lineage, tissue, organ, or anatomical structure of the body (APPENDIX and Reference 6). We call the parameter ***B***_***N1***_ the ***Cellular Allometric Birth*** and ***S***_***N***_, the ***Cellular Allometric Slope***.

This ***Cellular Phylodynamic*** approach also makes it possible to reconstruct plausible ***cell-lineage-trees*** of animals and their parts from the ***Universal Growth*** and ***Cellular Allometric Growth Equations*** and their parameters, by a method we call ***Cellular Population Tree Visualization Simulation*** (APPENDIX and Reference 6).

Finally, the ***Cellular Phylodynamic*** examination of various parts of the body reveals that they often mold their final composition by making ***Clones Within Clones***, in such cases as the relative growth of the front and back parts of the drosophila wing, the creation and growth of the organs of nematodes, the creation and maintenance of the crypts of the mammalian intestine, or the creation and clonal expansion of the cells of the immune system (APPENDIX and Reference 6). Indeed, ***Cellular Phylodynamic Clones Within Clones*** provides us with a mathematical framework around which we can build methods for optimizing COVID-19 vaccination impact, a point we shall return to in the DISCUSSION.

### Death

The relationship between ***age, t***, and ***Mitotic Fraction, m***, can be calculated by combining Equations#1 and #3: 

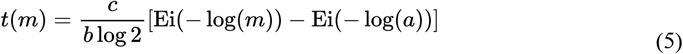

We call this expression, the ***Universal Mitotic Fraction in Time Equation***. With it, and the ***a, b***, and ***c*** values derived from the fit of growth data to the ***Universal Growth Equation*** (#3), we could generate curves, shown in FIGURE 3, for each of the 10 species listed above, ending at the age of the longest known individual in each species. These curves make clear that while life begins with virtually all cells dividing, at life’s end, more than 99.9% of the body’s cells are no longer engaged in mitosis (FIGURE 3). We call this boundary, marking where the ***Mitotic Fraction*** has declined from almost 100% at conception, to less than 0.1%, at the end of life, the ***Death Zone***.

**FIGURE 3:**
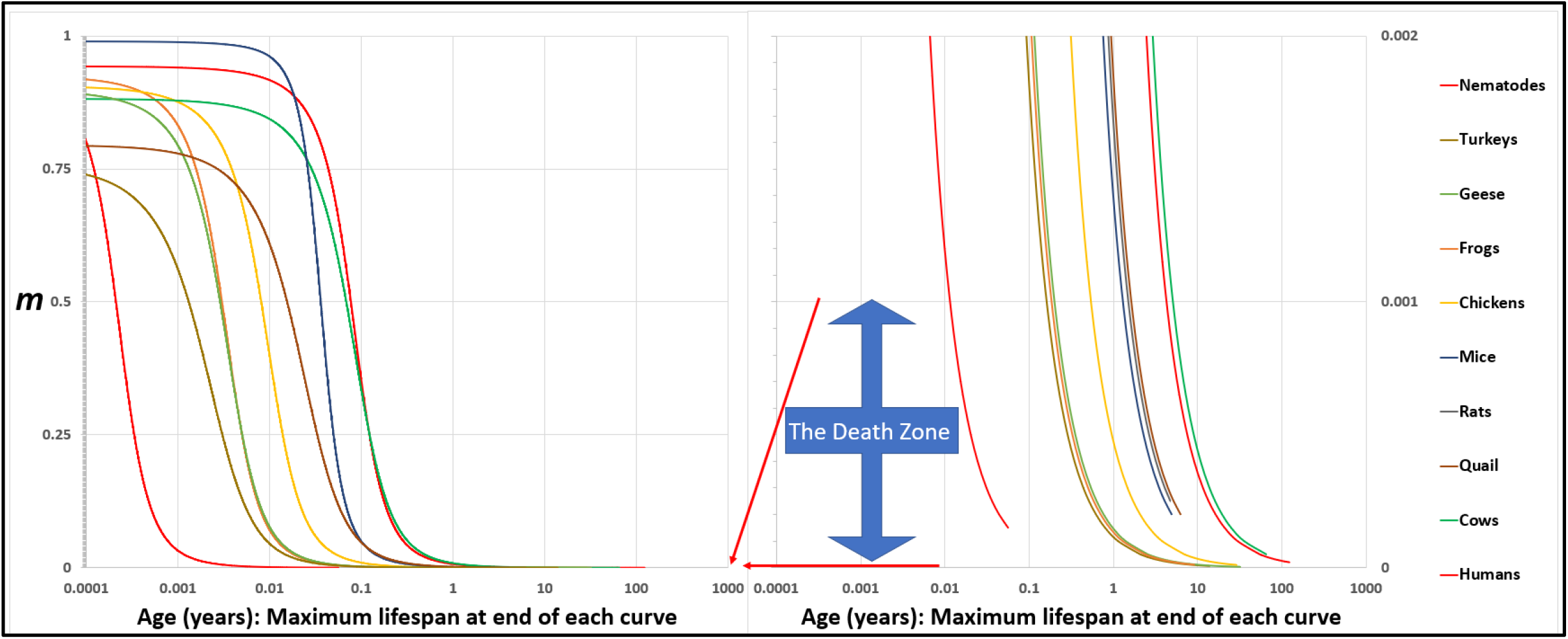
The *Universal Mitotic Fraction in Time Equation* (#5) captures change in *Mitotic Fraction, m*, with *Age, t*. 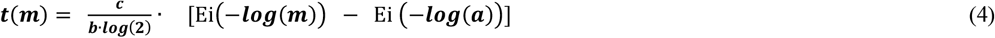

The graph on the right side shows the bottom 0.2% of the Y-axis of the graph on the left side: the ***Death Zone*** occurs when fewer the 0.1% of the cells of the body are dividing. Each curve ends at the age of the oldest known member of its species. Humans (♂) (Homo sapiens), Frogs (Rana pipiens), Nematodes (C. elegans), Chickens (Gallus gallus), Cows (Bos taurus), Geese (Anser anser), Mice (Mus musculus), Quail (Colinus virgianus), Rats (Rattus norvegicus), Turkeys (Meleagris gallopavo). See APPENDIX and Reference 6 for details.

### Dilution or Decay

Why would it be harmful for an animal’s ***Mitotic Fraction*** to dip to below one-in-a-thousand cells? Many irreversible chemical events, including protein denaturation, oxidation, and DNA mutation^28^, degrade cells, but most of this damage is flushed away by mitosis, when 50% or more of the cell’s components are made anew, and genome repair occurs.^29^ This concept of cell repair by dilution with new materials made at mitosis, ***Mitotic Dilution***, thus driving “the cause of aging and control of lifespan”, was articulated by Gladyshev,^30^ expanding Sheldrake’s general suggestion,^31^ and Hirsch’s mathematical examination.^32^ As Ferrucci noted: “Aging is the ratio between damage accumulation and compensatory mechanisms. … we can replace these molecules … new DNA, new protein, new organelles, and so on … [thus] … aging starts at conception.”^33^ This process of aging decline, by infrequency of mitosis, leading to insufficiency of dilution repair, from conception, until death, is captured quantitatively by the ***Universal Mitotic Fraction in Time Equation*** (#5).

### Lifespan

All three parameters of the ***Universal Growth Equation, a, b*** and ***c***, considered genetically, allow species to evolve to patterns of growth that give the greatest chances of survival, and to lifespans calculable with this expression:

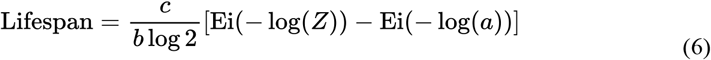

***Z*** is the ***Mitotic Fraction*** at the end of life. We call Equation #6 the ***Universal Lifespan Equation***, whose calculations, based on ***a, b***, and ***c*** values from growth data, distinguished between the longest lifespans (humans and cows), and the shortest (nematodes), lumping in between animals with 1-to-20-year lifespans (FIGURE 4). For the ten species we have examined, the average ***Z*** value of the average lifespan for each species, ***Z***_***A***_≈0.000037. The average ***Z*** value of the longest known lifespans, ***Z***_***L***_≈0.000024

**FIGURE 4:**
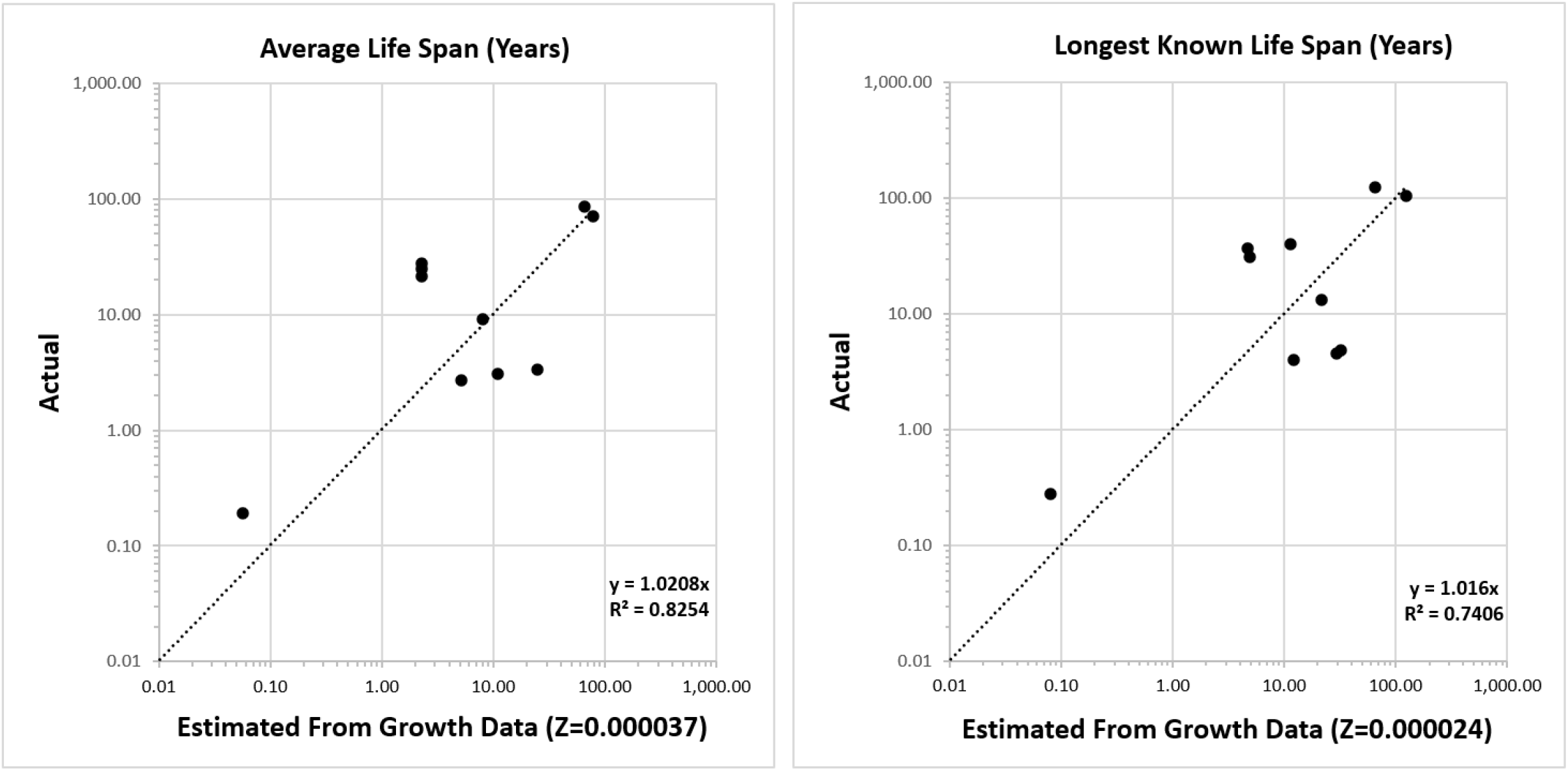
Life Span - Actual vs Predicted from Growth Data with the *Universal Lifespan Equation* (#6). Datapoints are for Humans, Frogs, Nematodes, Chickens, Cows, Geese, Mice, Quail, Rats, Turkeys. See APPENDIX and Reference 6.

### The Force of Mortality

In 1835, Gompertz reported that the chance of death for humans, ***D***, increases exponentially with age, ***t***, the ***Gompertzian Force of Mortality***, which describes the risk of death generally, and the risk of many diseases, including COVID-19^17^ (FIGURE 1):

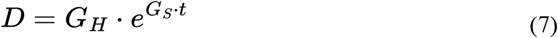

which is equivalent to: 

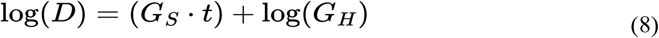

These are expressions of the ***Gompertz Mortality Equation***. We call ***G***_***s***_, the ***Gompertzian Slope***, and ***G***_***H***_, the ***Gompertzian Height***.

### Growth, Decay, and Aging

As we have seen above, the examination of body size in units of numbers of cells, ***N***, reveals that the rate of animal growth declines with age by the reduction in the fraction of cells that are dividing, by the ***Mitotic Fraction in Time Equation***. This decline begins gradually early in life, gains speed early in development, and then settles to a more regular rate as one enters adulthood. For example, the ***Mitotic Fraction in Time Equation*** tell us that after the age 50, the decline in the ***Mitotic Fraction, m***, appears like a slightly bent straight line (FIGURE 5), that is:

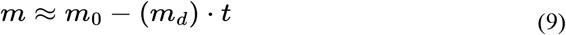

Where ***m***_***0***_ is the ***Mitotic Fraction*** projected back to the time of conception (***t*=0**, not biologically relevant, because the linearity in Equation #9 does not apply to early life, but mathematically convenient for studying old age), and ***m***_***d***_ is the approximate rate of decline in the ***Mitotic Fraction*** in old age.

**FIGURE 5:**
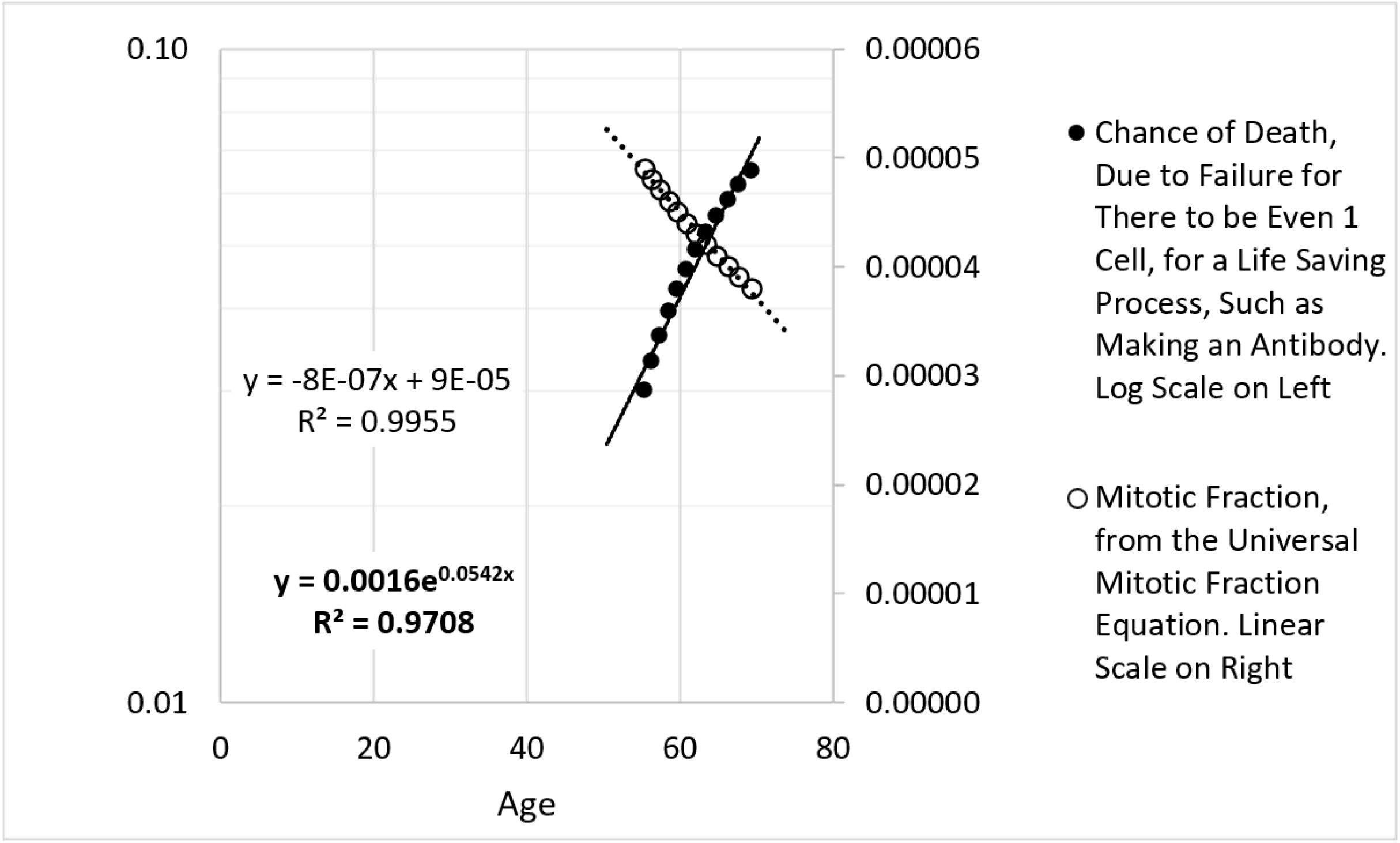

Consider that once a cell has divided, it will remain functional for ***F*** days, unless it divides again. It follows that a part of the body with ***N***_***p***_ cells will contain ***N***_***pf***_ functional cells:

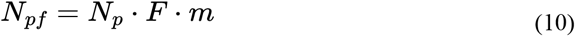

Combining Equations #9 and #10:

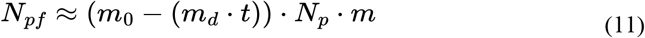

Body parts have specialized cells, doing different things.^19^ For example: in the pancreas, some cells make insulin, while others make digestive enzymes; in the heart some cells are muscle, others gristle; in the liver, some cells make blood proteins, others modify bile; etc.; etc. The immune system embraces this specialization with enthusiasm, with lymphocytes of many specialized types, starting from T-cells and B-cells, then subtypes, then yet more specialized cells as the results of massive cut-and-paste randomization of antibody and T-cell receptor genes, and then, somatic mutation, just to name a few. Generally, each cell of a part of the body has a probability, ***p***, of being able to do a certain task, where:

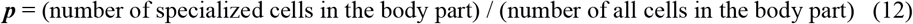

Let us consider the special case of ***p***_***a***_ as the chance of any single lymphocyte making an antibody to the COVID-19 virus spike protein. The body can make that antibody as long as it has at least one such functional lymphocyte with its genes arrayed to make that antibody. However, the ***Mitotic Fraction, m***, declines as we age, as seen in equation #9, the number of capable lymphocytes also declines, as seen in equation #11. Indeed, the chance of the body NOT having even one such capable lymphocytes, ***D***_L_, is:

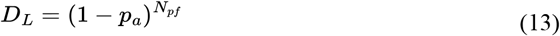

By plugging in Equation #11, which captures the approximately linear decline in ***Mitotic Fraction, m***, that occurs in late adult life, we arrive at:

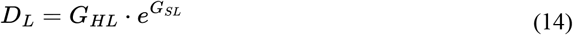

Which, remarkably, is the ***Gompertz Mortality Equation***, with:

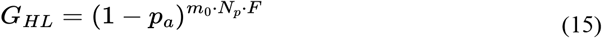

and:

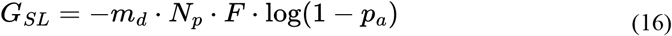

Thus, ***Cellular Phylodynamic Analysis***, the examination of growth in units of numbers of cells, ***N***, has allowed us to see that aging’s exponential ***Gompertzian Force of Mortality*** is the natural, and direct, result of the deceleration of growth that marks animal’s approach to adult size, as captured by the ***Universal Growth Equation***. For a detailed, and illustrative, closer look at this mathematical conclusion, the reader is referred to the APPENDIX.

Bear in mind that we have used lymphocytes making anti-COVID-19 antibodies to illustrate how the ***Gompertzian Force of Mortality*** will arise as a result of the decline in growth. However, this finding applies to lethality arising by failure of any type of specialized cell, in any of the body’s organs and tissues.

## DISCUSSION

### Counting Cells Has Told Us How We Grow and Why We Age

Simply counting cells has revealed that we reach adult size and old age by the same reduction in cell division: the ***Universal Mitotic Fraction Equation*** (#1). This provides a quantitative framework for comprehending how our lives come into being by the fundamental and ancient, invention of the control of cell division,^19-21^ as well as for comprehending how our lives come to an end by the inability of our cells to be rejuvenated by new materials made, and DNA repaired, at mitosis.

Of course, while ***Mitotic Dilution*** is a likely explanation for the relationship between growth and aging, even if some other biochemical cause is at work, the empirical relationship between growth and aging revealed by ***Cellular Phylodynamic Analysis*** remains mathematically sound. It is easily testable too. All that needs to be done is count more cells.

### Counting Cells Has Told Us How We Grow to Specific Sizes and What it Costs Us

The increase in susceptibility to illness that accompanies aging, the ***Gompertzian Force of Mortality***, is not the only cause of death. Predation, competition for food and space and metabolic efficiency, and a multitude of other forces impact selection, while animal size is a major determinant of adaptation to these forces. We grow to that fittest size by reducing the ***Mitotic Fraction*** to such a tiny rate that we do not seem to be getting any bigger, but the price of becoming that right size is decay and, ultimately, death.

Unlike humans, a few creatures, such as hydra, have learned to avoid the ***Death Zone*** with a “constant and rapid cell turnover”^34^, with surplus cells pushed out into new buds of life. Other apparent immortals, such as mole rats, may grow so slowly that we lose interest in waiting for them to become old, while others never stop growing; that is, they display indeterminate growth.^1^ However, most of us are fated by Darwinism to grow to sizes that maximize survival, achieved by the ***Mitotic Fraction*** declining into the ***Death Zone***.

### Counting Cells Has Told Us Why We Die of COVID and Other Things

Counting cells has taught us that as we age, fewer of our cells have experienced cell division recently, and thus fewer of our cells have been rescued from poisoning by ***Mitotic Dilution*** of the toxic compounds that accumulate naturally over time. Eventually, we run out of cells (Equations #9-16). For COVID-19, that means that as we age, our chance of having a lymphocyte that will defeat the virus fades exponentially, just as the chance of COVID-19 survival fades exponentially, and just as the chance of survival generally fades exponentially: the ***Gompertzian Force of Mortality***, as captured by the ***Gompertz Mortality Equation***.

Of course, such decay is not limited to the cells in the immune system. Aging robs us of functional cells in all of our tissues, organs, and anatomical structures. As a result, ***Cellular Phylodynamic*** analysis has the potential to explain why the chance of death from other sources of mortality, such as heart disease,^35^ increases as we age.

### Counting Cells and Cases Has Told Us How and Why COVID Becomes Less Deadly as We Immunize

As reported in the accompanying paper,^26^ by counting the numbers of cases and deaths by age, framed by the ***Gompertz Mortality Equation*** (#1), it was possible to tease out such clinical data measures of COVID-19 death (which we call ***Gompertzian Lethality***, Deaths/Cases), and COVID-19 infection (which we call ***Pasteurian Infectivity***, Cases/Population), a method we have called ***Gompertzian Analysis***. Such a ***Gompertzian Analysis*** of the numbers of cases and deaths has revealed that with each vaccination and booster, the chance of COVID death declines linearly, cumulatively pointing to zero death after 3 or 4 boosters, with no sign of waning.

***Gompertzian Analysis*** has also revealed that vaccine reduction in death (***Gompertzian Lethality***) and in infectiousness (***Pasteurian Infectivity***) appear to be separate features of the immune system. Fortunately, the math that led to Equation #11 gives us a way to count lymphocytes and see which cells of the immune system are behind COVID-19 ***Lethality*** and which cells are behind COVID-19 ***Infectivity***. When we find which cells in which locations come in numbers that match the ***Cellular Phylodynamic*** arithmetic of growth and death shown in FIGURE 5, we will have found the origins of COVID-19 ***Lethality***.

### Counting Cells May Help Us Get More Out of COVID Vaccines

Unfortunately, as reported in the accompanying paper,^26^***Gompertzian Analysis*** of COVID-19 cases and deaths has revealed that while sequential immunization has reduced the chance of COVID-19 death once infected (***Gompertzian Lethality***), it has not reduced the chance of infection (***Pasteurian Infectivity***) to the degree necessary for bringing the pandemic under control. Perhaps the problem lies in finding the way to optimize the immunization process, a task for which ***Cellular Phylodynamics*** might be of help. Such a ***Cellular Phylodynamic*** approach would essentially lie in counting cells in the immune system after vaccination to identify the equations that capture the number of immune cells that appear over time after immunization and, from these expressions, to estimate the amount of antibody produced. Such ***Cellular Phylodynamic*** math could then be used to identify, rationally, the optimal dosages, schedules, composition, adjuvantation, pharmaceutical augmentation, and other aspects of vaccine implementation at single moments in time and in the presence of evolving variants. Since current methods for doing this are surprisingly empirical and superficial,^36^ and for eradicating COVID-19, a failure, the possibility of developing a biologically sound, cell-number-based, ***Cellular Phylodynamic*** mathematics for optimizing COVID-19 vaccination would be appealing.

A ***Cellular Phylodynamic*** examination of COVID-19 immunization would build upon past successes in employing such cell-counting-centered approaches. We have seen here how a ***Cellular Phylodynamic Analysis*** of the growth of the body, counting cells, and using the numbers to find equations, has succeeded in revealing the ***Universal Mitotic Fraction Equation*** and the ***Universal Growth Equation***. Similarly, counting cells in parts of the body has succeeded in finding the ***Cellular Allometric Growth Equation***. Yet again, counting cells as we age has succeeded in revealing the basis of the ***Gompertz Mortality Equation***. Elsewhere, my colleagues and I have used ***Cellular Phylodynamic Analysis*** to identify much needed, biologically sound, testable equations for such things as how cancers becomes more lethal as they get larger, the ***SizeOnly*** Equation,^22-25^ then employing this ***Cellular Phylodynamic*** math to identify the most effective cancer screening schedules for savings lives, and then putting this knowledge to use in making national mammography screening recommendations.^25^ This cell-counting based approach to calculating cancer mortality exemplifies what we could achieve with a cell-based-approach to comprehending COVID-19 vaccine optimization, as the ***SizeOnly*** Equation could be expanded to calculate the impact of an enormous variety of prognostic factors into a single outcome measure,^22^ just as vaccine mathematics would be designed to calculate the impact of an enormous variety of vaccine implementation options into the single outcome of maximal infection control.

We appreciate that for most of the problems noted in the previous paragraph, the ***Cellular Phylodynamic*** method was brought to bear on biological entities that arise from single founder cells -whole bodies, body parts, and tumors-while the immune system contains a multitude of clones arising from countless founder cells. Fortunately, as outlined in the APPENDIX, the ***Cellular Phylodynamic*** method can also be extended to biological entities, such as the immune system, that are made of many clones, a method we have cells ***Clones-Within-Clones***. We also appreciate that for the cases noted above, the equations only increase in number with time. However, for immune responsiveness, we need to find expressions that comprehend how the cells increase in number, thus increasing antibody production, and then decrease in number, thus comprehending immunological waning. Finally, we also appreciate that the analysis suggested here is vague as to precisely which cells of the immune system should be counted, and indeed, the approach is equally applicable to many of the variety of cells that comprise the immune system.

Of course, such an undertaking can begin in mice and then be extended to humans, much as the ***Universal Growth Equation*** applies to both species, with specific growth qualities of each species captured by the values of the parameters ***a, b***, and ***c***. Mice have been used to examine COVID-19 vaccination^37^ and whatever findings might be observed in mice could obviously be extended to human materials.

### Life History, Lifespan, Evolution

Animals grow at different speeds, to different sizes, and different lifespans. The ***Cellular Phylodynamic*** mathematics described above show us how this variety, and the genetic variation leading to this variety, can be captured by the ***a, b***, and ***c*** parameters of the ***Universal Growth*** and ***Lifespan Equations***. These qualities go to the heart of evolution, as captured by life history theory.^38^ For example, the time till maturity and reproduction is determined by the curviness of the “***S*”** in the ***S-shaped*** growth curve of the ***Universal Growth Equation*** (#3); the “***S*”** can stand tall, rapidly growing towards adulthood, or slouch languidly, slowly edging away from adolescence, all determined by the ***a, b***, and ***c*** parameters, together with lifespan, characterized by the ***Universal Lifespan Equation*** (#6).

The ***a, b***, and ***c*** parameters are also polymorphic pleiotropic genetic determinants, with positive impacts on growth early in life and negative impacts on survival later, conforming to thee antagonistic pleiotropy theory of Williams^3^ and Hamilton^4^ and the disposable soma theory of Kirkwood.^39^

A species may also delay entry into the ***Death Zone*** by enlisting apoptosis to boost the ***Mitotic Fraction*** or investing in enzymatic repair. This may account for some species-specific variability in the value of the ***Z*** parameter below the one-in-a-thousand ***Death Zone*** border we have marked off. Medawar’s mutation accumulation theory^2^ provides a framework for comprehending the dilemma; is it worth the effort, as measured by Darwinian selection, to fight off the inevitable?

### Aging is Reversible

The cellular damage that occurs when the ***Mitotic Fraction*** enters the ***Death Zone*** is not irreversible. Germ cells appear to rejuvenate by resetting the ***Mitotic Fraction*** to ∼100%, a region called ***Ground Zero***, adopting the terminology and insight of Gladyshev^40^, and the mathematics of the ***Universal Mitotic Fraction Equation*** (#1) when ***N*≈0**. A similar process may rejuvenate embryonic stem cells and cells used to clone animals.^41^

### Growth and Aging Are Modifiable

Genetics, diet, pharmaceuticals, and other interventions have been found to change lifespan, often accompanied by altered growth. At the same time, large animals often live longer than small animals, and many genetic changes have been found to affect both growth and lifespan (reference 42, and others scattered throughout this report). These opportunities can now be examined quantitatively, with the ***Universal Growth*** and ***Universal Lifespan Equations*** offering both a mechanism and a mathematics for comprehending and testing aging’s link to growth.

### Aging and Health

For humans, ***Z*** (Equation #6) provides a measure of the medical and public health system’s capacity to deal with vulnerability induced by aging’s reduction in ***Mitotic Fraction***. This frames practical questions: Do we search for actions and treatments that delay ***Mitotic Fraction*’*s*** approach to the ***Death Zone***, or do we look for ways to remain healthier after the ***Mitotic Fraction*** enters the ***Death Zone***, learning how to live longer with the burden which growing poses to aging?^43^

### Anti-Aging Therapeutics

Many compounds have been found to extend life in a variety of animals, most relevantly to us, in mice, a few of which are beginning to be tested in humans.^44,45,46^ So, why haven’t we found the pill that will make us live longer? A plausible difficulty has been the inability to predict and test the impact that a candidate agent might have on the extension in life and the negative consequences associated with the pharmaceutical so that the right agent, at the right dose, and the right schedule, could be identified and put to work to achieve a reasonable and predictable lifespan extension with minimal negative consequences. The ***Cellular Phylodynamic*** mathematics of growth and aging outlined here should help fill that gap.

For those agents found to change growth and aging together, identifying which parameter of the ***Universal Growth*** and ***Universal Lifespan Equations*** is changed, that is, ***a, b*** or ***c***, should provide actionable information since each parameter can be expected to have different therapeutic effects and different adverse side effects. This can be measured in nematodes in days and in mice in months by fitting survival and growth data to the ***Universal Growth*** and ***Universal Lifespan Equations***. These equations can then be used to identify the optimal dose and schedule. They can also guide the search for new agents and inform the synthesis of new analogs with the appropriate pharmacological qualities of affinity, half-life, and slow release.

Pharmaceuticals that are found to change growth by ***a*** or ***b*** may bump up the ***Mitotic Fraction*** and reverse aging, returning the body to the vigor of a younger age but also increasing body size, that is, making the body acquire more cells, which will probably be harmful if used for long periods. However, such agents should be helpful for temporarily reversing aging, that is, for short-term increases in the ***Mitotic Fraction***, for such tasks as improving the impact of vaccines, for which COVID-19 comes to mind.

Pharmaceuticals that are found to slow growth by ***c*** will slow the decline in the ***Mitotic Fraction***, thereby stretching out ***Universal Lifespan Equation*** and slowing aging. They will not reverse the body to a younger state, but they will have the advantage of not having the potentially harmful adverse effects of increasing body size. Agents such as rapamycin-related compounds might be expected to have this quality, a possibility that should be examined experimentally. Identifying the magnitude of the impact of such agents on the ***c*** parameter would make it possible to predict the dose/outcome relationship by re-calculating the ***Universal Growth*** and ***Universal Lifespan Equations***. Such possibilities are testable in mice.

### Measuring Age and its Amelioration

Various laboratory measures of age have been developed,^47,48^ although the ultimate and causal metric might well be the ***Mitotic Fraction*** itself. By counting mitotic figures in histological sections, the ***Mitotic Fraction*** can be determined for single points in time, while measurements of the sizes of the clones in the liver marked by the activation of plasma protein genes, which occurs at a regular rate,^21^ can provide life-long values. These direct ***Mitotic Fraction*** measurements should also help provide rapid screening and testing of pharmaceuticals for anti-aging activity.

### Dilution, Mutation, Cancer

The features of cellular decay tell us that the mechanism of ***Mitotic Dilution*** is not a theory of aging, but a mechanism of all theories of aging, with the ***Universal Mitotic Fraction in Time Equation*** providing a measure for how fast all these processes go downhill. ***Mitotic Dilution*** is also unlikely to be the only mechanism of aging. In a recent technical tour de force, Cagan and colleagues found that the somatic mutation rate, as measured in the intestine, scales with lifespan.^28^ This effect, no doubt protective against cancer, is unlikely to be the result of the decline in the ***Mitotic Fraction***, as the cells in the intestine undergo a rate of mitosis vastly greater than the ***Mitotic Fraction*** in the body as a whole (although one point of view holds that the precursors of the crypt stem cells may have a more conventional level of mitotic activity^49,50^). As the authors concluded, “The most notable finding of this study is the inverse scaling of somatic mutation rates with lifespan. … Even if clear causal links between somatic mutations and aging are established, aging is likely to be multifactorial. Other forms of molecular damage involved in aging could be expected to show similar anticorrelations with lifespan”. The authors also point to their own superb, recent study^51^, which found that individuals with a genetic predisposition to DNA mutation display an increased risk of cancer but not a pronounced reduction in life expectancy.

Cancer frequently occurs late in life and thus has a surprisingly minor contribution to life expectancy. Gandjou has calculated that even if we found the imaginary cure for cancer, life expectancy at birth would only increase by about 3.25 years.^52^ Of course, animals with long lifespans would never reach their potential if they displayed cancer incidence rates seen in animals with short lifespans, and Cagan and colleagues have provided a long-sought answer to this previously mysterious problem. The ***Cellular Phylodynamic*** mathematics outlined here should provide a starting point for sorting out the contributions of growth, and other factors, such as mutation and cancer, to overall survival.

### Growth and Death

The ***Cellular Phylodynamic Analysis*** of the data utilized here, that is, examination in units of numbers of cells, ***N***, suggests that animal growth and death are inescapably bound together, both arising from the same taming of mitosis. At the end of life, our now minuscule and plummeting ***Mitotic Fraction*** may no longer be perceptible in the mathematically negligible increase in size suggested by the ***Universal Growth Equation***. However, it will be perceived by the increase in our frailty suggested by the ***Universal Lifespan Equation***. Thus, the size to which we grow, and the time at which we die, go hand in hand, both driven by the same reduction in the fraction of cells dividing. While the reward for this reduction in the ***Mitotic Fraction*** in space and time is the ability to grow to specific shapes and sizes^20^, the cost is the disability of cells starved of life-sustaining replacement materials made at mitosis, as we have seen for the age-associated susceptibility to COVID-19 and other pathogens. Counting our cells can help us find new ways to hold back that bitter hug of mortality.

Note: this study has not been peer reviewed, and the findings could change.

## Data Availability

All raw data in this paper was taken from other studies, and is readily available from those studies, which have been cited.

