## APPENDIX for "Counting Cells by Age Tells Us About How, and Why, and When, We Grow, and Become Old and Ill"

### Analysis, Data, and Mathematics, in Full Detail

**James Michaelson PhD**

Department of Pathology, Massachusetts General Hospital
Department of Surgery, Massachusetts General Hospital, USA
Department of Pathology, Harvard Medical School, USA
15 DEER COVE ST UNIT # 2
LYNN, MA 01902-3119
United States
TEL (cell phone) 617 501 0590


**TOC**

***CELLULAR PHYLODYNAMICS***

The ***Cellular Phylodynamics*** of growth

***Cellular Phylodynamics*** is a mathematics of the discrete nature of cells and the cellular events that cause change in cells.

The ***Computational Animal***

Current concepts in growth and development

***UNI-GROWTH***

Data for the ***Cellular Phylodynamic Analysis*** of the growth of the whole body.

Basic features of growth and cell division: ***exponential growth, biotic potential***, the ***Mitotic Fraction***, and ***quiescence***

The ***Mitotic Fraction Method***, where the value of the ***Mitotic Fraction***, ***m***, is measured over the full range of growth from fertilization until maturity.

As animals grow in size, $N_w$, the ***Mitotic Fraction***, ***m***, declines rapidly.

As animals grow in size, $N_w$, the declines in the ***Mitotic Fraction***, ***m***, is well fit by the ***Universal Mitotic Fraction Equation***, $m = a^{(N_w^b)}$

The relationship between size, $N_w$, and age, ***t***, is well fit by the ***Universal Growth Equation***: $\int \left(\frac{\log(2)}{c}\right) N_w a^{(N_w^b)}$

Closed form solution to the integration of the ***Universal Growth Equation***

Density dependent growth equations

The relationship of the ***Mitotic Fraction*** (***m***) and size ($N_w$) to the ***logistic equation***.

The relationship of the ***Mitotic Fraction*** (***m***) and size ($N_w$) to the ***Gompertz equation***.

The relationship of the ***Mitotic Fraction*** (***m***) and size ($N_w$) to the ***West equation***.

Fit of the relationship of the ***Mitotic Fraction*** (***m***) and size ($N_w$), to the ***Gompertz***, ***logistic***, and ***West equations***.

Fit of the relationship between size ($N_w$) and Age (***t***) to the ***Gompertz***, ***logistic***, and ***West equations***.

Fine scale "***ripples***" identify a lower level of growth detail that occurs in the life of the organism.

The Meaning of ***UNI-GROWTH***

The ***Universal Growth Equation*** describes how growth occurs

The ***Universal Growth Equation*** is linked to the cell biology behind growth.

The ***Universal Growth Equation's*** description of how fast we grow is linked to the ***Cell Cycle Time***, ***c***, which is linked to genome size, which is linked to how much junk DNA we have.

The ***Universal Growth Equation's*** description of the fraction of cells dividing, the ***Mitotic Fraction***, ***m***, is linked to the action of cell signaling molecules that induce mitotic quiescence.

***UNI-GROWTH***: Summary Definition

***ALLO-GROWTH***

***The Cellular Phylodynamic Analysis*** of the growth and development of the parts of the body

The ***Allometric Growth Equation***

***Cellular Allometric Growth Analysis***

Early development: ***Cell lineages***: ***Cellular Allometric Lineage Growth Analysis***:

Late development: ***Tissues***, ***Organs***, and ***Anatomical structures***: ***Cellular Allometric Batch Growth Analysis***:

The Meaning of ***ALLO-GROWTH***, and its defining abstraction, the ***Cellular Allometric Growth Equation***

The ***Cellular Allometric Growth Equation***

The ***log-linearity*** of body part ***ALLO-GROWTH*** can be traced to whole body ***UNI-GROWTH***

Parameter $S_N$, the ***Cellular Allometric Slope*** of the ***Cellular Allometric Growth Equation***, tells us how the embryo uses the ***Cell Cycle Time*** to drive ***Cellular Selection*** to adjust the size of each part of the body:

Parameter $S_N$, the ***Cellular Allometric Slope***, at work in adjusting the size of each part of the body:

Parameter $S_N$, the ***Cellular Allometric Slope***, at work early in development

Parameter $S_N$, the ***Cellular Allometric Slope***, at work late in development

Parameter, $S_N$, the ***Cellular Allometric Slope***, $S_N$, and the ***Cell Cycle Time***, $c_p$,

The role of DNA methylation on the time and nature of change in the ***Cell Cycle Time***, $c_p$ within the body

Parameter, $B_{N1}$, the ***Cellular Allometric Birth*** of the ***Cellular Allometric Growth Equation***, tells us how embryos create body parts from single cells

Data on the number of ***Founder Cells*** that make various parts of the body.

When the parts of the body are born: The ***Cellular Allometric Birth in Time***, $B_T$

The time of the ***Cellular Allometric Birth*** of a body part affects its size.

Body parts can be made of one ***Founder Cell*** or more than one ***Founder Cell***.

Body Part Size to Body Part Size Relationships; ***Body Part Proportion Equations***

***Clones Within Clones***

***ALLO-GROWTH***: Summary Definition

***AGING***

#### ***CELLULAR PHYLODYNAMICS***

In the main text of this communication, we presented a condensed description of a new method, ***Cellular Phylodynamics***,[1] and its findings, focusing on aging, by which we mean the consideration of growth, development, and aging in terms of numbers of cells, $\boldsymbol{N}$.[2] The equations derived from this approach for animals generally are summarized in **BOX 1.** Equations in the text are given numbers less than 100, while the additional equations introduced in this APPENDIX begin with 100. Related equations have a letter following the number (11b), followed by a number (11b1), etc.

In this APPENDIX we provide a more detailed and thorough report of this analysis. This includes derivations of all of the equations for the growth, development, and aging of animals and humans, and a more complete presentation of the growth data, which is exhaustive.

##### The ***Cellular Phylodynamics*** of growth

The ***Cellular Phylodynamics*** approach is based on counting the number of cells in the animal as a whole, $\boldsymbol{N_w}$. and in its various anatomical parts, $\boldsymbol{N_p}$. These values of $\boldsymbol{N_w}$ and $\boldsymbol{N_p}$ provide the raw material for mathematical analysis, from which equations could be discerned. Thus, the equations that emerge from a ***Cellular Phylodynamic Analysis*** of growth, development, and aging aren't mathematical inventions, describing imaginary humans and animals, but empirically based generalizations of actual growth and development, which occur in actual animals, including ourselves.

***Cellular Phylodynamics*** is a mathematics of the discrete nature of cells and the cellular events that cause change in cells.

We, and almost all animals, begin life as a single cell, which divides to become 2 cells, then 4 cells, then 8, growing to the trillion or so cells of a human[3] or the thousand or so cells of a nematode worm[20], comprised of ***cell lineages***, ***tissues***, ***organs***, and ***anatomical structures***, each of which grows to its recognizable size. How does this occur? At the microscopic scale, our cells are intrinsically discrete entities.[83] There is no such thing as 1/3rd of a cell or 1.33 cells. It follows that the number of cells in the body, or in any part of the body, can only change their integer numbers by binary events, such as 1 cell becoming 2 cells by mitosis, or 1 cell becoming 0 cells by cell death. We can, and will, avail ourselves of continuous mathematics when it can provide us with useful approximations, but the ***Cellular Phylodynamics*** of embryology is fundamentally discrete in nature.

##### The ***Computational Animal***

Since cells only come in integers, whose numbers only change by discrete events[83], multicellular animals remind us of computers, also comprised of integer units,[4] which have also been called "cells"[5], whose states also undergo discrete changes. As we shall see, ***Cellular Phylodynamics*** provides us with a way to understand ourselves as ***Computational Animals***, whose growth, development, and aging can be understood as the aggregate consequences of the many discrete changes that occur among our cells.

##### Current concepts in growth and development

In contrast to the discrete nature of animal life at the microscopic scale, from our macroscopic perspective the growth of the body and its parts appears in continuous qualities of length, volume, mass, and shape. These continuous features of growth have yielded to quantitative analysis, although not always in ways whose meanings are clear. For example, it has long been appreciated that ***as we grow older***, ***we grow bigger***, and ***we grow slower***, ***until growth becomes imperceptible***, and a number of ***density-dependent*** growth equations have been developed (the ***logistic***, ***Gompertz***, ***von Bertalanffy***, ***Richards***, ***West***, and other equations: see below), which capture this increase in size and decline in the speed, and form classic ***S-shaped growth curves***.[6-11] Unfortunately, none of these ***density-dependent*** growth equations accurately fit the growth of real animals.[12-14] It has also long been appreciated that as we grow, the sizes of the ***tissues***, ***organs***, and ***anatomical structures*** of the body, when compared with the size of the body as a whole, often form straight lines on log-log graphs, a phenomenon known as ***allometric growth***.[15-19] However, the reason behind the striking log-linearity of ***allometric growth*** has long been a mystery. Finally, many studies have used 4-dimensional microscopy to characterize the growth of ***cell lineages*** that arise from single ***Founder Cells*** at the beginning of development,[20,21] but the precise way in which the embryo uses the control of cell division to create these first body structures has been obscure.[22-24] ***Cellular-Selection***, that is, differential cellular proliferation and death, creates many anatomical structures by outgrowth from the undifferentiated body mass, and molds much of the fine scale detail of anatomy, although how cell division is harnessed to achieve this is obscure.[98,99] Cell lineages grow from individual ***Founder Cells***,[20] although how embryos use mitosis to create these parts, and how these ***Founder Cells*** undergo ***Cell-Heritable*** change that characterized each clone, remains unknown. The analysis of cellular diversification, by single cell mRNA expression,[25,]cell marking,[26,] and high resolution 4D microscopy,[22,23,102] has also wrestled with the problem of cell lineage formation. As we have seen in the main text of this communication briefly, and will present in greater detail below, when we re-examine these features of growth, development, and aging in units of numbers of cells, that is, by ***Cellular Phylodynamics***, the biological basis for many of these processes becomes clear.

***UNI-GROWTH***

Data for the ***Cellular Phylodynamic Analysis*** of the growth of the whole body.

To carry out the ***Cellular Phylodynamic Analysis*** of growth, we assembled data on the number of cells in the whole animal, $N_w$, by age, $t$, in days from fertilization until maturity, for 13 species of animals: zebrafish (***Danio rerio***),[27] European sea bass (***Dicentrarchus labrax***),[28,29] mice (***Mus musculus***),[30,31] rats (***Rattus norvegicus***),[32] cows (***Bos taurus***),[33,34] humans (***Homo sapiens***),[35,36,37] bobwhite quail (***Colinus virgianus***),[38,39] domestic chickens (***Gallus gallus domesticus***),[40,41] turkeys (***Meleagris gallopavo***),[42] geese (***Anser anser***),[38] nematode worms (***C. elegans***),[20] frogs (***Rana pipiens***),[43] and clams (***Merceneria mercenaria***).[44,45,46] Data on the numbers of cells in early embryos, from fertilization onward, were available for mice,[47,48] rats,[49] cows,[50] humans,[51] chickens,[52] turkeys,[53] nematode worms,[20] frogs,[54] clams,[45] and fish.[55,56] Data on the numbers of cells in later embryos were assembled from published values in units of weight or volume by taking advantage of the finding that there are about $10^8$ cells/gram-cc.[57] Excel files (TABLE A1) characterizing these basic growth data ("**Basic Data Files**"), and the calculations made from these data ("**Calculations Files**"), are available on request. Also available to interested readers are excel files with the data on the growth of the parts of the body parts that we examined (see below).

The graphs of these various datapoints for animal size's ($N_w$) versus time ($t$), are shown in FIGURES A1-A3. Graphing the number of cells in the body as a whole, $N_w$, vs. age, $t$, from fertilization until maturity, reveals, for each of these 13 species, that animal growth occurs by ***S-shaped*** growth curves (Fig A1). This is most easily viewed on log-log plots, because the greatest part of growth occurs rapidly at the beginning of life. For example, in terms of numbers of cells, $N_w$, humans grow about 11 orders of magnitude in the womb and about 2 orders of magnitude after birth. As we shall see below, the reason why all of these creatures have similar ***S-shaped*** growth curves is that they all grow by an equation of the same form, the ***Universal Growth Equation***.

Basic features of growth and cell division: ***exponential growth, biotic potential***, the ***Mitotic Fraction***, and ***quiescence***

Because cells increase in number by cell division, they are capable of exponential growth, in which the size of an organism, $N_w$ (in terms of cell number), with age, $t$, can be thought of in terms of its ***rate of growth***:

$$\frac{dN_w}{dt} = \frac{ln(2)}{c} \cdot N_w \qquad (100)$$

where $c$ is the average ***Cell Cycle Time*** among those cells that are dividing, which, for exponential growth, is all cells.

Integration of Equation #100, translates the integer nature of cells into the continuous appearance of growth:

$$N_w = e^{\left(\frac{ln(2)}{c}\right)\cdot t} \qquad (100b)$$

Thus, data on the relationship between cell number, $N_w$, and time, $t$, at the very beginning of development, when all cells are dividing, can give one a practical measure of the animal's ***Cell Cycle Time***, $c$.

Of course, exponential growth (Equation #100b) is ultimately unsustainable, a fact captured by the concept of the ***biotic potential***.[58] The departure from the ***biotic potential*** of our cells is a fundamental feature of animal growth, which can be captured by introducing the term $m$, the ***Mitotic Fraction***, giving us this expression for the ***rate of growth***:

$$\frac{dN_w}{dt} = \frac{ln(2)}{c} \cdot N_w \cdot m(N_w) \qquad (100c)$$

Equation #100c embraces all forms of growth, but without specificity, as the manner by which the ***Mitotic Fraction***, $m(N_w)$, changes with the animal size, $m(N_w)$, is completely unspecified. As we shall see below, this lack of specificity gives the Equation #100c the power to accomplish such things as leading to a way to calculate how the ***Mitotic Fraction***, $m$, changes as we grow (see the ***Mitotic Fraction Method*** below). Additionally, insertion of the ***Mitotic Fraction***, $m(N_w)$, into the exponential growth equation (#100b), converts it into Equation #100c, and gives us the general form of a ***density dependent growth*** equation, which balances the unlimited quality of exponential growth ($\frac{ln(2)}{c} \cdot N_w$) with the inevitable limit of resources ($m(N_w)$).

Although our treatment of the ***Mitotic Fraction***, $m$, relies on an obsessively rigorous adherence to this mathematical definition in Equation #100c, from time to time we will revert, for the purposes of clarity, to speaking of $m$ as a rough measure of the ***growth fraction***, $G$, the fraction of cells dividing, which is usually measured by the cytogenetic analysis of the uptake of DNA precursors.[59] The two abstractions of ***Mitotic Fraction***, $m$, and ***growth fraction***, $G$, are obviously closely, but not precisely, related, since other factors (cell death, cell size, cell density) play roles in growth that are likely to be secondary to cell division. Definitionally, the ***Mitotic Fraction*** is a measure of the amount of actual growth that occurs as a fraction of the growth that would have occurred had each cell divided, ignoring other effects, such as cell death and cell size, a plausible approximation, as mitosis is growth's principal driving force. Thus, the ***Mitotic Fraction*** $m$ can be thought of as the main component of the ***growth fraction***, $G$, the precise fraction of cells that are dividing. Operationally, this imprecision has little practical impact, as it will be cancelled out when we reintroduce $m$ into new equations of growth that we shall derive below.

When we think imprecisely of the ***Mitotic Fraction***, $m$, as the fraction of cells dividing:

$$m \approx N_{Wd}/(N_{Wd} + N_{Wq}) \qquad (100d)$$

where $N_{Wd}$ is the number of cells dividing and $N_{Wq}$ is the number of cells that have ceased from dividing, a process known as ***mitotic quiescence***. ***Quiescence*** is the complex biochemical mechanism that occurs within cells to prevent them from progressing to mitosis, which is set in motion by inhibitory and stimulators growth factor molecules, hormones, and other signals that cells use to communicate.[60-62]

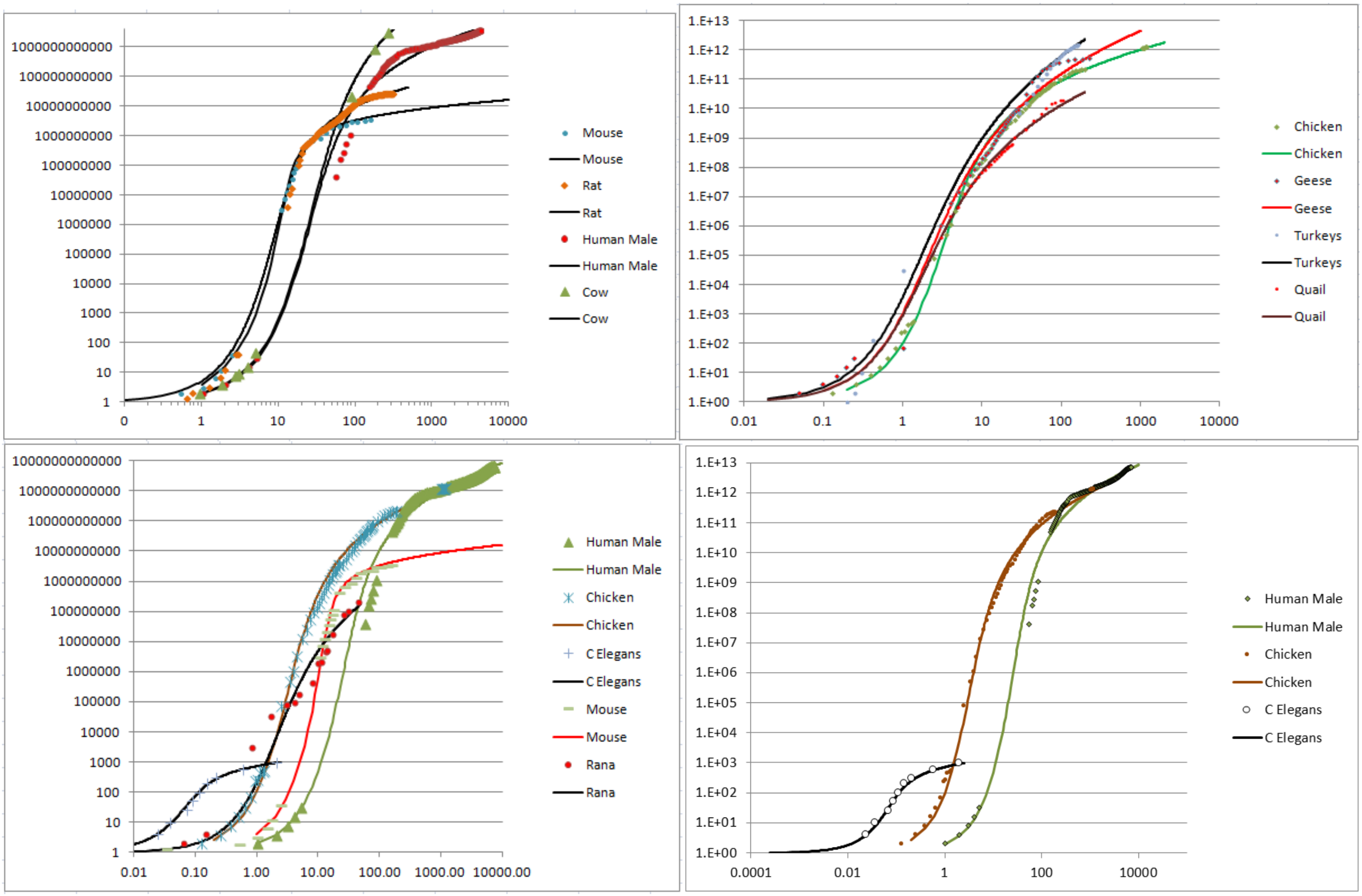


**FIGURE A1**: **Growth, in units of numbers of cells, $N_w$, from fertilization until maturity, in units of days, *t*.** Datapoints for animal size's ($N_x$) versus time (*t*), for ***C elegans***, chickens, mice, turkeys, quail, geese, frogs, and humans, and their fit to the numerically integrated form of the ***Universal Growth Equation*** (#5b)**.**

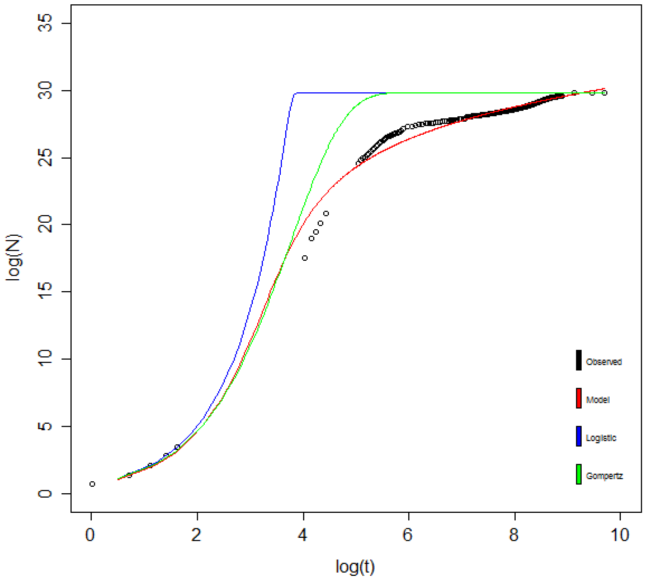


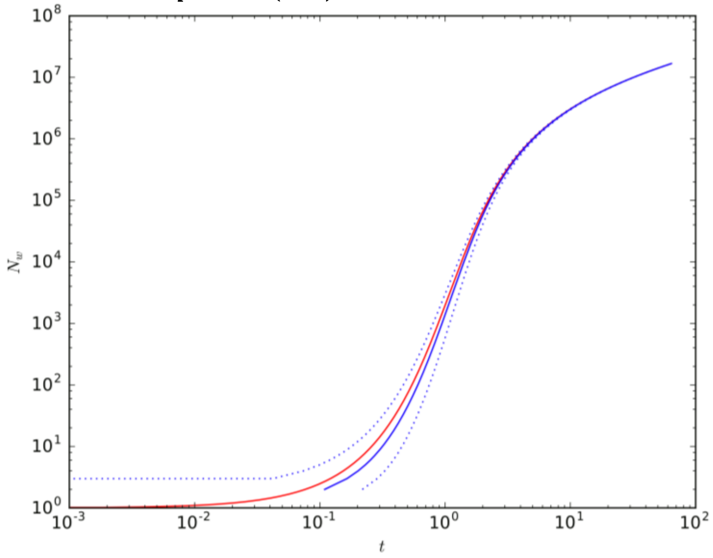


**FIGURE A2**: **Fit of human growth to the *Universal Growth Equation* (red), the *logistic equation* (blue), and the *Gompertz equation* (green).** Data values as black circles.

**FIGURE A3:**
**Analysis of the closed form integration of the *Universal Growth Equation* (#3).**

The ***Mitotic Fraction Method***, where the value of the ***Mitotic Fraction***, ***m***, is measured over the full range of growth from fertilization until maturity.

The value the ***Mitotic Fraction***, ***m***, for animals of various sizes, $N_w$, can be calculated by reversing ***Equation*** #100c:

$$m = \frac{dN_w}{dt} \,/\, [\frac{ln(2)}{c} \cdot N_w] \tag{3b}$$

We call the practical implementation of this equation the "***Mitotic Fraction Method***". To execute these ***Mitotic Fraction Method*** calculations, we use our assembled growth data to calculate the value of the ***Mitotic Fraction***, ***m***, (column M of the excel files listed in "**Calculations Files**" column of Table 1) by:

$$m_i = \frac{2N'(t_i)}{r(N_i+N_{i+1})} \tag{101}$$

The same iteration was also carried out based on an exponential calculation, rather than on the linear calculation show above, yielding essentially the same outcome (not shown).

Equation #101 occasionally resulted in a small number of data points with $m > 1$. Since this is both theoretically and numerically impossible (the biological meaning would be one cell dividing to give rise to more than two cells, which is nonsensical), these values of ***m*** were manually overwritten with a value of $m = .999$ noted by orange highlights in the excel spreadsheets.

As animals grow in size, $N_w$, the ***Mitotic Fraction***, ***m***, declines rapidly.

Our ***Mitotic Fraction Method*** calculations, based on the animal growth data noted above (Table 1), characterized how the value of the ***Mitotic Fraction***, ***m***, declines as growth occurs, that is, as $N_w$ increases, from fertilization until maturity, for nematodes, chickens, cows, geese, quail, turkeys, fish, clams, mice, rats, humans, and many other animals (FIGURES A4-A6). Graphs of the values of the ***Mitotic Fraction***, ***m***, calculated by the ***Mitotic Fraction Method***, for each of the diverse species of animals noted above, reveal that growth is close to exponential during the first few cell divisions ($m \approx 1$), but soon begins its "slippery slope" descent, resulting in a rapid and accelerating decline in the value of ***m***, reaching values of $10^{-2}$ to $10^{-6}$ by the time adult size is reached (FIGURES A4-A6).

As animals grow in size, $N_w$, the declines in the ***Mitotic Fraction***, ***m***, is well fit by the ***Universal Mitotic Fraction Equation***, $m = a^{(N_w^b)}$

The basis for the relationship between the ***Mitotic Fraction***, ***m***, and $N_w$ becomes evident by graphing the $\log(N_w)$ vs$-\log(-\log(m))$, revealing roughly straight rows of dots (FIGURE A4d). This suggests that the relationship between the ***Mitotic Fraction***, ***m***, and $N_w$ can be captured with the expression:

$$m = a^{(N_w^b)} \tag{1}$$

The best fit values for ***a*** and ***b*** for each species were calculated as regression inputs of $\log N_i$ and $-\ln(-\ln g_i)$ using Excel's LINEST function. Values for ***a*** and ***b*** were calculated from the LINEST output (in columns P-Q of the excel files listed in Table 1) together with the $r^2$ value. This relationship is well borne out, as indicated by the high $r^2$ values from non-linear regressions for each of the eleven animals listed in the Table 2.

Because Equation #1 so closely captures the relationship between the ***Mitotic Fraction***, ***m***, and size, $N_w$, from fertilization until maturity, for all of the animals we have examined, we call it the "***Universal Mitotic Fraction Equation***". ("Universal"; from the Latin "universalis"; Etymology: "uni (Latin, "one"), Versum (Latin, "turned"); thus something "turned into one"). Note that the ***Universal Mitotic Fraction Equation***, and the ***Universal Growth Equation***, that is derived from it, and which we shall describe below, holds for all of the animals that we have examined, and for all of the sizes we have examined, from the first fertilized cell onward.

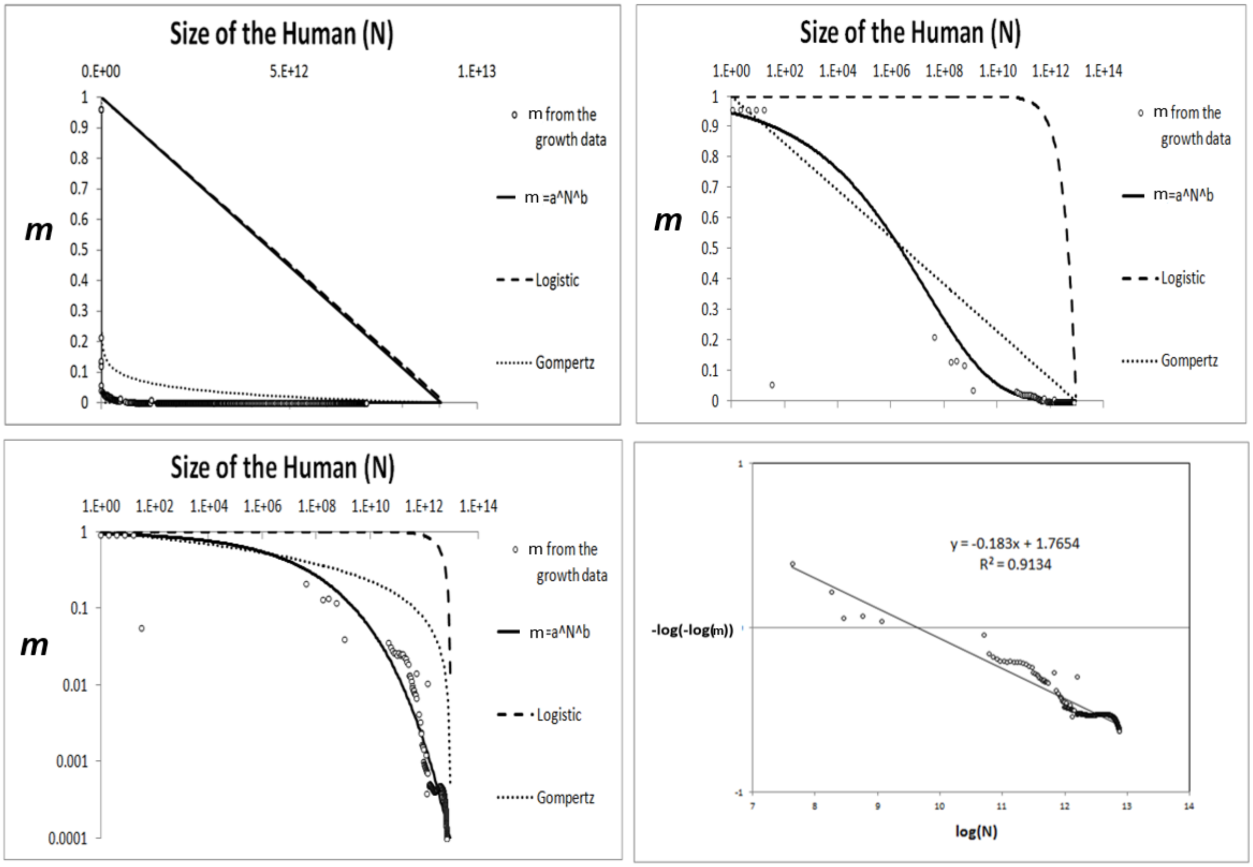


**FIGURE A4**: **Decline in fraction of cells dividing, the *Mitotic Fraction, m*, calculated by the *Mitotic Fraction Method*, that occurs as animals increase in size, as seen by the *Universal Mitotic Fraction Equation*.** Data points for size, $N_w$, in integer units of numbers of cells, vs. the ***Mitotic Fraction***, ***m***, from fertilization, until maturity, for humans.

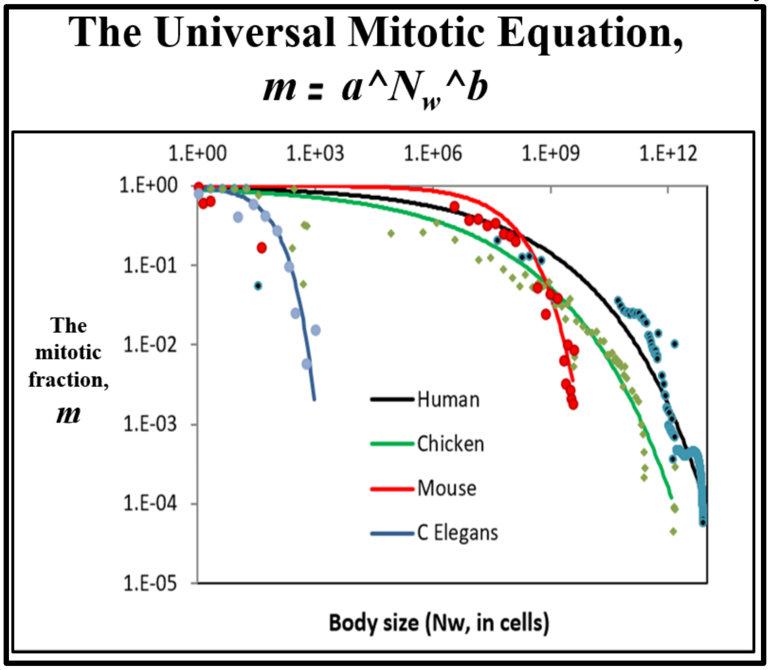


**FIGURE A5**: **Values of the *Mitotic Fraction, m*, calculated by the *Mitotic Fraction Method*, as a function of animal size, $N_w$, as shown at various scales.** Data points for animal size, $N_w$, in integer units of numbers of cells, vs. the ***Mitotic Fraction***, ***m***, from fertilization, until maturity, for humans, chickens, ***C elegans*** nematode worms, and mice.

The relationship between size, $N_w$, and age, $t$, is well fit by the ***Universal Growth Equation***: $\int \left(\frac{\log(2)}{c}\right) N_w a^{(N_w^b)}$

Combining Equation #3 and #4 leads to this expression for describing the rate of animal growth:

$$\frac{dN_w}{dt} = \frac{ln(2)}{c} \cdot N_w \cdot a^{(N_w^b)} \quad (5a)$$

The relationship between size, $N_w$, and age, $t$, is captured by numerical integration of Equation #5, where the age, $t$, at fertilization is ∼0, and the age at death is $d$:

$$N_w = \int_{t=0}^{t=d} \frac{ln(2)}{c} \cdot N_w \cdot a^{(N_w^b)} \quad (5b)$$

Numerical integration of Equation #5, to derive Equation #5b, was carried out 4th order Runge-Kutta method, employed manually in Excel to carry out such growth curve reconstructions, so as to examine relationship between Size ($N_w$) and Age ($t$). The step size $h$ (cell b11 of the excel files listed in "**Calculations Files**" column of the excel files enumerated in Table 1) was set such that 10,000 steps was used for each organism. At each $t_i$ (column W), the model value of $N_i$ was calculated recursively in column X as

$$N_i = N_{i-1} + \frac{h(k_1+2k_2+2k_3+k_4)}{6} \quad (200)$$

where:

$$\text{if: } f(N) = N' = rNg \quad (201)$$

$$k_1 = f(N_i) \quad (202)$$

$$k_2 = f\left(N_{i-1} + \frac{k_1}{2}h\right) \quad (203)$$

$$k_3 = f\left(N_{i-1} + \frac{k_2}{2}h\right) \quad (204)$$

$$k_4 = f(N_{i-1} + k_3 h) \quad (205)$$

The values of each $k_i$ are calculated in columns Z-AC of the excel files listed in "**Calculations Files**" column of the table above. Excel's VLOOKUP and CORREL functions were then used to calculate correlations of observed and model **ln *g*** and **ln *N*** (columns AM-AU). These correlations, as well as the regression correlation coefficient, were used as quality-of-fit metrics for the model and recorded in the summary columns A-B in the excel files.

The fit of Equation #5b, the numerically integrated form of Equation #5, to the various datapoints for animal size's ($N_w$) versus time ($t$), are shown in FIGURE A1. The strength of this relationship was confirmed by the high $r^2$ values for each of the eleven animals listed in the Table 2: humans, frogs, nematodes, chickens, cows, geese, mice, quail, rats, turkeys, fish, and clams. Because Equation #5, in its numerically integrated form (Equation (#5b)) closely captures the relationship between size, $N_w$, and age, $t$, from fertilization until maturity, for all of the animals we have examined, we call it the "***Universal Growth Equation***". We call the biological process which the ***Universal Mitotic Fraction*** and ***Universal Growth Equations*** capture "***UNI-GROWTH***".

Closed form solution to the integration of the ***Universal Growth Equation***

A closed form solution to the integration of the ***Universal Growth Equation*** (#5), can also be seen by considering:

$$\frac{dN}{dt} = r\,N\,a^{(N^b)} \tag{301}$$

where $r = \frac{\log(2)}{c}$. Rearranging, one obtains:

$$N^{-1}\,a^{(-N^b)}\,dN = r\,dt \tag{302}$$

which can be integrated as:

$$\int_1^{N_w} N^{-1}\,a^{(-N^b)}\,dN = \int_0^t r\,d\tau \tag{303}$$

Leading to the growth equation:

$$t = \left[\mathrm{Ei}\left(-N_w^b \log(a)\right) - \mathrm{Ei}(-\log(a))\right] \frac{c}{b\,\log(2)} \tag{3}$$

where $t = 0$ when $N_w = 0$, and **Ei** is the exponential integral function[63].

Equation #3 can be considered as a continuous approximation of the expected time required by a Poisson model with individual cell growth $ra^{(N^b)}$.

We show this by considering the random variable $t_N$ representing the time required to reach the number of cells *N*, starting from a single cell. This variable has expected value:

$$E[t_N] = \sum_{n=2}^{N} E[t_n - t_{n-1}] \tag{305}$$

We assume that each one of the *n* cells that make up the organism at a given time behaves according to a Poisson process with rate $ra^{(n^b)}$ and that they are all independent. As a result, the event represented by the growth of a new cell out of any of the *n* current cells behaves according to a Poisson process with rate $n\,r\,a^{(n^b)}$ (this can be easily seen by considering the cumulative distribution function of the exponential distribution). Therefore:

$$\overline{t_N} = E[t_N] = \sum_{n=2}^{N} \frac{1}{nra^{(n^b)}} \tag{306}$$

$$\sigma_N = \sqrt{\mathrm{var}(t_N)} = \sum_{n=2}^{N} \frac{1}{\left(nra^{(n^b)}\right)^2} \tag{307}$$

where $\overline{t_N}$ and $\sigma_N$ are, respectively, the mean and standard deviation of the time required to reach the number of cells *N*. The continuous version in #6 and the discrete (point-process) version in #306 provide very similar results. For example, using $c = 0.070$, $a = 0.925$, $b = 0.269$, we get the behavior shown in FIGURE A3 for the continuous version in red, and the discrete version in blue (mean ± std) (FIGURE A3).

Density dependent growth equations

A number of ***density-dependent*** growth equations have been developed to capture either the growth of the number of organisms in a population,[64,65] or the growth of the size of individual organisms,[7-11] none of which accurately fit the growth of real animals.[12-14] The simplest of these ***density-dependent*** growth equations, the logistic, occurs by $\boldsymbol{m}$ declining linearly as the size of the organism, $\boldsymbol{N_w}$, increases.[7] In the ***Gompertz equation***[8], $\boldsymbol{m}$ declines as the log of $\boldsymbol{N_w}$. Similarly, for many other equations, including the ***von Bertalanffy***[9], the ***Richards***[10], and ***West***[11] equations, the value of $\boldsymbol{m}$ declines monotonically as size, $\boldsymbol{N_w}$, increases.

The relationship of the ***Mitotic Fraction*** (***m***) and size ($\boldsymbol{N_w}$) to the ***logistic equation***.

***Density dependent*** growth is captured by the ***logistic equation***[7] in the form:

$$\boldsymbol{m = 1 - \frac{N_w}{K}} \tag{401}$$

The relationship of the ***Mitotic Fraction*** (***m***) and size ($\boldsymbol{N_w}$) to the ***Gompertz equation***.

***Density dependent*** growth is captured by the ***Gompertz equation***,[8] in the form:

$$\boldsymbol{m = \left(\frac{1}{\log K}\right) \log\left(\frac{N_w}{K}\right)} \tag{402}$$

The relationship of the ***Mitotic Fraction*** (***m***) and size ($\boldsymbol{N_w}$) to the ***West equation***.

***Density dependent*** growth is captured by the ***West equation***,[11] for total mass of the organism (***M***), in the form

$$\boldsymbol{\frac{dM}{dt} = aM^{\frac{3}{4}}\left(1 - \left(\frac{M}{K}\right)^{\frac{3}{4}}\right)} \tag{403}$$

Units of mass are unspecified, so we can treat Equation #403 such that the units of mass (***M***) of our organism are in units of "the weight of a cell" ($\boldsymbol{N_w}$). West's equation then becomes:

$$\boldsymbol{\frac{dN_w}{dt} = aN^{\frac{3}{4}}\left(1 - \left(\frac{N_w}{k}\right)^{\frac{3}{4}}\right)} \tag{404}$$

By re-arranging:

$$\boldsymbol{\frac{dN_w}{dt} = \left[\left(\frac{\log 2}{c}\right) N\right]\left[\left(\frac{ac}{\log 2}\right) N^{-\frac{3}{4}}\right]\left[1 - \left(\frac{N_w}{k}\right)^{\frac{3}{4}}\right]} \tag{405}$$

Thus, for any organism growing by the ***West equation***, the fraction of cells dividing, ***m***:

$$m = \left[\left(\frac{ac}{\log 2}\right) N_W^{-\frac{3}{4}}\right]\left[1 - \left(\frac{N_W}{k}\right)^{\frac{3}{4}}\right] \tag{406}$$

Equation #406 can be thought of as the ***West equation***, phrased in terms of the fraction of cells dividing, ***m***.

Fit of the relationship of the ***Mitotic Fraction*** (***m***) and size ($\mathbf{N}_\mathrm{W}$), to the ***Gompertz***, ***logistic***, and ***West equations***.

As can be seen in FIGURE A4, over the full range from conception to adult size, neither the Gompertz (Equation #402), nor the logistic (Equation #401) provides a close characterization of the behavior of the ***Mitotic Fraction***, ***m***, with respect to $\boldsymbol{N_w}$. As can be seen in FIGURE A6, while the ***West equation*** can be used to quite accurately capture growth over its full expanse, to do so requires $\boldsymbol{m > 1}$ early in development, which is mathematically valid, but biologically impossible, since individual cells would have to give rise to more than two daughter cells in a single ***Cell Cycle Time***. However, both our ***Universal Growth Equation*** (Equation #5), and the ***West equation*** (#406), work quite well (FIGURE A6) later in development. This means that our ***Universal Growth Equation*** is capable of achieving growth within the limits of the blood supply's capacity to support growth, as captured by the ***West equation***.[11]

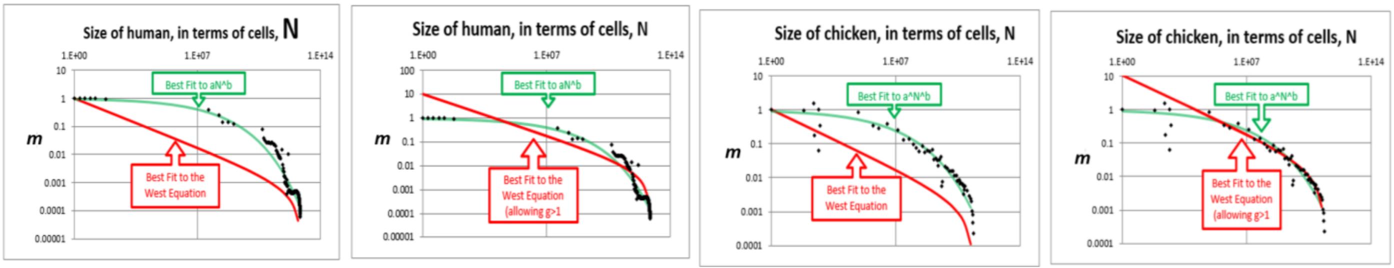


**FIGURE A6**: **Comparison of the fit of *Mitotic Fraction*, *m*, as a function of size, $\boldsymbol{N_w}$, with human growth data, to the *Universal Mitotic Fraction Equation* (green) and *West equation* (red).**

Fit of the relationship between size ($N_w$) and Age ($t$) to the ***Gompertz***, ***logistic***, and ***West equations***.

We tested our data on body size, $N_w$, and age, $t$, against three of the most widely used density dependent growth equations (the ***Gompertz***, the ***logistic***, and the ***West*** [a variation of the ***von Bertalanffy***]) (FIGURE A2). None of these growth equations fit the actual growth data as well as the ***Universal Growth Equation*** (FIGURE A2).

Fine scale "***ripples***" identify a lower level of growth detail that occurs in the life of the organism.

Of course, no equation, however closely it may summarize actual data, can ever capture the world precisely. In fact, while the values for body size, $N_w$, and age, $t$, that we have assembled, fall remarkably close to the ***Universal Growth Equation***, we can also see hints of fine-scale minor "***ripples***" departing from the smooth edge of the curve (FIGURE A1 and A2). We suspect that these ***ripples*** identify specific events in the life of the organism, such as gastrulation, somite formation, weaning, puberty, and individual differences, that are associated with sex, malnutrition, genetic variation, and so on. To extract this second layer of growth information, we derived an expression, Equation #500, to measure the magnitude and timing of these growth residuals, $\boldsymbol{\delta_j}$:

$$m = A^{N^B} + \sum\delta_j \tag{500}$$

where each $\delta_j$ models a wave-like deviation away from the trend (the ***Universal Growth Equation*** #5b) that corresponds to an interpretable life-event in the growth of the organism. The appropriate functional forms for the $\delta_j$ may be determined through regression of the residuals of observed ***m*** against the ***Universal Growth Equation*** (#5b).

Indeed, a graph of these $\delta_j$ values, as a function of the size of the developing human male, $N_w$, which is shown in FIGURE A7 bears out our suspicion that these ***ripples*** identify biologically relevant processes, such as the right-most peak (at $\log N_w \approx 29.3$), which corresponds to puberty.

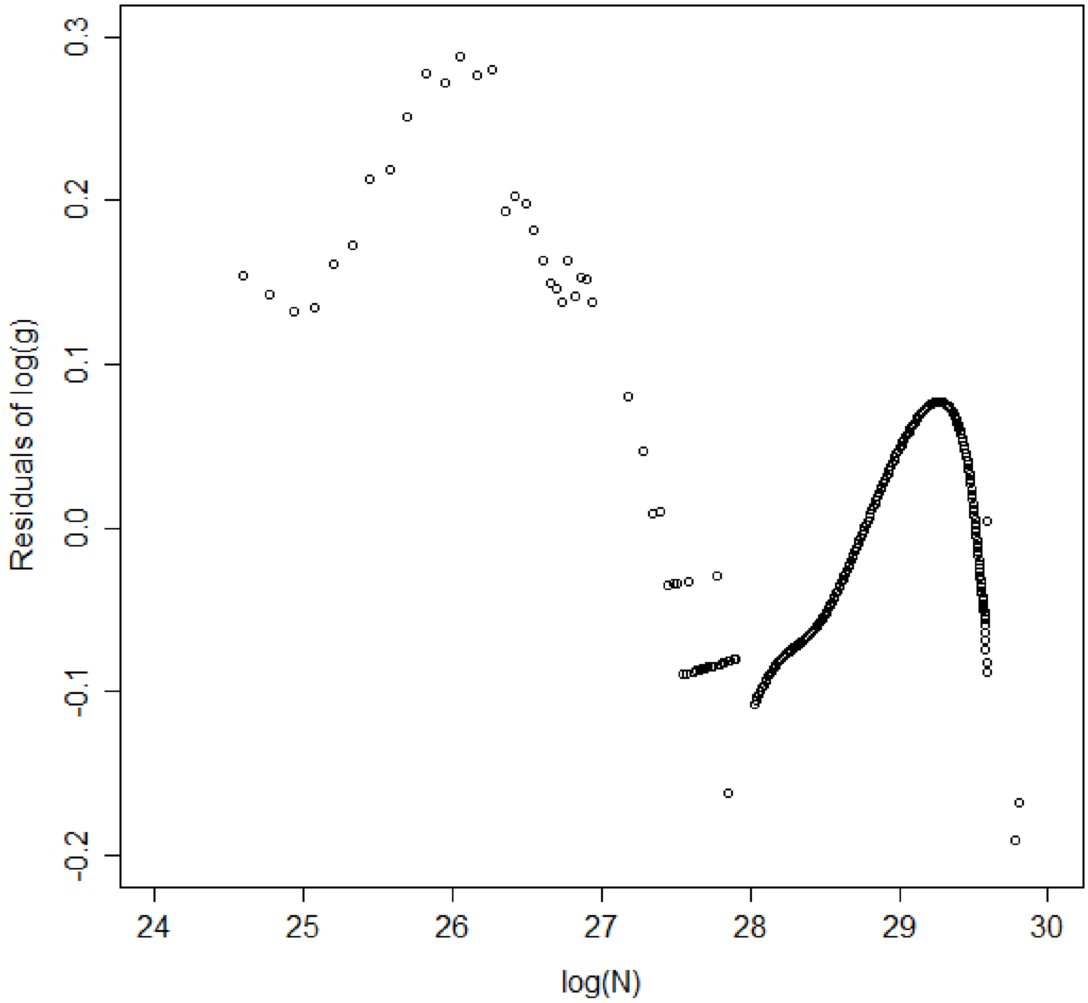


**<u>FIGURE A7</u>: Wave-like deviations ("*ripples*") from the *Universal Growth Equation curve*, for human growth.**

The Meaning of ***UNI-GROWTH***

The growth data shown in Tables 1 and 2, and FIGURE A1-A6 make clear that animals as diverse as mollusks, nematodes, fish, amphibians, birds, and mammals, including humans, all grow by the ***Universal Growth Equation*** (#5). The values of ***a***, ***b***, ***c***, and ***R*** make the ***Universal Growth Equation*** (#5) specific for each individual. Let us unpack the biological meanings of the form of the ***Universal Growth Equation*** and its parameters.

The ***Universal Growth Equation*** describes how growth occurs

Mathematically, each of the parameters of the ***Universal Growth Equation*** (#5) has a specific impact on the shape of the "***S-shaped***" growth curve. The ***a*** and ***b*** parameters determine the curviness of the "***S***". The ***c*** parameter, the ***Cell Cycle Time***, determines the speed of growth, expanding or contracting the "***S***" like an accordion.

The ***Universal Growth Equation*** is linked to the cell biology behind growth.

Biologically, each of the parameters of the ***Universal Growth Equation*** (#5) is linked to a specific aspect of cell division. The ***Mitotic Fraction***, ***m***, reflects how many cells are engaged in mitosis; ***m*** itself, which decreases as we become larger, is determined by the ***a*** and ***b*** parameters of ***Universal Mitotic Fraction Equation*** (#4), which lies within the ***Universal Growth Equation*** (#5). The ***c*** parameter, the ***Cell Cycle Time***, of the ***Universal Growth Equation*** (#5) describes how fast it takes cells to divide.

The ***Universal Growth Equation's*** description of how fast we grow is linked to the ***Cell Cycle Time***, ***c***, which is linked to genome size, which is linked to how much junk DNA we have.

What might be the mechanism behind the ***Cell Cycle Time***, the ***c*** parameter of the ***Universal Growth Equation*** (#5), which not only determines how long it takes a cell to divide, but also determines the speed of the growth of the body as a whole? Biochemically, the principal determinant of the ***Cell Cycle Time***, ***c***, is the amount of DNA that the cell contains. The more DNA a cell has to copy, the longer it takes to divide. This was first found by Van't Hof and Sparrow, who discovered a linear relationship between the total amount of amount of DNA and ***Cell Cycle Time*** for plant cells.[66] Their observation has subsequently been confirmed in many studies of many types of organisms.[67,68] For most animals, the largest part of the genome is non-coding DNA, sometimes called junk DNA,[69] no doubt imprecisely named.[70,71] The amount of this non-coding DNA has been found to be correlated with growth in salamanders,[72] anurans,[73] amphibians,[74] insects,[75] and copepods.[76]

The speed of growth has a profound impact on survival.[77,78] Human fetuses that grow to full-size in 10 months, or in 8 months, have a much lower chance of survival than fetuses that reach optimal size in 9 months.[79] Non-coding DNA, which determines genome size, is very susceptible to duplication or deletion, and thus provides populations of animals abundant genetic variation in genome size.[80-82] Perhaps this genetic variation in genome size leads to genetic variation in the ***Cell Cycle Time***, ***c***, which leads to genetic variation in the speed of body growth. Such genetic variation would appear to give species a powerful resource to draw upon, so that they can evolve, by the bitter reality of Darwinian selection, to growth rates that give them the greatest chance of survival. Indeed, this makes us wonder whether junk DNA may owe its very existence to its role in determining the speed of growth.

The ***Universal Growth Equation's*** description of the fraction of cells dividing, the ***Mitotic Fraction***, ***m***, is linked to the action of cell signaling molecules that induce mitotic quiescence.

What might be the mechanism behind the ***Mitotic Fraction***, ***m***, the fraction of cells that divide, which declines as we increase in size? What could cause a fraction of the body's cells to enter into a state of ***mitotic quiescence***, the internal block to cell division, which is set in motion by external signaling molecules?[60-62] Curiously, as we shall see below, this decline in the ***Mitotic Fraction***, by the ***Universal Mitotic Fraction Equation*** (#4), $\boldsymbol{m = a^{(N_w^b)}}$, might well trace its origin to the deceptively simple and unobvious discrete allocation of ligand molecule among cells[83]. To picture how this might occur, consider the case of an embryo growing in a constant volume, such as a bird's egg or mammal's uterus, whose cells produce an inhibitory growth factor and carry a receptor for that inhibitory molecule. Early in development, when our idealized embryo is just a few cells, those few cells would produce only a small number of inhibitory molecules, and thus the concentration of these inhibitory molecules would be low, and thus very few cells would have bound the number of inhibitory molecules needed to prevent cell division. Thus, at the beginning of development, most cells would divide, with the value of the ***Mitotic Fraction***, ***m***, being close to **1**. However, as the embryo grows, more and more cells are present to produce inhibitory molecules, the concentration of these inhibitory molecules would increase, more and more cells would have bound the number of inhibitory molecules needed to prevent cell division, fewer and fewer cells would be able to divide, and the value of the ***Mitotic Fraction***, ***m***, would decline. Remarkably, as we shall see next, when we rephrase this idealized case in mathematical terms, such a decline in the ***Mitotic Fraction***, ***m***, can be seen to occur in exactly the form of the ***Universal Mitotic Fraction Equation*** (#4), $\boldsymbol{m = a^{(N_w^b)}}$ .

Let us consider growth factor molecules in terms of integers.[83] Thus, when growth factor molecules bind to cells, they shall display a discrete allocation among cells, that is, as a Poisson process, which can be comprehended in terms of a Poisson probability (Equation #611 below).

Let **Ic** be the number of inhibitory growth factor molecule made by each cell which makes the molecule. (This number is a constant, being the result of the rates of synthesis and decay, most likely being first order reactions.)

Let $\boldsymbol{N}$ be the number of cells in the organism. We shall consider growth from conception, that is, from when $\boldsymbol{N = 1}$.

Let $\boldsymbol{N^b}$ be the number of cells producing the inhibitory growth factor molecules. If every cell produces the molecules, $\boldsymbol{b = 1}$. If only the cells on the surface produce the molecule, $\boldsymbol{b = \frac{2}{3}}$, since this captures the relationship between the surface of an isomorphic 3-dimensional shape, such as a sphere, and its volume.

Let $\boldsymbol{V}$ be the volume in which the embryo develops. (For a bird $\boldsymbol{V}$ is the volume of the egg; for a mammal it is the volume of the uterus.)

The total number of molecules of **I** in the body is $\boldsymbol{Ic \cdot N^b}$**.**

**[I]** is the concentration of inhibitory growth factor molecule in the body, assuming uniform diffusion.

Let us now put all this into mathematical form.

It follows that:

$$[\mathbf{I}] = \frac{\boldsymbol{Ic \cdot N^b}}{\boldsymbol{V}} \tag{601}$$

$$[\mathbf{I}] = \left(\boldsymbol{Ic \cdot N^b}\right)\left(\frac{\mathbf{1}}{\boldsymbol{V}}\right) \tag{602}$$

Let $\mathbf{P_i}$ be the probability of a receptor molecule binding an inhibitory growth factor molecule, where **[R]** is the concentration of receptor molecules.

$$\mathbf{P_i} = \frac{[\mathbf{IR}]}{[\mathbf{R}]} \tag{603}$$

$$\frac{[\mathbf{I}][\mathbf{R}]}{[\mathbf{IR}]} = \mathbf{k} \tag{604}$$

according to the Law of Mass Action, thus

$$\frac{[\mathbf{IR}]}{[\mathbf{R}]} = \frac{[\mathbf{I}]}{\mathbf{k}} \tag{605}$$

$$\mathbf{P_i} = [\mathbf{I}] \cdot \frac{\mathbf{1}}{\mathbf{k}} \tag{606}$$

$$\mathbf{P_i} = \left(\mathbf{Ic} \cdot \mathbf{N^b}\right)\left(\frac{\mathbf{1}}{\mathbf{V}}\right)\left(\frac{\mathbf{1}}{\mathbf{k}}\right) \tag{607}$$

$$\mathbf{P_i} = \mathbf{N^b}(\mathbf{Ic})\left(\frac{\mathbf{1}}{\mathbf{V}}\right)\left(\frac{\mathbf{1}}{\mathbf{k}}\right) \tag{608}$$

$$\mathbf{P_i} = \mathbf{N^b}\left(\frac{\mathbf{Ic}}{\mathbf{V \cdot k}}\right) \tag{609}$$

$$\mathbf{P_i} = \left(\frac{\mathbf{Ic}}{\mathbf{V \cdot k}}\right)\mathbf{N^b} \tag{610}$$

Let **Ro** be the number of receptors per cell.

We shall use the Poisson probability[84] to calculate the value of $\boldsymbol{m}$:

$$\mathbf{P}(\mathbf{x};\boldsymbol{\mu}) = \frac{(\mathbf{e^{-\mu}})(\boldsymbol{\mu}^{\mathbf{x}})}{\mathbf{x}!} \tag{611}$$

where **x** is the actual number of successes that result from the experiment.

**e**: A constant equal to approximately 2.71828

**μ**: The mean number of successes that occur in a specified region.

**x**: The actual number of successes that occur in a specified region.

$\mathbf{P}(\mathbf{x};\boldsymbol{\mu})$: The Poisson probability that exactly **x** successes occur in a Poisson experiment, when the mean number of successes is **μ**.

It follows that

$$\boldsymbol{m} = \mathbf{P}(\mathbf{x};\mathbf{u}) = \frac{(\mathbf{e^{-\mu}})(\boldsymbol{\mu}^{\mathbf{x}})}{\mathbf{x}!} \tag{612}$$

It follows that $\boldsymbol{x = 0}$ (since we want to calculate the chance of a cell binding **0** molecules of inhibitory growth factor).

And where $\boldsymbol{\mu} = \mathbf{Ro} \cdot \mathbf{P_i}$ (since the mean number of successes for each cell is the number of receptors, time the chance that a molecule will bind one of those receptors):

$$\boldsymbol{m} = \mathbf{P}(\mathbf{x};\boldsymbol{\mu}) = \frac{(\mathbf{e^{-\mu}})(\boldsymbol{\mu}^{\mathbf{0}})}{\mathbf{0}!} \tag{613}$$

$$m = \mathbf{P}(\mathbf{x}; \boldsymbol{\mu}) = \frac{(\mathbf{e}^{-\boldsymbol{\mu}})(\mathbf{1})}{\mathbf{1}} \quad (614)$$

$$m = \mathbf{P}(\mathbf{x}; \boldsymbol{\mu}) = \mathbf{e}^{-\boldsymbol{\mu}} \quad (615)$$

$$m = \mathbf{P}(\mathbf{x}; \boldsymbol{\mu}) = \mathbf{e}^{-\mathbf{Ro}\cdot\mathbf{P_i}} \quad (616)$$

or

$$m = \mathbf{e}^{-\mathbf{RoP_i}} \quad (617)$$

It follows that

$$m = \mathbf{e}^{-\mathbf{Ro}\left(\frac{\mathbf{Ic}}{\mathbf{V\cdot k}}\right)(\mathbf{N^b})} \quad (618)$$

Let $Z = Ro\left(\frac{Ic}{V\cdot \mathbf{k}}\right)$

$$m = \mathbf{e}^{-\mathbf{Z}(\mathbf{N^b})} \quad (619)$$

Let $a = e^{-Z}$

$$m = a^{(\mathbf{N^b})} \quad (4)$$

Which is the ***Universal Mitotic Fraction Equation***!

This leads to our ***Universal Growth Equation***, when combined with Equation #5a:

$$\frac{dN}{dt} = \frac{\ln(2)}{c} \cdot N \cdot a^{(N^b)} \quad (5a)$$

where

$$a = \ln\left(Ro\left(\frac{Ic}{V\cdot \mathbf{k}}\right)\right) \quad (620)$$

Note that all of the biochemistry (the rate of growth factor synthesis, abundance of receptors, dissociation constants, etc.) is in the parameter ***a***, while all of the geometry (size of the tissues that making the growth factor in relationship to the size of the organism) is in the parameter ***b*** (see "Let $N^b$ be …" above).

Having worked through Equation #601-620, let us step back and review the core implications of this math. This math is telling us that little more than the discrete allocation of mitotic signaling molecules among cells is all that is needed to explain how cell division becomes limited to a faction of cells, ***m***, the ***Mitotic Fraction***, as captured by the ***Universal Mitotic Fraction Equation*** (#2), and its two parameters ***a*** and ***b***, and from this, to cause our growth to occur by the ***Universal Growth Equation***. These mitotic signaling molecules, the receptors on the cells to which they bind, and the internal mitotic control molecules that respond to these signals, are all ruled by ordinary chemistry. Thus, little more than the conventional thermodynamics of all chemical reactions is sufficient to account for growth by the ***Universal Growth Equation***, and thus the biological process of ***UNI-GROWTH***. Let us also recall that since signaling molecules, their receptors, and the internal mitotic control molecules that determine whether cells divide, are the products of our genes, and thus genetic polymorphism in the value of ***a*** or ***b*** might well be expected to identify genetic polymorphisms of the proteins that do this mitotic signaling, a possibility that would certainly be worthy of experimental examination.

***UNI-GROWTH***: Summary Definition

Let us summarize: ***UNI-GROWTH*** is the process by which growth slows as we increase in size. ***UNI-GROWTH*** is captured by the ***Universal Mitotic Fraction*** and ***Growth Equations***, and their parameters, ***a***, ***b***, and ***c***, the average ***Cell Cycle Time***. ***UNI-GROWTH*** occurs by the decline in the fraction of cells dividing, the ***Mitotic Fraction***, occurring by the form of ***Universal Mitotic Fraction Equation***, ***m=a^(N^b)***. From this expression, we could then derive, and test, the ***Universal Growth Equation***, whose accuracy was confirmed, from fertilization until maturity, for 13 species, including nematodes, mollusks, amphibians, fish, birds, rodents, cows, and humans.

#### ***ALLO-GROWTH***

The ***Cellular Phylodynamic Analysis*** of the growth and development of the parts of the body

The ***Allometric Growth Equation***

For more than a century, biologists have been studying relative growth, that is, the growth of one part of the body, $\boldsymbol{p}$, against another part, $\boldsymbol{p_2}$, or against the body as a whole, $\boldsymbol{w}$.[15,18] These studies of relative growth, usually carried out in units of weight, volume, or length, had found that the relationships between $\boldsymbol{p}$ and $\boldsymbol{w}$ often form straight rows of dots on log-log graphs.[15-19] This log-linearity of relative growth, called ***allometric growth***[85], can be captured by:

$$\log(\boldsymbol{w}) = \frac{\boldsymbol{1}}{\boldsymbol{S}} \cdot \log(\boldsymbol{p}) + \log(\boldsymbol{B}) \tag{630}$$

which is equivalent to:

$$\boldsymbol{w} = \boldsymbol{B} \cdot \boldsymbol{p}^{\frac{\boldsymbol{1}}{\boldsymbol{S}}} \tag{631}$$

Here, we shall call $\boldsymbol{B}$ the "***allometric birth***" and $\boldsymbol{S}$ the "***allometric slope***". Below, it will become clear why we have named the parameters in this fashion, as their meanings emerge when relative growth is framed in terms of numbers of cells, $\boldsymbol{N}$.

While the empirical validity of the log-linearity of allometric growth has been evident from countless studies of the growth of many parts of the body, in many animals,[15-19] the meaning of this phenomenon has long been a mystery, as have been the biological meanings of the parameters $\boldsymbol{B}$ and $\boldsymbol{S}$.[19,86,87] However, as we shall outline next, when we frame the relationship of the growth of one part of the body, $\boldsymbol{p}$, against the body as a whole, $\boldsymbol{w}$, in terms of the number of cells in the part of the body, $\boldsymbol{N_p}$, against the number of cells in the body as a whole, $\boldsymbol{N_w}$, the biological basis and significance of the log-linear feature of allometric growth becomes evident, together with the biological meanings of the two parameters that define it, $\boldsymbol{B}$ and $\boldsymbol{S}$.

***Cellular Allometric Growth Analysis***

The approach that we have taken to deciphering the nature of the relative growth of each part of the body is the same as the approach we described above for examining the growth of the body as a whole: ***Cellular Phylodynamic Analysis***, by which we again mean examining growth in terms of integer numbers of cells, $\boldsymbol{N}$. To do so, we examined the sizes of the various parts of the body, $\boldsymbol{p}$, in these integer units of numbers of cells, $\boldsymbol{N_p}$, in comparison to the numbers of cells in the body as a whole, $\boldsymbol{w}$, also in these integer units of numbers of cells, $\boldsymbol{N_w}$, an approach we call "***Cellular Allometric Growth Analysis***" (FIGURES A8-A17). Graphically, such a ***Cellular Allometric Growth Analysis*** of relative growth is carried out by examining paired $\boldsymbol{N_p}$:$\boldsymbol{N_w}$ values on log-log graphs (FIGURES A8-A17).

When we have a full lineage chart for an animal, which characterizes each cell from the first fertilized egg onward, we call this "***Cellular Allometric Lineage Growth Analysis.***" When we are carrying this out without lineage data, but simply with cell numbers, whether these cell numbers have been counted, or estimated from weight values, based on the finding that there are about $10^8$ cells in every gram of tissue,[57] we call this approach "***Cellular Allometric Batch Growth Analysis.***"

Early development: ***Cell lineages***: ***Cellular Allometric Lineage Growth Analysis***:

To carry out a ***Cellular Phylodynamic Analysis*** of the relative growth of the various parts of the body early in development, we assembled values of $\boldsymbol{N_p}$ vs. $\boldsymbol{N_w}$ for each of the ***cell lineages*** (FIGURE A8) that have been characterized in the ***cell lineage charts*** of ***Caenorhabditis elegans***[20] and ***Meloidogyne incognita***[21] nematode worms, as well as for ***Oikopleura dioica*** chordate tunicates (FIGURES A9-A11).[88] To accomplish such ***Cellular Allometric Lineage Growth Analysis***, we simply counted the number of cells in each ***lineage***, $\boldsymbol{N_p}$, from the first ***Founder Cell*** onward ($\boldsymbol{N_p} = 1$), together with the corresponding values for the number of cells in the embryo as a whole, $\boldsymbol{N_w}$ (FIGURES A9-A11).

Such a ***Cellular Allometric Lineage Growth Analysis*** was easily carried out for the ***M incognita***, whose ***cell lineage chart*** fits on a single sheet of paper (FIGURES A9). However, ***Cellular Allometric Lineage Growth Analysis*** was considerably more difficult for ***C elegans*** and ***O dioica***, whose ***cell lineage charts*** have to be 6 feet long to see each cell, requiring many months of work to assemble the values of $\boldsymbol{N_p}$ and $\boldsymbol{N_w}$ shown in FIGURES A10 AND A11. This motivated the development of a new technique, which we shall describe below, the ***BinaryCellName Method***, for carrying out such ***Cellular Allometric Lineage Growth Analysis*** computationally.

Late development: ***Tissues***, ***Organs***, and ***Anatomical structures***: ***Cellular Allometric Batch Growth Analysis***:

To carry out a ***Cellular Phylodynamic Analysis*** of the relative growth of the various parts of the body later in development, that is, ***Cellular Allometric Batch Growth Analysis***, we assembled values of $\boldsymbol{N_p}$ vs. $\boldsymbol{N_w}$ for the ***tissues***, ***organs***, and ***anatomical structures*** evident in embryos and juveniles from published weight and volume values, again relying on the observation that there are about $10^8$ cells in every gram of tissue.[57] We assembled these values for the body parts of chicks (gizzards, livers, hearts, kidneys),[89] rats (livers, brains, kidneys, forelegs, ears, stomachs, spinal cords),[90] humans (brains, livers, kidneys, lungs, pancreases, adrenals, thymuses, spleens, lower extremities, upper extremities, stomach, heart, intestines),[91] zebrafish (eye lens),[92-95] mice (livers, brains, kidneys, forelegs),[90,96] clams and goldfish (FIGURES A12-A17).

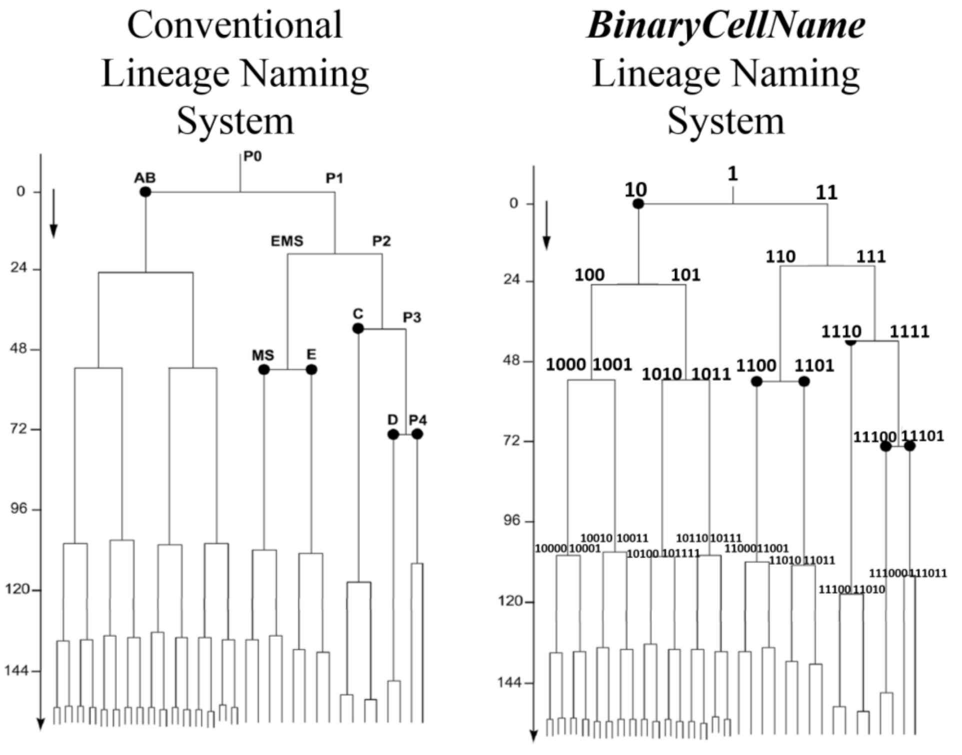


**<u>FIGURE A8</u>: *Cell lineage chart* of the *cell lineages* of the developing root knot nematode, *Meloidogyne incognita*. Shown are the conventional and *BinaryCellName Lineage* naming methods. The AB *lineage* of nematodes forms the skin while the E *lineage* forms the intestine. *Cell lineage chart*** and data from Calderón-Urrea et al[21]

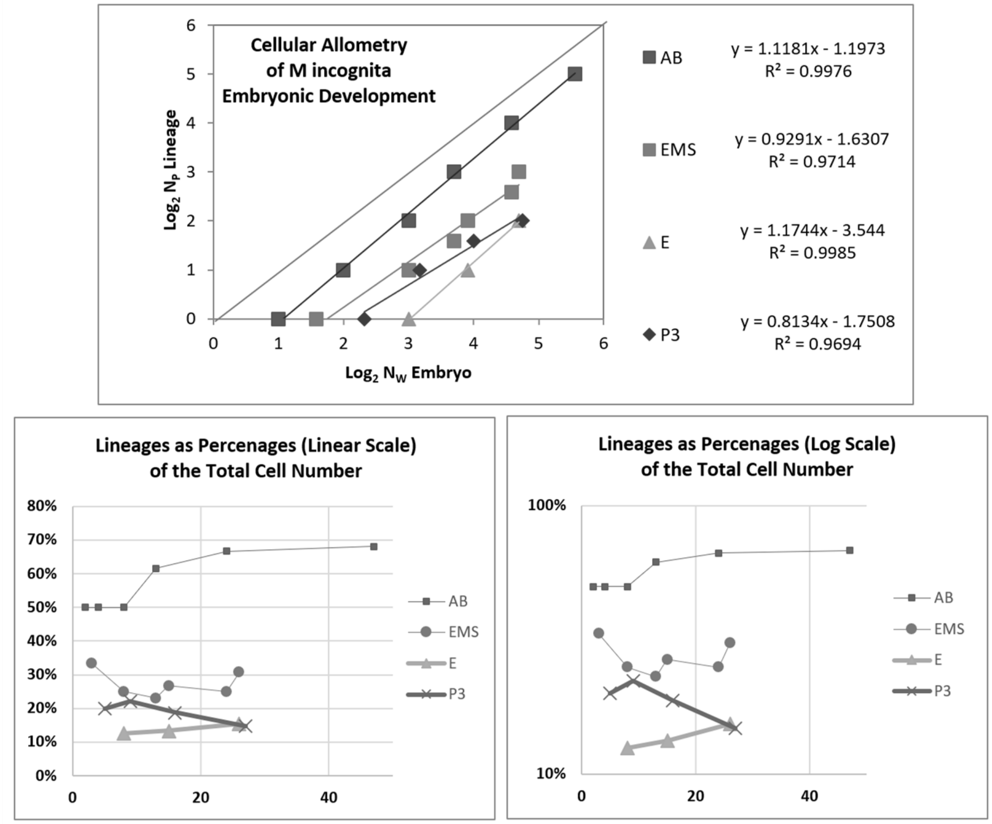


**<u>FIGURE A9</u>: *Cellular Allometric Lineage Growth Analysis* of the *cell lineages* of the developing root knot nematode, *Meloidogyne incognita***.[21]

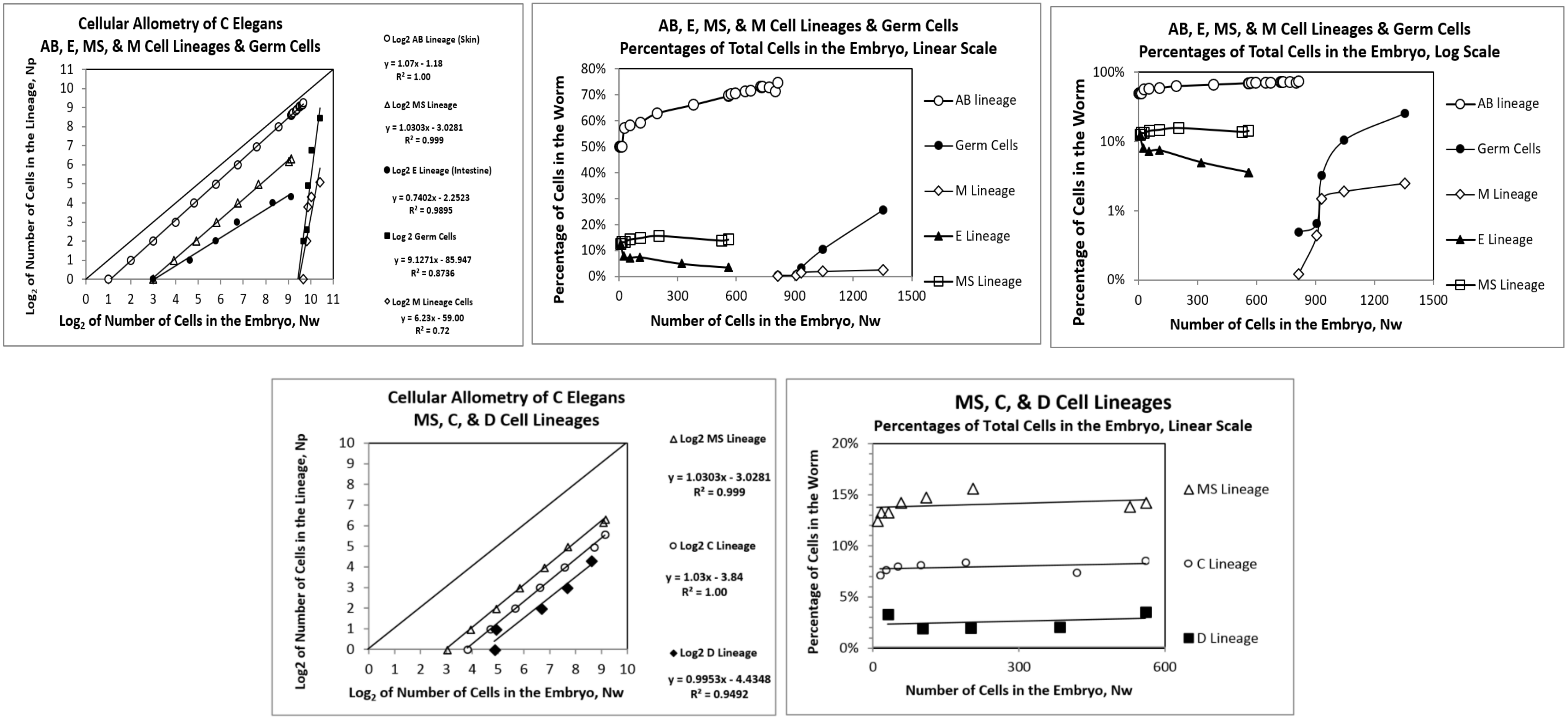


**<u>FIGURE A10</u>**: ***Cellular Allometric Lineage Growth Analysis*** **of the** ***cell lineages*** **of the developing** ***C elegans*** **nematode**

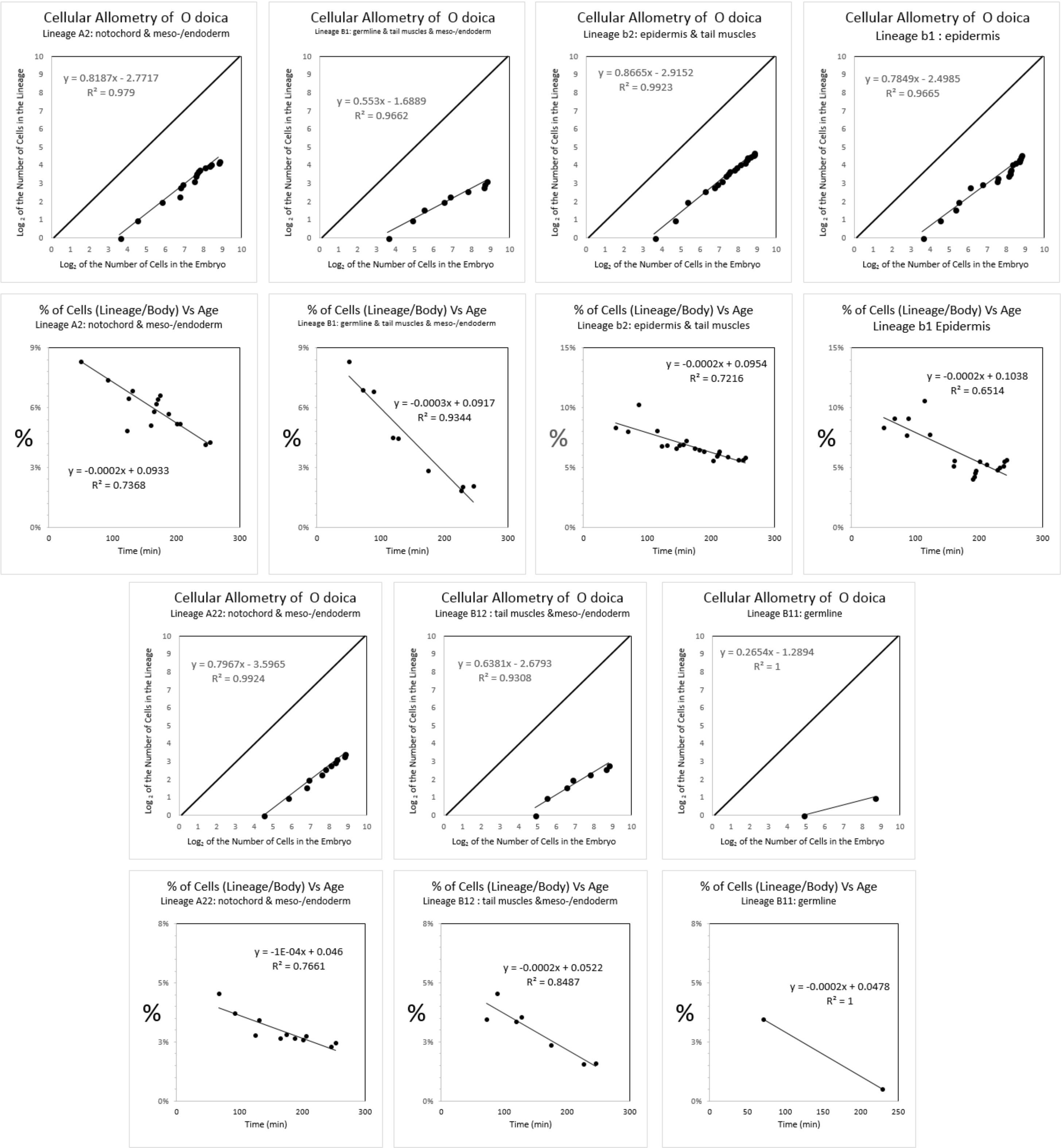


**FIGURE A11:** ***Cellular Allometric Lineage Growth Analysis* of the *cell lineages* of the chordate tunicate *Oikopleura dioica***. Unlike ***C elegans*** and ***M incognita***, all of the cases of ***Cellular Selection*** for this species reflect reductions in cell number, although this could be due to the limited size of the dataset rather than a biological difference. ***Cell lineage chart*** data from Stach et al[88]

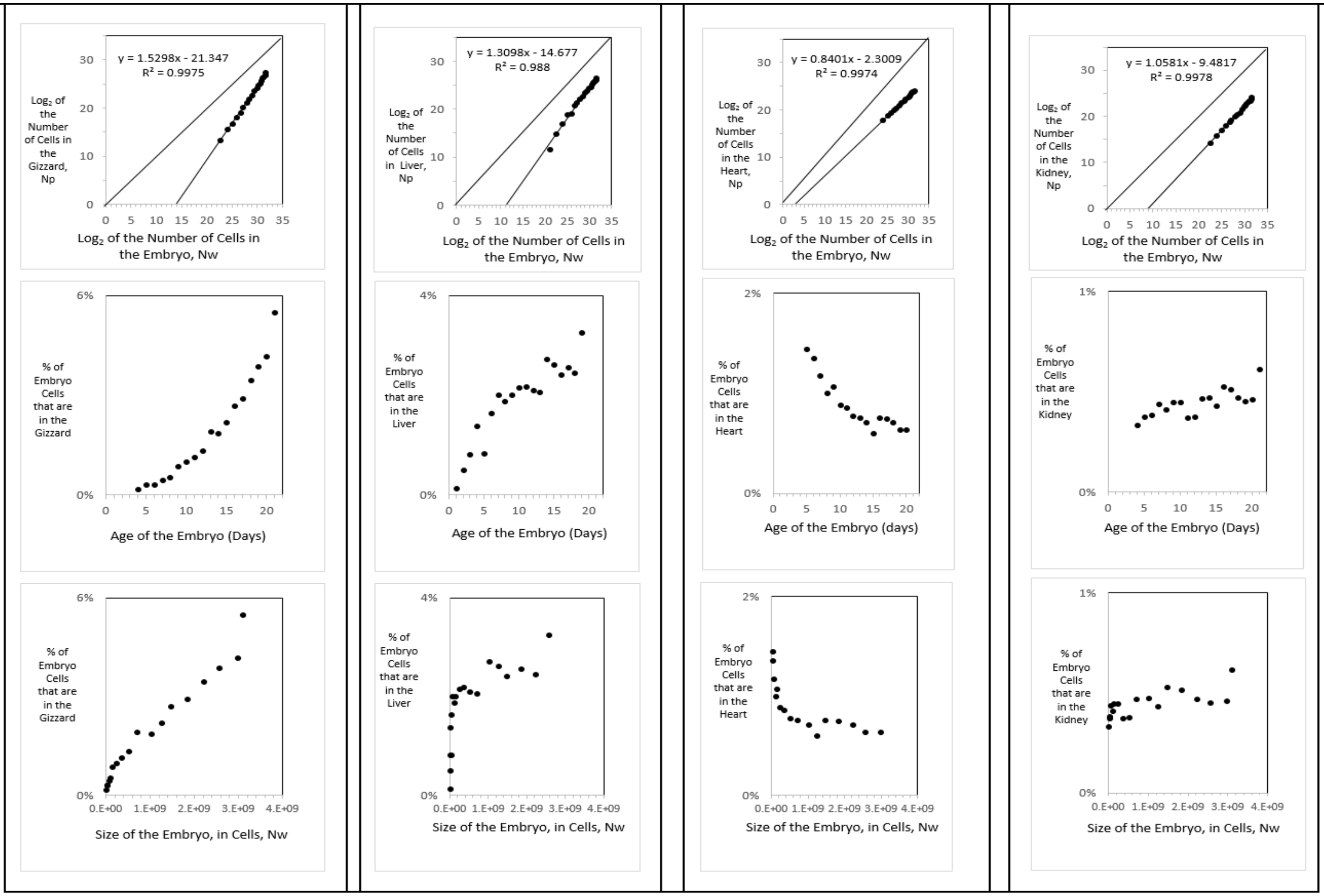


| | % | age | Size of Embryo in Cells | % | age | Size of Embryo in Cells, N | % | age | Size of Embryo in Cells, N | % | age | Size of Embryo in Cells, N |
|---|---|---|---|---|---|---|---|---|---|---|---|---|
| smallest value | 0.17% | 4 | 6.E+06 | 0.14% | 3 | 2.E+06 | 1.35% | 6 | 3.E+07 | 0.33% | 4 | 6.E+06 |
| largest value | 5.49% | 21 | 3.E+09 | 3.26% | 21 | 3.E+09 | 0.64% | 20 | 3.E+09 | 0.61% | 21 | 3.E+09 |
| % increase absolute | 5.32% | | 3.E+09 | 3.12% | | 3.E+09 | -0.72% | | 3.E+09 | 0.28% | | 3.E+09 |
| % increase relative | 33 | | 516 | 23 | | 1475 | 0.47 | | 87 | 2 | | 516 |
| absolute increase relative | 16,990 | | | 33,633 | | | 41 | | | 950 | | |

**<u>FIGURE A12</u>:** ***Cellular Allometric Batch Growth Analysis* of *tissues, organs,* and *anatomical structures* of the chick embryo.** Growth of the parts of the chick embryonic gizzard, liver, heart, and kidney, in units of numbers of cells, in comparison to the number of cells in the embryo as a whole shown on log-log graphs, showing how the fraction of cells comprising in the gizzard and liver increases as the number of cells in the embryo increase with development and how the fraction of cells comprising in the heart decreases as the number of cells in the embryo increase with development, and how the fraction of cells comprising in the kidney remains roughly constant as the number of cells in the embryo increases with development.

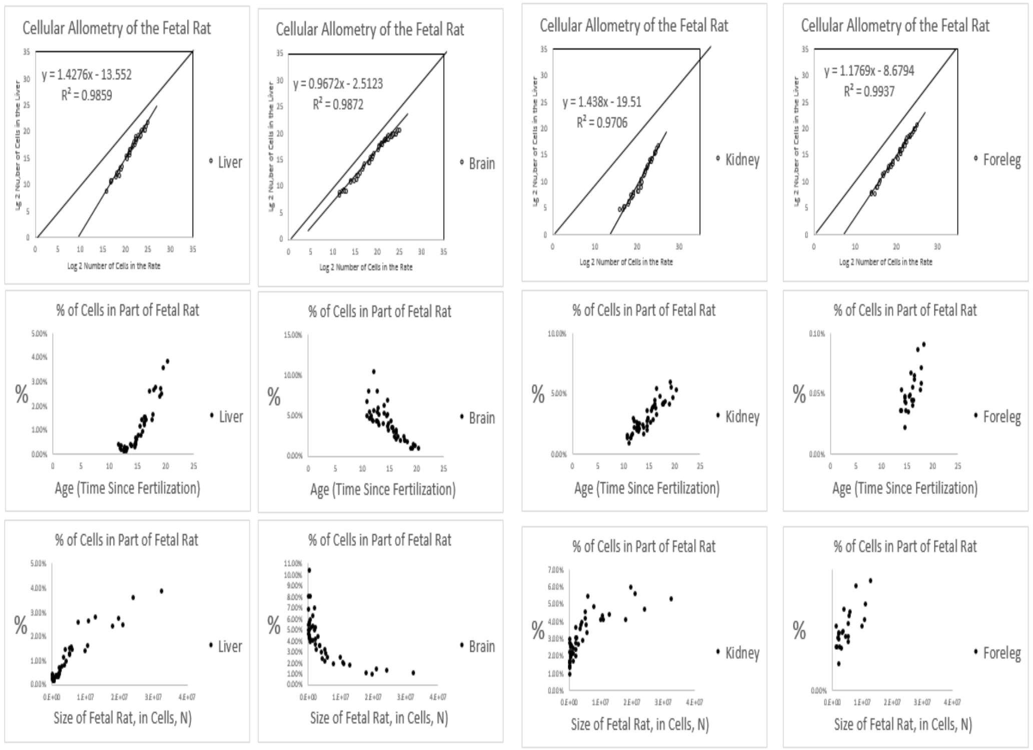


| | % | age | Size of Embryo in Cells, N |
|---|---|---|---|
| first value | 0.46% | 12 | 5.E+04 |
| last value | 3.89% | 20 | 3.E+07 |
| ; increase absolute | 3.43% | | 3.E+07 |
| % increase relative | 8.51 | | 589 |
| te increase relative | 5,012 | | |

| | % | age | Size of Embryo in Cells, N |
|---|---|---|---|
| first value | 6.92% | 11 | 7.E+04 |
| last value | 1.10% | 20 | 3.E+07 |
| % decrease absolute | -5.82% | | 3.E+07 |
| % decrease relative | 6.31 | | 5.E+02 |

| | % | age | Size of Embryo in Cells, N |
|---|---|---|---|
| first value | 1.70% | 11 | 7.E+04 |
| last value | 5.37% | 20 | 3.E+07 |
| % increase absolute | 3.67% | | 3.E+07 |
| % increase relative | 3.16 | | 5.E+02 |
| absolute increase relative | 1,549 | | |

| | % | age | Size of Embryo in Cells, N |
|---|---|---|---|
| first value | 0.04% | 14 | 1.E+06 |
| last value | 0.21% | 20 | 3.E+07 |
| % increase absolute | 0.17% | | 3.E+07 |
| % increase relative | 5.62 | | 29 |
| absolute increase relative | 162 | | |

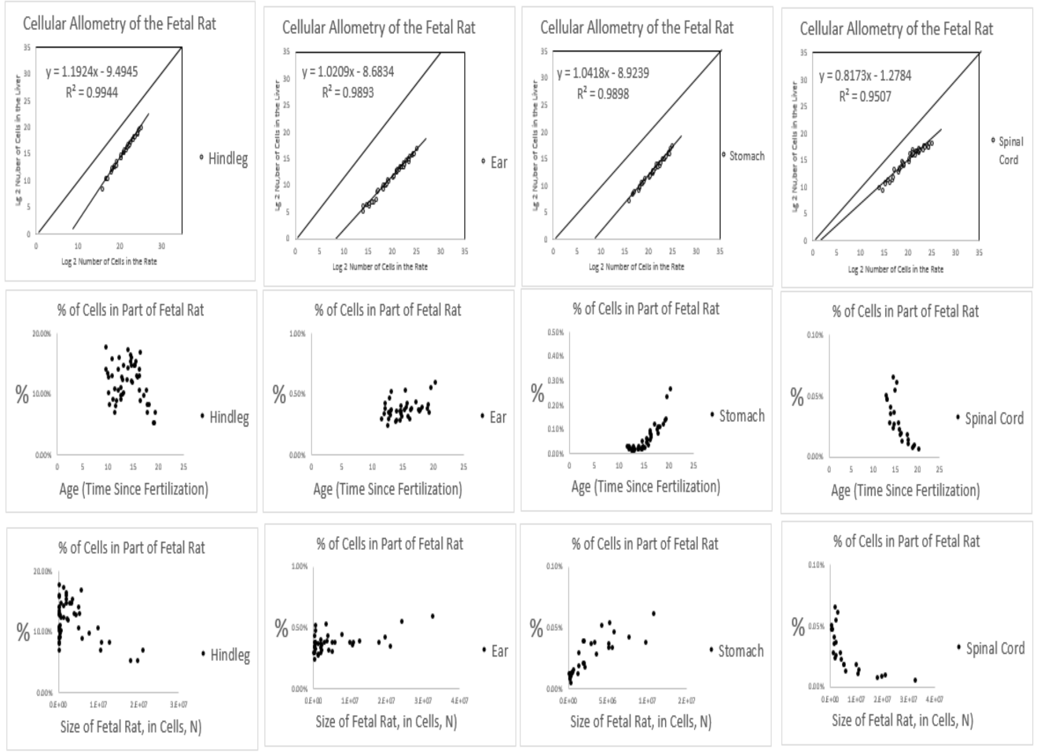


| | % | age | Size of Embryo in Cells, N |
|---|---|---|---|
| first value | 17.78% | 12 | 5.E+04 |
| last value | 5.25% | 20 | 3.E+07 |
| % decrease absolute | -12.53% | | 3.E+07 |
| % decrease relative | 3.39 | | 589 |

| | % | age | Size of Embryo in Cells, N |
|---|---|---|---|
| first value | 0.30% | 12 | 5.E+04 |
| last value | 0.60% | 20 | 3.E+07 |
| % increase absolute | 0.30% | | 3.E+07 |
| % increase relative | 2.00 | | 589 |
| absolute increase relative | 1.175 | | |

| | % | age | Size of Embryo in Cells, N |
|---|---|---|---|
| first value | 0.03% | 12 | 5.E+04 |
| last value | 0.27% | 20 | 3.E+07 |
| % increase absolute | 0.24% | | 3.E+07 |
| % increase relative | 8.13 | | 589 |
| absolute increase relative | 4.786 | | |

| | % | age | Size of Embryo in Cells, N |
|---|---|---|---|
| first value | 0.05% | 12 | 5.E+04 |
| last value | 0.01% | 20 | 3.E+07 |
| % decrease absolute | -0.04% | | 3.E+07 |
| % decrease relative | 0.13 | | 589 |

**FIGURE A13**: ***Cellular Allometric Batch Growth Analysis* of tissues, organs, and anatomical structures of the rat embryo**
Growth of body parts of the rat embryonic liver, brain, kidney and foreleg, in units of numbers of cells, in comparison to the number of cells in the embryo as a whole, are shown on log-log graphs. Note how the fraction of cells comprising in the liver kidney and foreleg increases as the number of cells in the embryo increase with development, while the fraction of cells comprising the brain decreases.

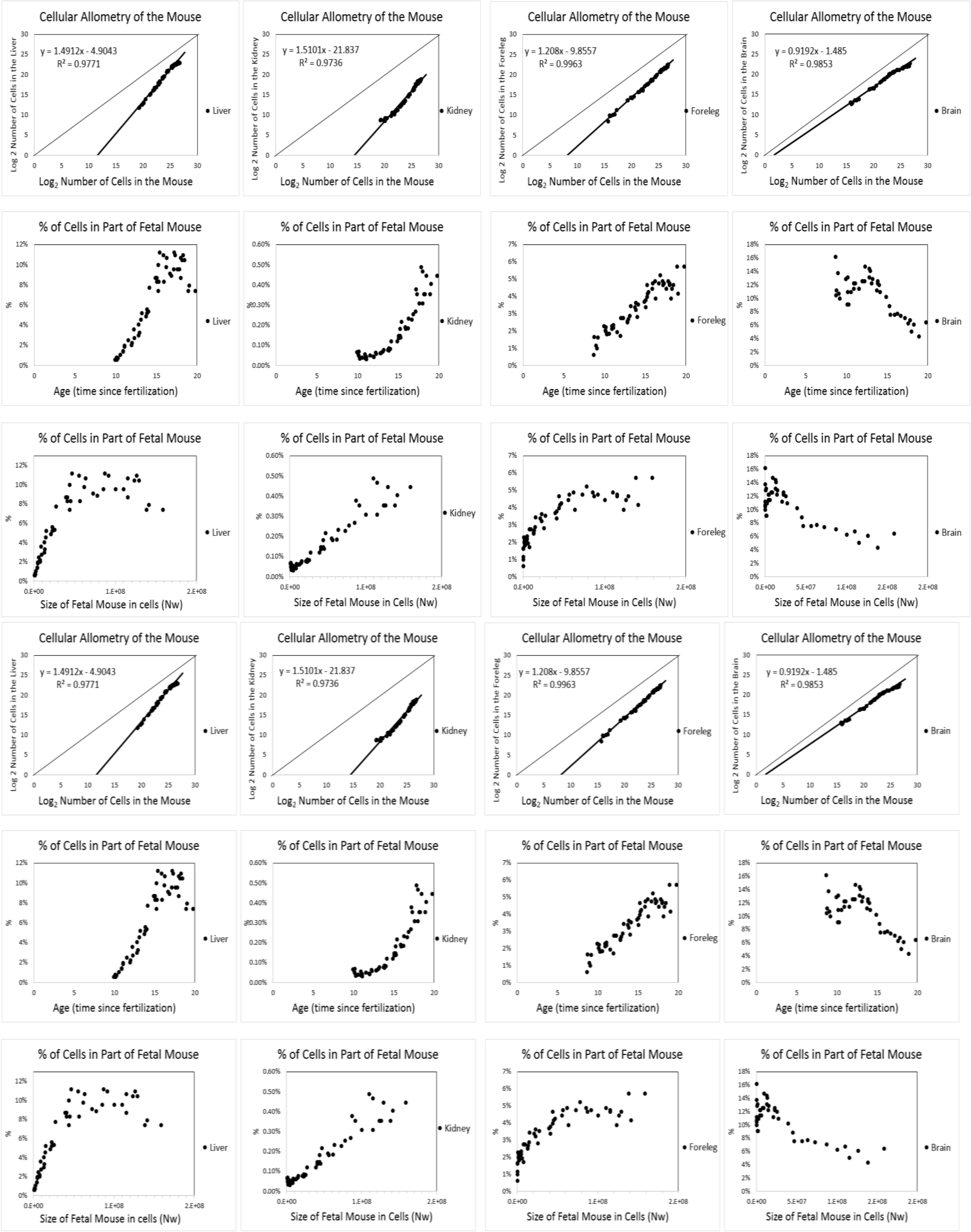


**FIGURE A14**: ***Cellular Allometric Batch Growth Analysis* of tissues, organs, and anatomical structures of the mouse embryo.** Growth of body parts of the mouse embryonic liver, brain, kidney and foreleg, in units of numbers of cells, $N_p$, in comparison to the number of cells in the embryo as a whole, $N_w$, shown on log-log graphs. Note how the fraction of cells comprising in the liver kidney and foreleg increases as the number of cells in the embryo increase with development, while the fraction of cells comprising the brain decreases.

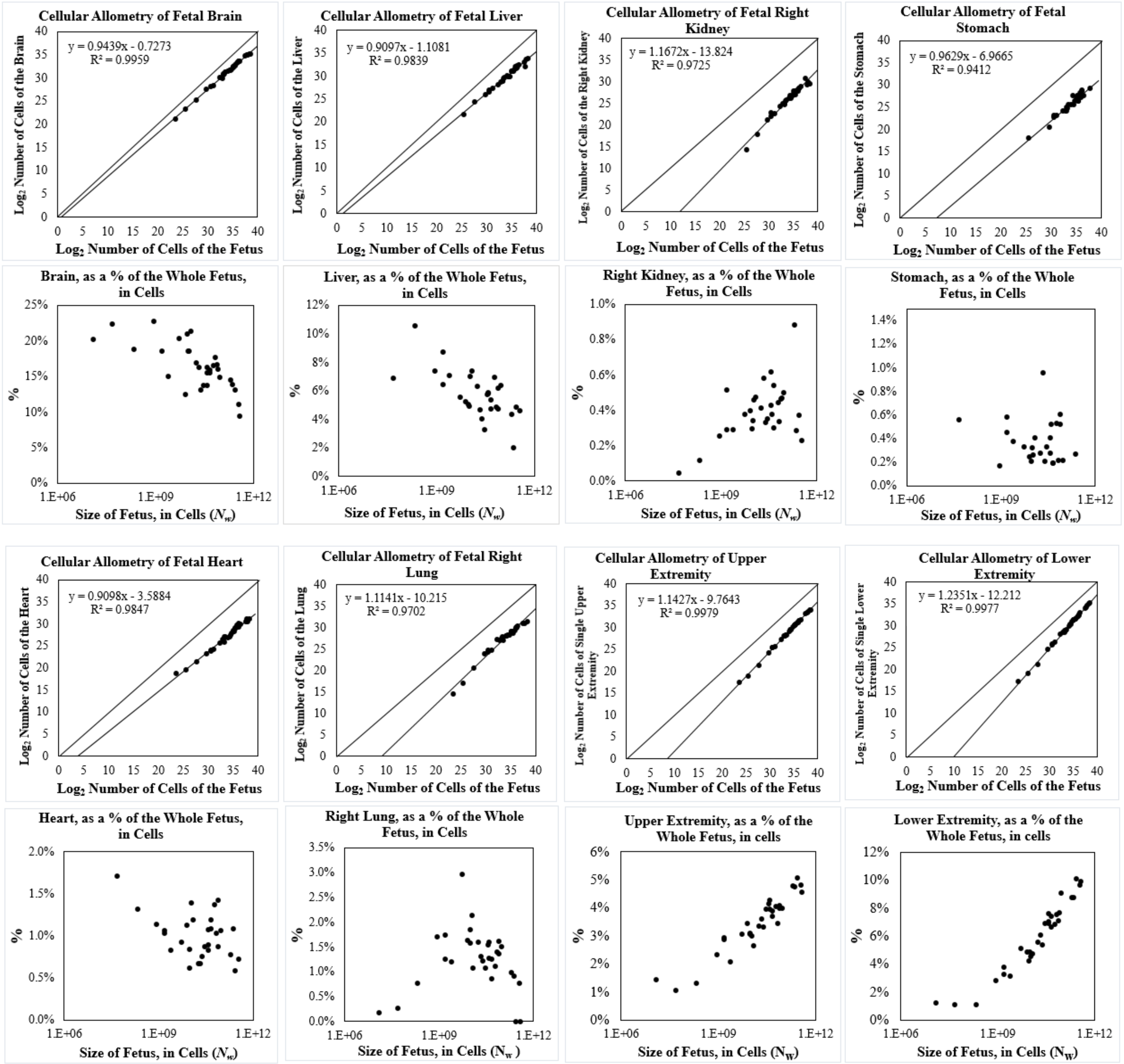


**<u>FIGURE A15</u>: *Cellular Allometric Batch Growth Analysis* of tissues, organs, and anatomical structures of the developing human fetuses, from autopsy data, of individual fetuses**.

For additional human relative growth data, see FIGURE A16

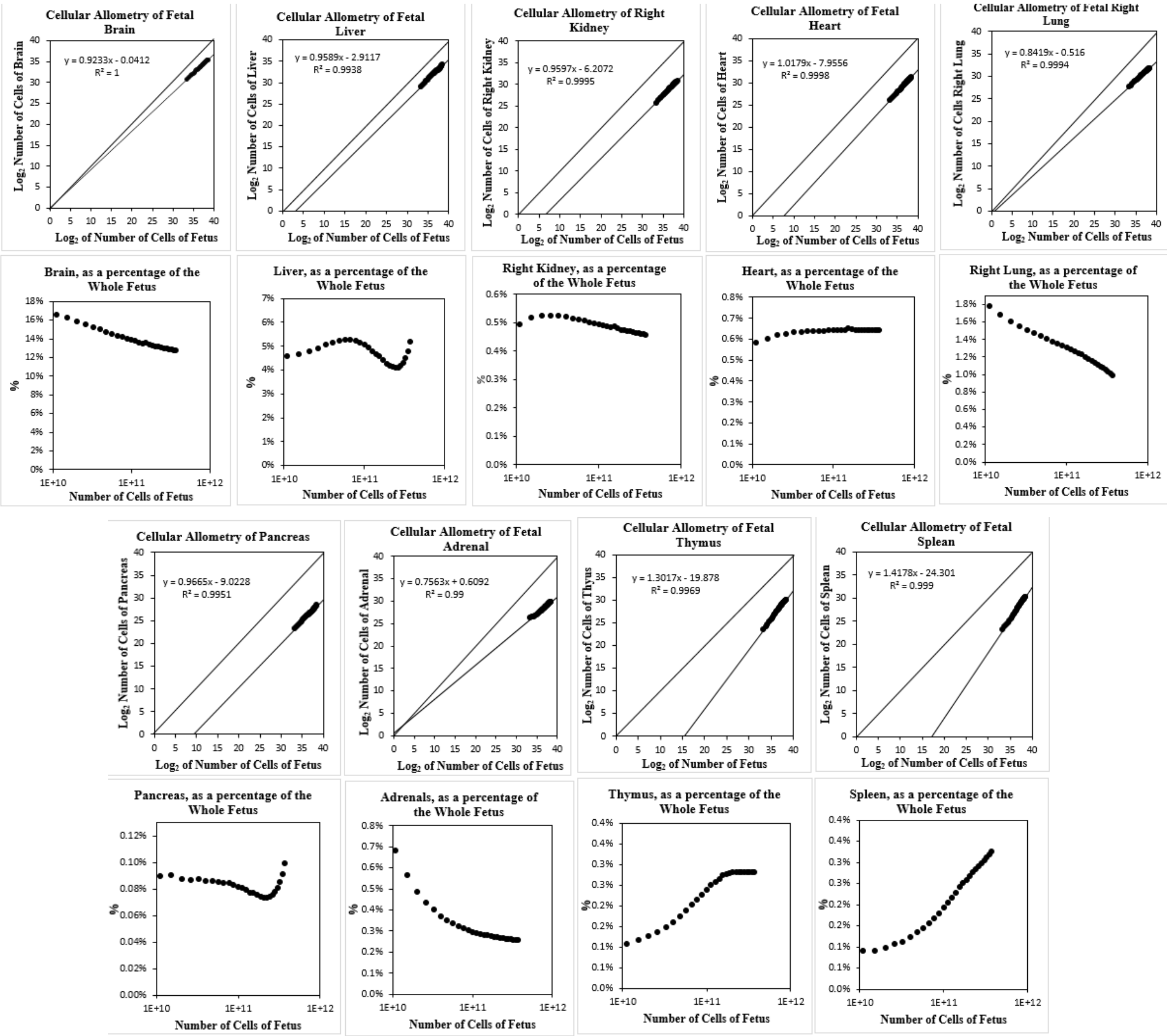


**<u>FIGURE A16</u>: *Cellular Allometric Batch Growth Analysis* of tissues, organs, and anatomical structures of the developing human fetuses, from autopsy data, averaged values**.[91,97]

For additional human relative growth data, see FIGURE A15

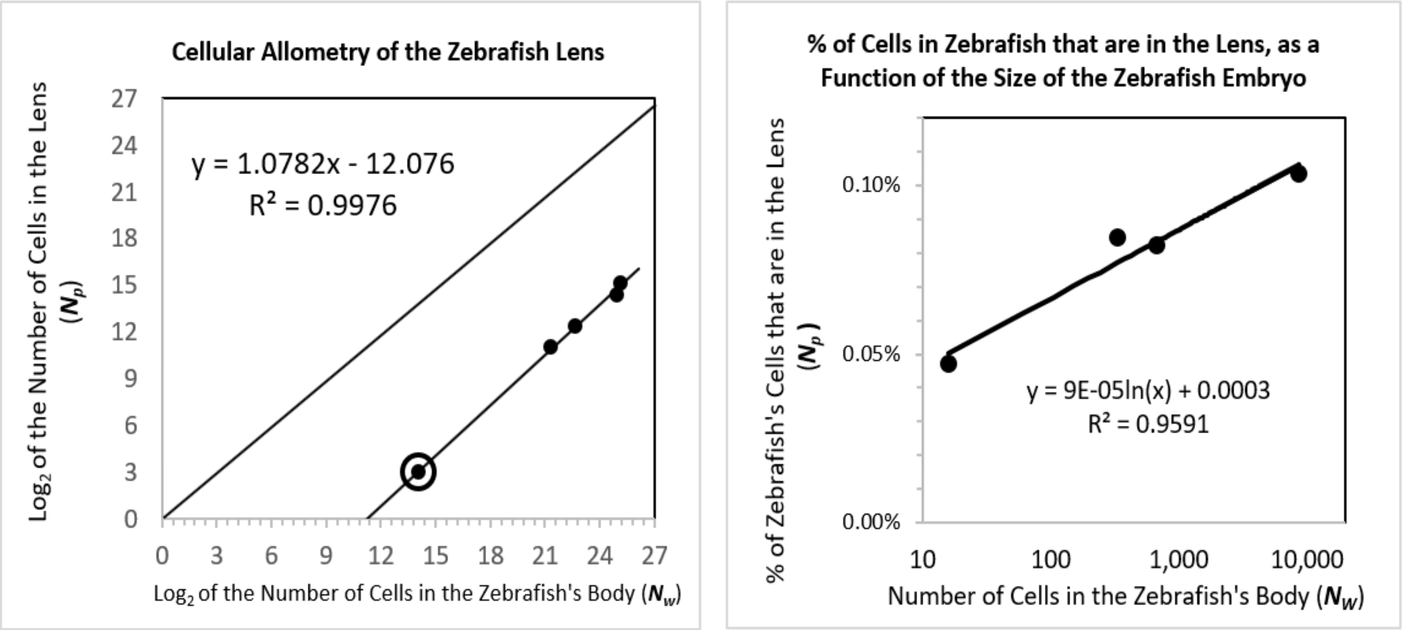


**<u>FIGURE A17</u>: *Cellular Allometric Batch Growth Analysis* of the Zebrafish Lens.**

The ***Cellular Allometric Growth Equation***

With the data outlined above, we were able to compare, on log-log graphs, the number of cells in various parts of the body (***cell lineages***, ***tissues***, ***organs***, and ***anatomical structures***), $N_p$, with the number of cells in the embryo as a whole, $N_w$. Remarkably, in the great majority of these cases, comparisons of $N_p$ vs. $N_w$ appeared as ramrod-straight rows of dots on log-log graphs (FIGURES A9-A17). The high $r^2$ values attest to the strength of these observations of cellular log-linearity. These empirically based observations make plain that ***relative body-part growth***, when examined in numbers of cells, $N_p$ vs $N_w$, often takes the form of the ***Allometric Equation***:

$$\log(\boldsymbol{N_w}) = \frac{\boldsymbol{1}}{\boldsymbol{S_N}} \cdot \log(\boldsymbol{N_p}) + \log(\boldsymbol{B_{N1}}) \quad (4)$$

which is equivalent to:

$$\boldsymbol{N_w} = \boldsymbol{B_N} \cdot \boldsymbol{N_p}^{\frac{1}{Sn}} \quad (4b)$$

We call these expressions "***Cellular Allometric Growth Equations***" (FIGURE A18), and the biological process which is captured "***ALLO-GROWTH***". We call the parameter $B_{N1}$ the "***Cellular Allometric Birth***"' and $S_N$, the "***Cellular Allometric Slope***".

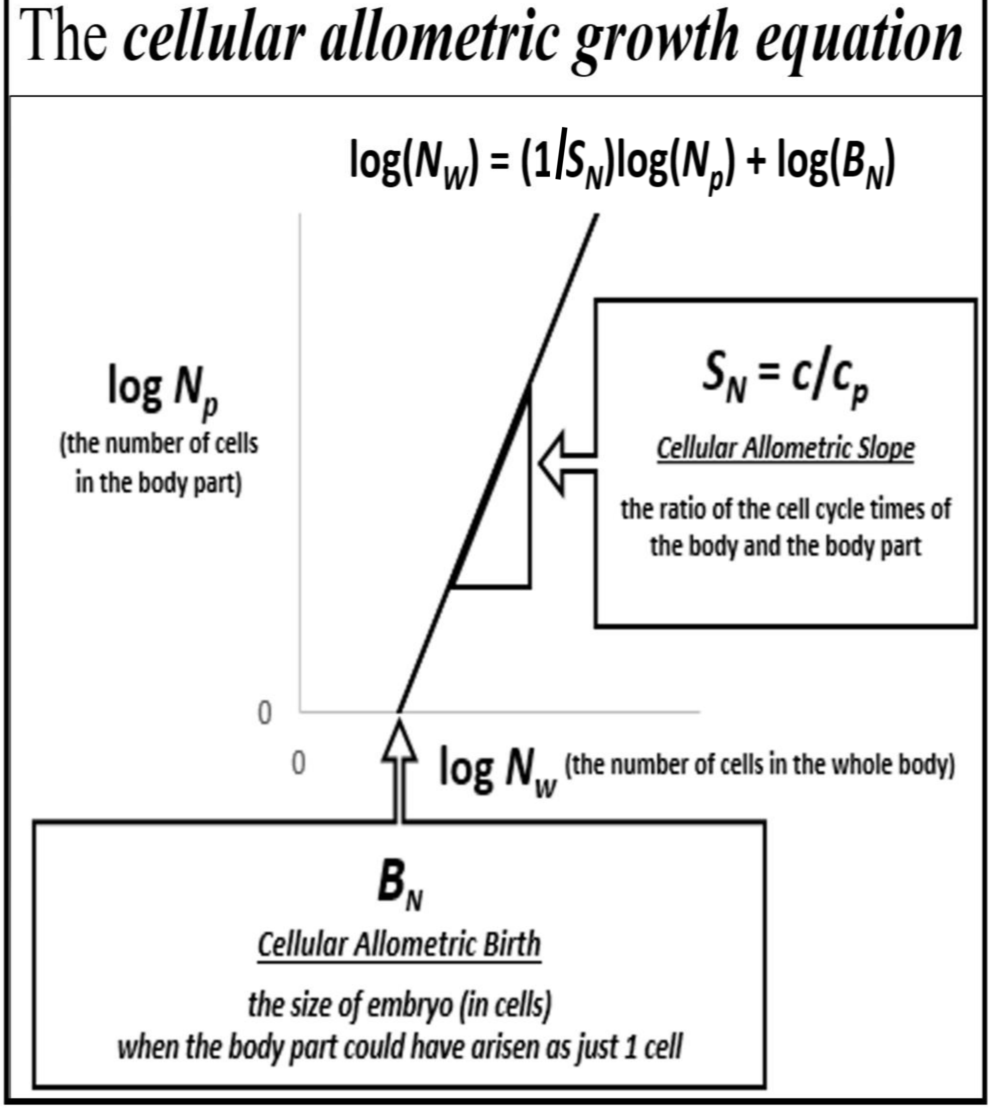


**FIGURE A18: The *Cellular Allometric Growth Equation*, log($N_w$) = (1/$S_N$) log($N_p$) + log($B_{N1}$).**

Body part ***ALLO-GROWTH*** results from whole body ***UNI-GROWTH***.

Recall that the ***Cellular Allometric Growth Equation*** captures the rate of growth of the whole embryo, while the rate of growth for each body-part is:

$$\frac{dN_p}{dt} = \frac{ln(2)}{c_p} \cdot N_p \cdot (m_p) \quad (650)$$

where $c_p$ is the ***Cell Cycle Time***, and $m_p$ the ***Mitotic Fraction*** of the cells of the body part. Thus, the ratio of the rate of growth of the body to the body-part is:

$$\frac{dN_w}{dN_p} = \left[\frac{ln(2)}{c} \cdot N_w \cdot (m)\right] \cdot \left[\frac{ln(2)}{c_p} \cdot N_p \cdot (m_p)\right]^{-1} \text{, where } m = a^{(N_w^b)} \text{ and } m_p = a_p^{(N_w^{bp})} \quad (651)$$

Consider the case where $m_p$=$m$ and $c_p$= $c$, that is, where a body part has the same ***Mitotic Fraction*** and ***Cell Cycle Time*** as the embryo, and starts sometime later than the first cell:

$$\frac{dN_w}{N_w} = \frac{dN_p}{N_p} \quad (652)$$

Integration[1] reveals:

$$log(N_w) = \log(N_p) + \log(B_N) \quad (652)$$

which is the allometric relationship, with $S_N$=**1.** Thus, growth of a part of the body by the ***Cellular Allometric Growth Equation*** is the consequence of ***Founder Cells*** giving rise to cell lineages in animals growing by the ***Universal Growth Equation***.

Parameter $S_N$, the ***Cellular Allometric Slope*** of the ***Cellular Allometric Growth Equation***, tells us how the embryo uses the ***Cell Cycle Time*** to drive ***Cellular Selection*** to adjust the size of each part of the body:

The ***Cellular Allometric Slope***, $S_N$, can be traced to the ***Cell Cycle Time***, $c_p$.

Now, let us consider the case where $m_p$=$m$ but $c_p \neq c$, that is, where a body part has the same ***Mitotic Fraction*** as the embryo but a different ***Cell Cycle Time***:

$$\frac{dN_w}{N_w} = \frac{c_p}{c} \cdot \frac{dN_p}{N_p} \quad (653)$$

Integration reveals:

$$log(N_w) = \frac{c_p}{c} \cdot \log(N_p) + \log(B_N) \quad (653)$$

where:

$$S_N = \frac{c}{c_p} \quad (655)$$

which is the allometric relationship, with $S_N$,≠**1**. Thus, $S_N$, the ***Cellular Allometric Slope*** of the ***Cellular Allometric Growth Equation***, can be traced to the ***Cell Cycle Time***, $c_p$ in each part of the body (FIGURE A18).

On the other hand, similar analysis revealed the ***Mitotic Fraction,*** $m_p$, that is, a change in the fraction of cell dividing, to be an unlikely source for allometric log-linearity. Indeed, changes in the value of either ***a*** or ***b*** of the ***Universal Mitotic Fraction Equation*** (#4), which sets the value of $m_p$, does not result in straight lines on log-log graphs but in curves (not shown).

The ***Cellular Allometric Slope***, $S_N$, and ***Cellular Selection***.

The value of $S_N$, the ***Cellular Allometric Slope*** of the ***Cellular Allometric Growth Equation***, appears on log-log graphs at a measure of whether a part of the body is growing faster ($S_N$>**1**), slower ($S_N$<**1**), or at the same speed ($S_N$=**1**) as the body as a whole. Thus, $S_N$, the ***Cellular Allometric Slope*** of the ***Cellular Allometric Growth Equation*** (#7b) (FIGURE A18) shows how embryos adjust the size of each body part by differential cellular proliferation. These changes in the value of $c_p$ are small, and thus have minor impact on the average ***Cell Cycle Time*** in the embryo as a whole, ***c***, but can have dramatic cumulative impacts on the relative sizes of body parts (FIGURES A8-A17). Thus ***ALLO-GROWTH***, and its mathematical abstraction, the ***Cellular Allometric Growth Equation*** (#7), distills ***Cellular Selection***, that is, differential cellular proliferation,[98,99]down to a single value, $S_N$. Embryos accomplish this by ***Cell-Heritable*** change the ***Cell Cycle Time***, $c_p$.

[1] For a superb YouTube discussion of why the integration of Equation #10 that leads to Equations #7c and #7d, which Salman Khan asserts is "one of the two coolest derivatives in all of calculus", see:
https://www.khanacademy.org/math/in-in-grade-12-ncert/in-in-advanced-differentiation-two/copy-of-proofs-for-derivatives-of-ex-and-lnx-ab/v/proof-d-dx-ln-x-1-x-old
and
https://www.khanacademy.org/math/ap-calculus-ab/ab-antiderivatives-ftc/ab-common-indefinite-int/v/antiderivative-of-x-1

Parameter $S_N$, the ***Cellular Allometric Slope***, at work in adjusting the size of each part of the body

What does the ***Cellular Allometric Slope***, $S_N$, of the ***Cellular Allometric Growth Equation*** (FIGURE A18) tell us about how embryos create our anatomy? The value of $S_N$, the ***Cellular Allometric Slope***, provides us with a measure of how the embryo adjusts the sizes of each of our parts by ***Cellular Selection***, that is, differential cellular proliferation, a remarkably subtle but powerful force in shaping the composition of the embryo.[98,99] We see in $S_N$, the ***Cellular Allometric Slope***, nothing less than the determination of the size of the embryo's ***cell lineages***, ***tissues***, ***organs***, and ***anatomical structures***, by ***Cellular Selection***, driven by the ***Cell-Heritable***, average ***Cell Cycle Time***, $c_p$, of the cells in each of these parts of the body. Thus, the ***Cellular Allometric Growth Equation*** allows us to distill the action of the fundamental morphogenetic force of ***Cellular Selection*** down to a single value, $S_N$.

Parameter $S_N$, the ***Cellular Allometric Slope***, at work early in development

Early in development, we can see ***Cellular Selection*** at work in molding the composition of the first structures of the embryo, the ***cell lineages***. For example, note in FIGURE A10 how the nematode ***C elegans* AB *lineage***, which forms the skin of the worm, has a ***Cellular Allometric Slope***, $S_N$, slightly greater than 1 ($\boldsymbol{S_N \approx 1.1}$); this results in the **AB *lineage*** growing from 50% of the cells of the worm at its creation in the **AB *Founder Cell*** to almost 70% of the worm's body at hatching (FIGURE A10). In the other direction, the ***C elegans* E *lineage***, which forms the intestine, also grows as a straight line on a log-log graph, but with a ***Cellular Allometric Slope***, $S_N$, of less than 1 ($\boldsymbol{S_N \approx 0.7}$); this results in the **E** intestinal ***lineage*** declining from about 12% of the cells of the ***C elegans*** when it first arises from its **E *Founder Cell*** to about 3% at hatching (FIGURE A10). Even more dramatically, the ***C elegans*** germ ***cell lineage*** rises by log-linear ***Cellular Selection*** from a single cell, comprising about 0.02% of the embryo. to about 20% of the adult, roughly a 100-fold increase, through the action of a ***Cellular Allometric Slope***, $S_N$, of ~9 (FIGURE A10). A similar dramatic log-linear post-embryonic increase can be seen for the ***C elegans* M *lineage***, which form muscle cells (FIGURE A10). Many other ***cell lineages*** display such ***Cellular Selection***, as can be seen in FIGURES A9 and A11.

Parameter $S_N$, the ***Cellular Allometric Slope***, at work late in development

Later in development, we again see ***Cellular Selection*** at work molding body part size, this time for ***tissues***, ***organs***, and ***anatomical structures*** (FIGURES A12 - A17). For example, note in FIGURE A12 how the chick's liver starts out as just 0.17% of the cells of the embryo 2 days after the egg is laid, but has grown, with a ***Cellular Allometric Slope***, $S_N$, of ~1.3, to about 5.5% of the embryo by day-21, a 33-fold increase. In the other direction, the chick's heart contains just 1.35% of the embryo's cells at day 2, declining with a ***Cellular Allometric Slope***, $S_N$, of ~0.84, to about 0.64% of the embryo by day-21, a ~2-fold decrease (FIGURE A12). Such ***Cellular Selection*** can be seen to be at work in molding the sizes of many of the ***tissues***, ***organs***, and ***anatomical structures*** of animals, whose log-log graphs can be seen in FIGURES A12 - A17. We found this log-linearity of ***Cellular Selection*** in the relative growth of these embryonic structures of developing mice, rats, clams, and goldfish (data not shown), and human embryos as well (FIGURES A15 and A16).

Parameter, $S_N$, the ***Cellular Allometric Slope***, $S_N$, and the ***Cell Cycle Time***, $c_p$,

Where does the ***Cellular Selection*** captured by the ***Cellular Allometric Slope***, $S_N$, come from? By expanding ***Cellular Allometric Growth Equation*** (#8c), we have been able to see above that the underlying cause of ***Cellular Selection*** can be traced to the average ***Cell Cycle Time*** of the cells in a part of the body, $c_p$, in comparison to the average ***Cell Cycle Time*** of the cells in the body as a whole, $c$, with this expression, which we repeat here for convenience:

$$\boldsymbol{S_N = \frac{c}{c_p}} \qquad (655)$$

In short, the ***Cellular Allometric Growth Equation***, and the relative growth data that it summarizes, tell us that our bodies adjust the size of our body-parts by adjusting the average ***Cell Cycle Time***, $c_p$, of the cells in each of our parts, in a ***Cell-Heritable*** fashion. The ***Cellular Allometric Growth Equation*** is also pointing to possible times when this specification of body part size occurs: the ***Cellular Allometric Birth***. These graphs shown in FIGURES A9-A17, empirical summaries of actual data, of many body parts, in many animals, are also pointing to ***Cellular Allometric Births*** for virtually all body parts, whether large or small, as mapping to very early in development. This suggests that many parts of the body could arise from small numbers of cells, experiencing cell heritable changes in the ***Cell Cycle Time***, $c_p$, very early in development.

The changes in the ***Cell Cycle Time***, $c_p$, that occur within us, and which drive the changes in relative growth that we have seen in the ***Cellular Allometric Slope***, $S_N$, from all of the $N_p$ vs. $N_w$ comparisons we have carried out (FIGURES A9-A17), are generally very small, just a few percentage points. Indeed, these change in the ***Cell Cycle Time***, $c_p$, that occur within the body are tiny in comparison to the hundred-fold differences in the ***Cell Cycle Times***, $c$, which we have seen between different species of animals (Table 2). Nonetheless, these small internal changes in the ***Cell Cycle Time*** can accumulate, often leading to dramatic changes in the body's composition, such as the 33-fold increase noted above in the size of the liver in chick embryos caused by an $S_N$ value of ~1.3.

The role of DNA methylation in the change in the ***Cell Cycle Time***, $c_p$ in body parts

What could cause such a ***Cell-Heritable*** change in ***Cell Cycle Time*** that lies behind the ***Cellular Allometric Slope***, $S_N$, and drives the ***Cellular Selection***, and thus determine the sizes of our ***cell lineages***, ***tissues***, ***organs***, and ***anatomical structures***? There are a number of ***Cell-Heritable*** biological processes that influence the time it takes for a cell to divide, of which DNA methylation has been the most widely studied; methylated DNA takes longer to copy, and thus causes cells to take more time to divide.[100,101]

When might DNA methylation, or some similar ***Cell-Heritable*** process, set the value of the ***Cell Cycle Time***, $c_p$, and thus the ***Cellular Allometric Slope***, $S_N$, of the cells of a part of the body, and thus, ultimately, the size of that part? An appealing possibility for consideration is the moment when that part could have been created, is it arose form a single cell, that is, at its ***Cellular Allometric Birth*** $B_{N1}$, when the part could have been but a single cell. Should this be found to be the case, then the ***Cellular Allometric Birth*** $B_{N1}$, would appear to be the moment when both the part, and its size, are determined by the embryo.

The Parameter, $B_{Na}$, the ***Cellular Allometric Birth*** of the ***Cellular Allometric Growth Equation***, tells us how embryos create body parts from single cells

The value of $B_{Na}$, the ***Cellular Allometric Birth*** of the ***Cellular Allometric Growth Equation***, can be seen easily on log-log graphs (FIGURE A18), as the place where the ***Cellular Allometric Growth Equation*** crosses the ***x***-axis, and thus where $\log(N_p) = 0$, and, therefore, where $N_p = 1$.

Should a part of the body come into existence as a single ***Founder Cell***, the ***Cellular Allometric Birth***, $B_{N1}$, corresponds to the number of cells in the body of the embryo, $N_w$), when that single ***Founder Cell*** arose by mitosis (i.e. $B_N = N_w$ when $N_p = 1$). In fact, this is precisely what has been seen for those animals for which we have ***cell lineage charts***, which have been collected to describe every cell in the embryo from the zygote onward (FIGURE A8), that is, for nematode worms ***Caenorhabditis elegans***[20] and ***Meloidogyne incognita***[21], and for tunicates, ***Oikopleura dioica***,[88] which are chordates quite closely related to ourselves.

For the ***tissues***, ***organs***, and ***anatomical structures*** for which we have growth data later in development, we seldom have data on their cellular origins. There has been much scholarship on this point, but, fortunately, modern light sheet 4D microscopy should allow us to answer these questions,[22,23,102] and the ***Cellular Allometric Growth Equation*** points us to where we should look. Furthermore, whether a body part arises from a single ***Founder Cell***, or multiple ***Founder Cells***, ***Cellular Phylodynamic Analysis*** allows us to characterize the features of the process. For example, should a part of the body come existence from ***z Founder Cells***, each of which is born at the same time, and has the same ***Cell-Heritable Cell Cycle Time***, $c_p$, the part made of these ***z Founder Cells*** will also grow by the ***Cellular Allometric Growth Equation***. Even more complex such examples are amenable to this ***Cellular Phylodynamic Analysis*** approach, a topic we shall address below.

When the parts of the body are born: The ***Cellular Allometric Birth in Time***, $B_T$

The ***Cellular Allometric Growth Equation*** tells us that embryos create our body parts from individual ***Founder Cells***. When does this occur? In terms of the size of the embryo, each part is born when the size of the embryo is $B_{N1}$ ($B_N = N_w$ when $N_p = 1$). In terms of the time when each part is born (relative to the time of fertilization, when $t = 0$), which we call the "***Cellular Allometric Birth in Time***", $B_T$, we can reach back to the ***Universal Growth Equation*** (#5):

$$B_T = \left[\mathrm{Ei}\left(-B_N^b\, log(a)\right) - \mathrm{Ei}(-\,log(a))\right] \frac{c}{b\ log(2)} \tag{660}$$

where $B_T$ is the time of a ***Cellular Allometric Birth***, when a ***Founder Cell*** becomes a body part.

Furthermore, early in development, when most ***Founder Cells*** appear, growth is quite close to exponential, as most cells are dividing. Thus, building from Equation #2:

$$B_T \approx c \cdot (ln(B_N)) \cdot [ln2]^{-1} \tag{661}$$

The time of the ***Cellular Allometric Birth*** of a body part affects its size.

An interesting functional correlate of the single cell origin of ***cell lineages*** is that the structures they form can be big or small by having earlier or later ***Cellular Allometric Births***, $B_{N1}$. For example, while the **MS**, **C**, and **D** ***lineages*** of ***C elegans*** all grow on log-log graphs right along with the embryo as a whole, thus with ***Cellular Allometric Slopes***, $S_N$, close to **1**, the cells of the **MS** ***lineage*** make up about 12%-14% of the worm because it is born at the 8-cell stage ($B_N = 8$), while the cells of the **C** ***lineage*** make up about 8% of the worm by being born at about the 16-cell stage ($B_N = 16$), and the cells of the **D** ***lineage*** makes up about 3% of the worm by being born at about the 32-cell stage ($B_N = 32$) (FIGURE A11). Thus, the option of generating structures from single cells at various points in the growth of the body as a whole gives the embryo another trick for adjusting the relative sizes of its many parts.

Data on the number of ***Founder Cells*** that make various parts of the body.

Early in development, we can see the creation of body parts from single ***Founder Cells*** for those animals for which ***cell lineage charts*** have been made. For example, note in FIGURES A9 and A10, how the nematode ***C elegans*** **AB** ***Founder Cell*** arises when the embryo is just 2 cells in size, whose progeny go on to form the **AB** ***lineage***, which forms the skin from the ***Cellular Allometric Birth*** of $\boldsymbol{B_N = 2}$ (i.e., $\boldsymbol{N_w = 2}$ when $\boldsymbol{N_p = 1}$). Similarly, note in FIGURES A9 and A10, how the ***C elegans*** **E** ***Founder Cell*** arises when the embryo is 8 cells in size, whose progeny go on to form the **E** ***lineage***, which forms the intestine from the ***Cellular Allometric Birth*** of $\boldsymbol{B_N = 8}$ (i.e., $\boldsymbol{N_w = 8}$ when $\boldsymbol{N_p = 1}$). Many other such examples of the creation of body parts from single ***Founder Cells*** can be seen for ***C elegans*** and ***M incognita*** nematode worms and ***O dioica*** chordate tunicates in FIGURES A9 and A10.

Later in development, we get hints of the creation of body parts from single ***Founder Cells***, as the ***Cellular Phylodynamic Analysis*** of the ***tissues***, ***organs***, and ***anatomical structures*** of the body shows that the line of the ***Cellular Allometric Growth Equation*** points down to the spot on the $\boldsymbol{x}$-axis where the ***Cellular Allometric Birth***, $\boldsymbol{B_{N1}}$, lies (FIGURES A12-A17). Remarkably, in almost every case, the line of the ***Cellular Allometric Growth Equation*** points to a ***Cellular Allometric Birth***, $\boldsymbol{B_{N1}}$, that corresponds to the number of cells in the body of the embryo, $\boldsymbol{N_w}$ when the embryo was quite early in its development, and never to numbers of cells less than **2**. For example, as can be seen in FIGURE A12, $\boldsymbol{B_N \approx 8}$ for the chicken heart, suggesting that the heart could have arisen from just 1 cell when the embryo was ~8 cells in size. For the chicken liver, $\boldsymbol{B_N \approx 2,000}$, suggesting that the liver could have arisen from just 1 cell when the embryo was ~2,000 cells in size (FIGURE A12). Many other such examples of the creation of body parts from single ***Founder Cells*** can be seen in the relative growth of these embryonic structures of developing mice, rats, clams, and human embryos as well (FIGURES A15 and A16). While few of these large structures have been measured down to the $\boldsymbol{B_{N1}}$ intersection point, the zebrafish eye lens comes close, as Greiling and Clark documented its growth from just 8 cells, when the embryo is 16 hours post fertilization, with the lens's ***Cellular Allometric Growth Equation*** pointing down to a ***Cellular Allometric Birth*** when the embryo was about 2000 cell in size, and thus $\boldsymbol{B_N \approx 2,000}$ when $\boldsymbol{N_p = 1}$ (FIGURE A17).[93-95] These observations complement other studies that have also suggested that large anatomical structures may arise from small numbers of cells,[103,104] including recent CRISPR/Cas9 ***cell lineage*** labeling studies.[105,106]

Body parts can be made of one ***Founder Cell*** or more than one ***Founder Cell***

Curiously, whether a part of the body arises from **1** ***Founder Cell***, or more than **1** ***Founder Cell***, ***ALLO-GROWTH*** will result in body parts growing by the ***Cellular Allometric*** and ***Allometric Growth Equations***, and, early in development, by the ***Exponential Growth Equation***, if all ***Founder Cells*** of a body part arise at the same time and with the same ***Cell Cycle Time***, $\boldsymbol{c_p}$. This could be seen in a simple calculation of an idealized part of the body that is made from **3** ***Founder Cells***, arising early in development, when growth is close to exponential (FIGURE A19). As can be seen in the top two graphs of FIGURE A19, if all three ***Founder Cells***, arise at the same time, and have the same ***Cell Cycle Time***, $\boldsymbol{c_p}$, their growth together will be indistinguishable from a part made of **1** ***Founder Cell*** (once it gets up to 3+ cells!). Furthermore, as can be seen in the other graphs of FIGURE A19, no matter when the three ***Founder Cells*** are born, or what their ***Cell Cycle Times***, $\boldsymbol{c_p}$, are, the aggregate growth of the part will still appear as a quite straight row of datapoints on a log graph. Thus, the fact that the growth of a body part points back to a single ***Cellular Allometric Birth***, doesn't mean that it arose from a single cell; we still have to do the microscopy work to see what actually happened.[22,23,93,107]

This simple excise also shows, however, that body parts made of more than a single ***Founder Cell*** do have some challenges. Note for example, that ***Founder Cells*** born after the first ***Founder Cell***, or having a lower ***Cell Cycle Time***, $\boldsymbol{c_p}$, than the fastest growing ***Founder Cell***, will soon become irrelevant, and the body part will become functionally monoclonal. A ***Founder Cell*** born three ***Cell Cycle Times***, $\boldsymbol{c}$, after the first ***Founder Cells***, which in a human or a mouse is about three days, will make up only about 10% of the part, while a ***Founder Cell*** born eight ***Cell Cycle Times***, $\boldsymbol{c}$, after the first ***Founder Cell*** will make up only about 1% of the part. ***Founder Cells*** that have ***Cell Cycle Times***, $\boldsymbol{c_p}$, slower than the fastest ***Founder Cell*** will soon loose in the race to make up a significant percentage of the part.

While these this simple exercise examined the idealized case of a part of the body arising early in development, when growth is close to exponential, the results shown here are likely to be similar later in development as well, when ***UNI-GROWTH*** begins to be felt, although exploring precise details for this would be worthwhile.

Of course, we can imagine all sorts of complicated multi-clonal anatomical structures. Perhaps their ***Cell Cycle Times*** are elegantly intertwined, or they adjusted their ***Mitotic Fractions*** to stay in league. Fortunately, however, whatever the cellular behavior, if one collects the cell numbers, ***Cellular Phylodynamic Analysis*** provides us with a way to isolate the underlying processes.

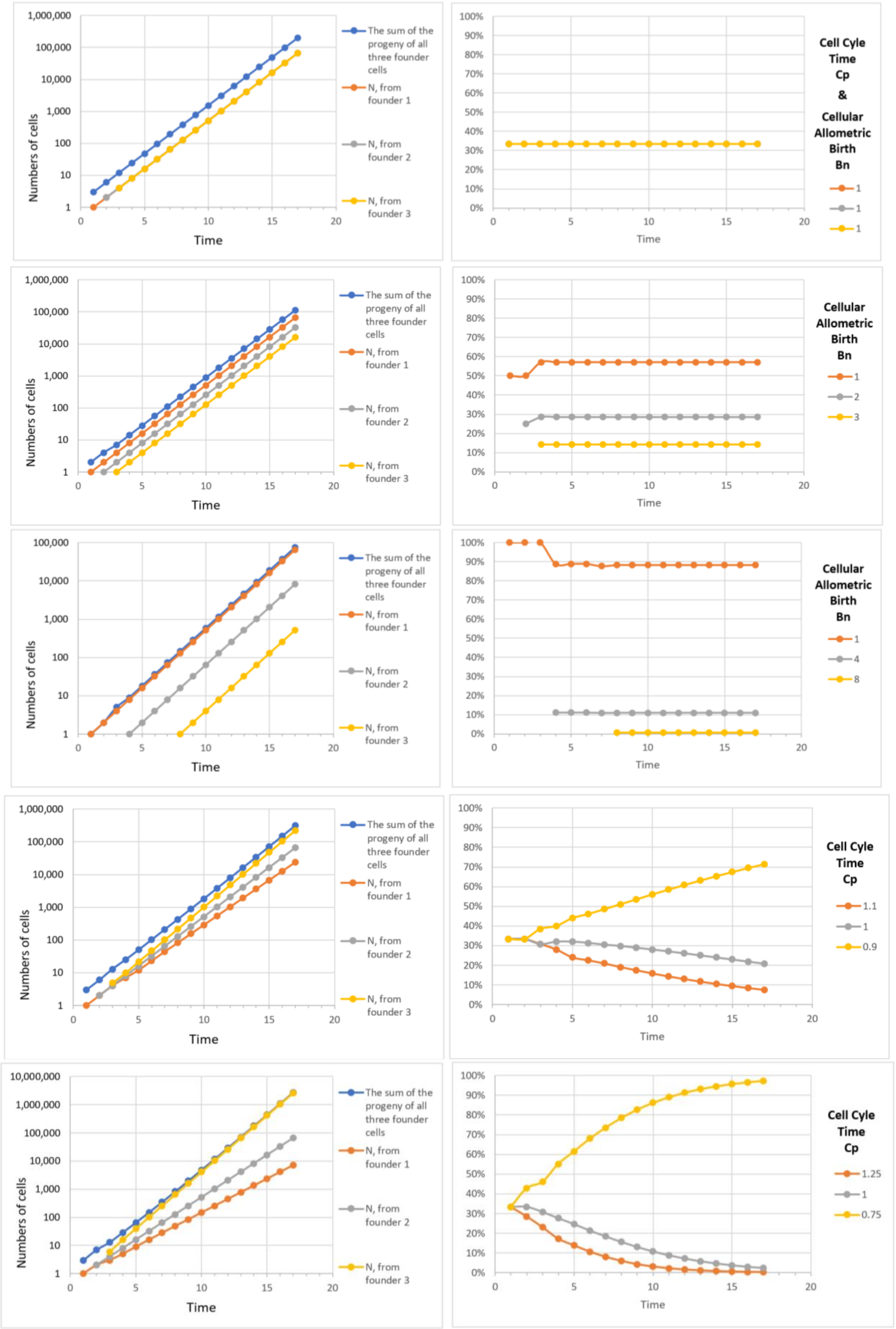


**<u>FIGURE A19</u>: Calculations for an idealized part of the body that is made from 3 *Founder Cells*, arising early in development**

Body Part Size to Body Part Size Relationships; ***Body Part Proportion Equations***

Let us now consider the growth of two parts of the body, $\boldsymbol{p_1}$ and $\boldsymbol{p_2}$, both of which display allometric growth with reference to the growth of the body as a whole, ***w***.

For part 1:

$$\log(N_w) = \left(\frac{S_{N1}}{c}\right)\log(N_{p1}) + \log(B_{N1}) \tag{719}$$

For part 2:

$$\log(N_w) = \left(\frac{S_{N2}}{c}\right)\log(N_{p2}) + \log(B_{N2}) \tag{720}$$

It follows that:

$$\log(N_w) = (S_{N1})\log(N_{p1}) + \log(B_{N1}) = (S_{N2})\log(N_{p2}) + \log(B_{N2}) \tag{721}$$

which is equivalent to:

$$\log(N_{p1}) = \left(\frac{S_{N2}}{S_{N1}}\right)\log(N_{p2}) + \log\left(\frac{B_{N2}}{B_{N1}}\right) \tag{721b}$$

In other words, if two parts of the body show log-linear allometric growth with respect to the size of the body as a whole, they will also show log-linear allometric growth with respect to each other, with each part's ***Cellular Allometric Birth***, $\boldsymbol{B_N}$ and ***Cellular Allometric Slope***, $\boldsymbol{S_N}$ repackaged into the slope and intercept of this new variation on the ***Cellular Allometric Growth Equation***.

It follows that if the body parts in question are in units of mass or volume or length, we can repeat the treatment outlined in the previous section. Thus:

$$\log(p_1) = \left(\frac{1}{S_{1,2}}\right)\log(p_2) + \log(B_{1,2}) \tag{722}$$

which is equivalent to:

$$p_1 = B_{1,2} \cdot p_2^{\frac{1}{S_{1,2}}} \tag{722b}$$

We call these expressions ***Body Part Proportion Equations***.

***Clones Within Clones***

As can be seen in Equation #722 above, if each of two parts of the body are seen by ***Cellular Allometric Growth Analysis*** to conform to the ***Cellular Allometric Growth Equation*** (#7), when each is compared with the body as a whole, they will also conform to the ***Cellular Allometric Growth Equation*** when compared with each other. An actual example of this can be seen in FIGURE A23, in which we display the number of cells in the anterior part of the drosophila wing, $\boldsymbol{N_{p-a}}$, in comparison to the number of cells in the wing as a whole, $\boldsymbol{N_{p-w}}$. These values were generated from measurements of the area of the whole wing disc, and of its anterior part, that have been collected by Parker and Shingleton,[108] translated into likely cell numbers, $\boldsymbol{N_{p-a}}$, and $\boldsymbol{N_{p-w}}$, with cell number per area data derived by Ulrike.[109]

These log-log comparisons of $\boldsymbol{N_{p-a}}$ (number of cells in the anterior part of the drosophila wing), and $\boldsymbol{N_{p-w}}$ (number of cells in the wing as a whole), are shown in FIGURE A23. This **log**($\boldsymbol{N_{p-a}}$) vs **log**($\boldsymbol{N_{p-w}}$) comparison reveals that the ***Cellular Allometric Growth Equation*** captures the growth of the anterior part of the wing, pointing to a ***Cellular Allometric Birth***, $\boldsymbol{B_{N1}}$, growing from when the wing as a whole was 8 cells in size ($\boldsymbol{B_{N1}=2^3=8}$), should the anterior part have arisen from a single ***Founder Cell***. Furthermore, the ***Cellular Allometric Slope***, $\boldsymbol{S_{Np}}$, is greater than **1**, indicating an increase in speed of the ***cell cycle time***, $\boldsymbol{c_p}$, of the cells in the anterior part of the wing, thus accounting for the progressively increasing relative size of the anterior part of the wing in relationship to the wing as a whole (FIGURE A24). We call this process "***Clones Within Clones***", a mechanism which provides animals a way to create functional modularity.

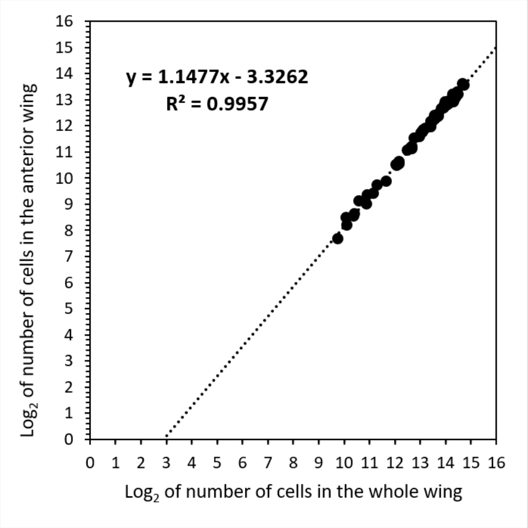


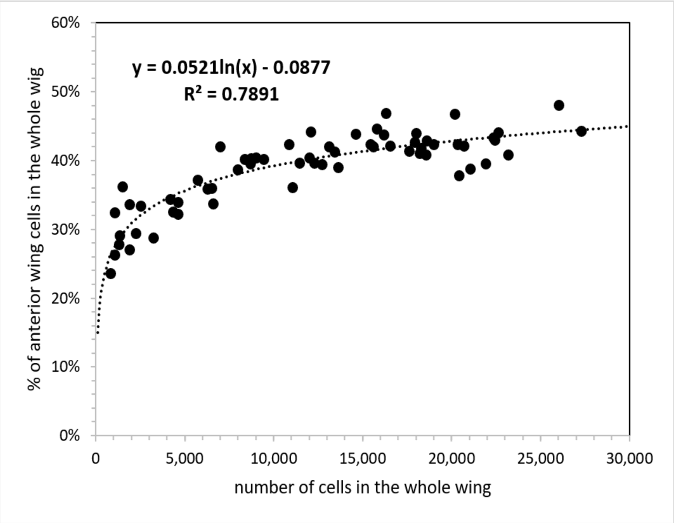


**FIGURE A23: Left. *Cellular Allometric Growth Analysis* of the anteriors wing vs whole wing.**

**FIGURE A24: Right. Percentage of anteriors wing cells in the whole wing vs number of cells in the whole wing.**

These observations are also strikingly complementary to the long appreciated finding the anterior and posterior parts of the fly's wing are clonally separate entities, called ***compartments***.

Whether the anterior part of the wing grew from a single ***Founder Cell***, at the ***Cellular Allometric Birth***, $\boldsymbol{B_{N1}}$, or from more than **1** ***Founder Cell***, after the ***Cellular Allometric Birth***, $\boldsymbol{B_{N1}}$, cannot be known from the data we have, since the values for $\boldsymbol{N_{p-a}}$, and $\boldsymbol{N_{p-w}}$ only point down to that origin at when the wing as a whole was 8 cells in size ($\boldsymbol{B_{N1}=2^3=8}$). Nonetheless, this poses an eminently answerable question, which could be settled by light sheet microscopy, again providing motivation for experiment.[22,23,93]

The formation of ***Clones Within Clones*** occurs in many instances of embryonic development of many animals. For example, the melanocytes of the skin of mice appear to be derived from 34 ***Founder-Cells*** (17 on each side of the body) created around the 12th day of life.[110] ***Clones Within Clones*** are most dramatically at work in the immune system, where ***clonal selection*** selects individual cells and expand them up to large populations of cells through the action of antigens stimulating cell division by attaching to antibodies or T-cell receptors at the cell surface.[98,99,111] Such a viewpoint may be especially useful in comprehending and learning how to maximize COVID-19 vaccines' impact. Similar processes may well occur in other systems, such as in the production of plasma proteins by the liver, in which each of the hundred-or-so blood proteins produced by the liver appears to be made by separate cells, in separate clones, that arise as patches continuously throughout life.[83,99] Anatomically, the creation of such functional diversity by the creation of ***Clones Within Clones*** has been seen time after time, such as in the clones that form the crypts of the intestine,[112] and in the sequential clones of spermatocytes that produce the spermatozoa,[113] as well as the many anatomical structures that seem to be made of countless subunits, such as in the lungs, the liver, the kidney, and so on. ***Cellular Phylodynamic Analysis*** seems poised to make sense of how cells create such modular structures.

***ALLO-GROWTH***: Summary Definition

Let us summarize: ***ALLO-GROWTH*** is the process by which body parts are created from ***Founder Cells***. ***ALLO-GROWTH*** is captured by the ***Cellular Allometric*** and ***Allometric Growth Equations***, and their parameters, $\boldsymbol{S}$, $\boldsymbol{B}$, $\boldsymbol{S_N}$, the ***Cellular Allometric Slope***, and $\boldsymbol{B_{N1}}$, the ***Cellular Allometric Birth***. ***ALLO-GROWTH*** occurs by a ***Founder Cell*** acquiring a ***Cell-Heritable***, ***Cell Cycle Time***, $\boldsymbol{c_p}$ at the ***Founder Cell's Cellular Allometric Birth***, $\boldsymbol{B_{N1}}$, and then undergoing mitotic expansion, as captured by the ***Cellular Allometric Slope***, $\boldsymbol{S_N}$, of the ***Cellular Allometric Growth Equation***. Whether a body part arises from a single ***Founder Cell***, or multiple ***Founder Cells*** arising at the same time with the same ***Cell Cycle Time***, growth by the ***Cellular Allometric*** and ***Allometric Growth Equations*** will occur. For body parts arising from a single ***Founder Cell***, the ***Cellular Allometric Birth***, $\boldsymbol{B_{N1}}$, corresponds to the number of cells in the body when the ***Founder Cell*** arose.

#### AGING

***Mitotic Fraction* and Age**

The relationship between ***age***, ***t***, and ***Mitotic Fraction***, ***m***, can be calculated by combining Equations #1 and #3:

$$\boldsymbol{t}(\boldsymbol{m}) = \frac{\boldsymbol{c}}{\boldsymbol{b}\cdot\boldsymbol{log}(2)}\cdot \quad [\mathrm{Ei}(-\boldsymbol{log}(\boldsymbol{m})) \; - \; \mathrm{Ei}\,(-\boldsymbol{log}(\boldsymbol{a}))] \qquad (5)$$

We call this expression, the ***Universal Mitotic Fraction in Time Equation***. With it, we could generate curves for the 10 species listed above, and in TABLE II, which, when ended at the age of the longest known individual in each species, reveals lifespan to be correlated with an age when fewer than 1-in-1,000 cells are dividing (FIGURE 2 in text, FIGURES A 26 and A21). We call this region the ***Death Zone***, marking the thousand+fold decline in ***Mitotic Fraction*** from conception to death.

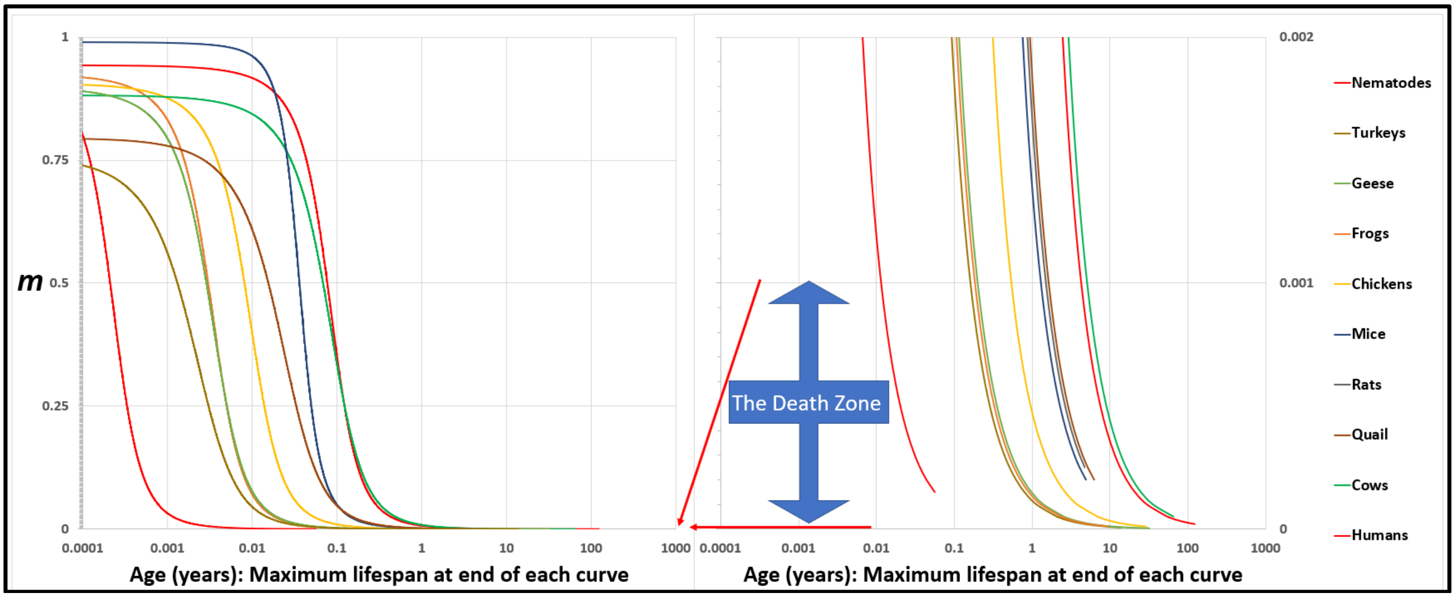


**FIGURE A20: The *Universal Mitotic Fraction in Time Equation*** (#5) **captures change in *Mitotic Fraction*, *m*, with *Age*, *t*.**

$$\boldsymbol{t}(\boldsymbol{m}) = \frac{\boldsymbol{c}}{\boldsymbol{b}\cdot\boldsymbol{log}(2)}\cdot \quad [\mathrm{Ei}(-\boldsymbol{log}(\boldsymbol{m})) \; - \; \mathrm{Ei}\,(-\boldsymbol{log}(\boldsymbol{a}))] \qquad (4)$$

The graph on the right side shows the bottom 0.2% of the Y-axis of the graph on the left side: the ***Death Zone*** occurs when fewer the 0.03% of the cells of the body are dividing. Each curve ends at the age of the oldest known member of its species.

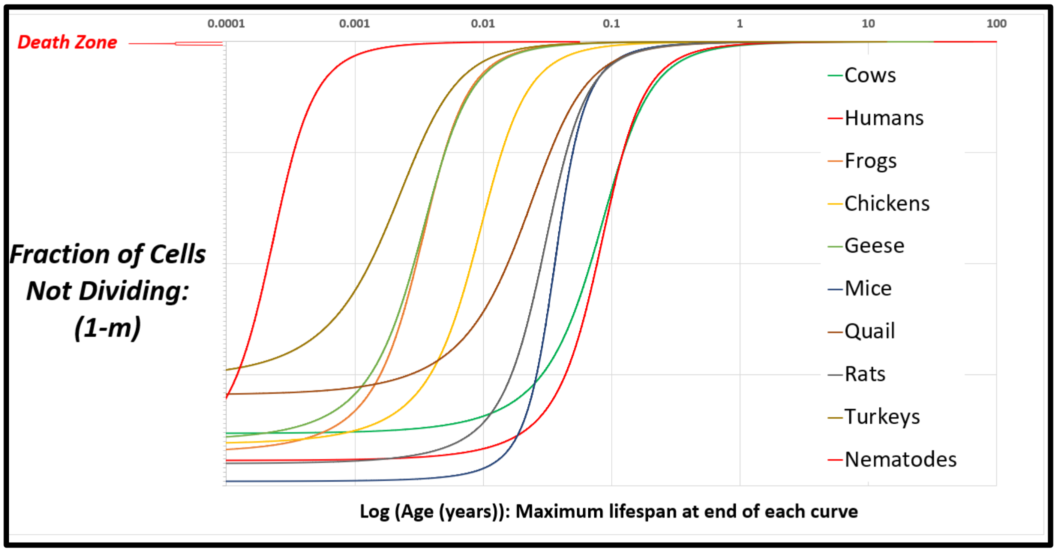


**FIGURE A21: The inverse of the *Universal Mitotic Fraction in Time Equation*** (#5) **visualizes the change in the fraction of cell not dividing with age.**

##### Lifespan

All three parameters of the ***Universal Growth Equation***, $\boldsymbol{a}$, $\boldsymbol{b}$ and $\boldsymbol{c}$, considered genetically, allow species to evolve to patterns of growth that give the greatest chances of survival, and to lifespans calculable with this expression:

$$\boldsymbol{Life\ Span} = \frac{\boldsymbol{c}}{\boldsymbol{b}\cdot\boldsymbol{log(2)}}\cdot\ \ [\mathrm{Ei}\big(-\boldsymbol{log(Z)}\big)\ -\ \mathrm{Ei}\ \big(-\boldsymbol{log(a)}\big)] \tag{6}$$

$\boldsymbol{Z}$ is the ***Mitotic Fraction*** at the end of life. We call Equation#5 the ***Universal Lifespan Equation***, whose calculations, based on $\boldsymbol{a}$, $\boldsymbol{b}$, and $\boldsymbol{c}$ values from growth data, distinguished between the longest lifespans (humans and cows), and the shortest (nematodes), lumping in between animals with 1-to-20-year lifespans (FIGURE 3 in text, FIGURE A22). For the ten species we have examined, the average $\boldsymbol{Z}$ value of the average lifespan for each species, $\boldsymbol{Z_A}\approx 0.000037$. The average $\boldsymbol{Z}$ value of the longest known lifespans, $\boldsymbol{Z_L}\approx 0.000024$.

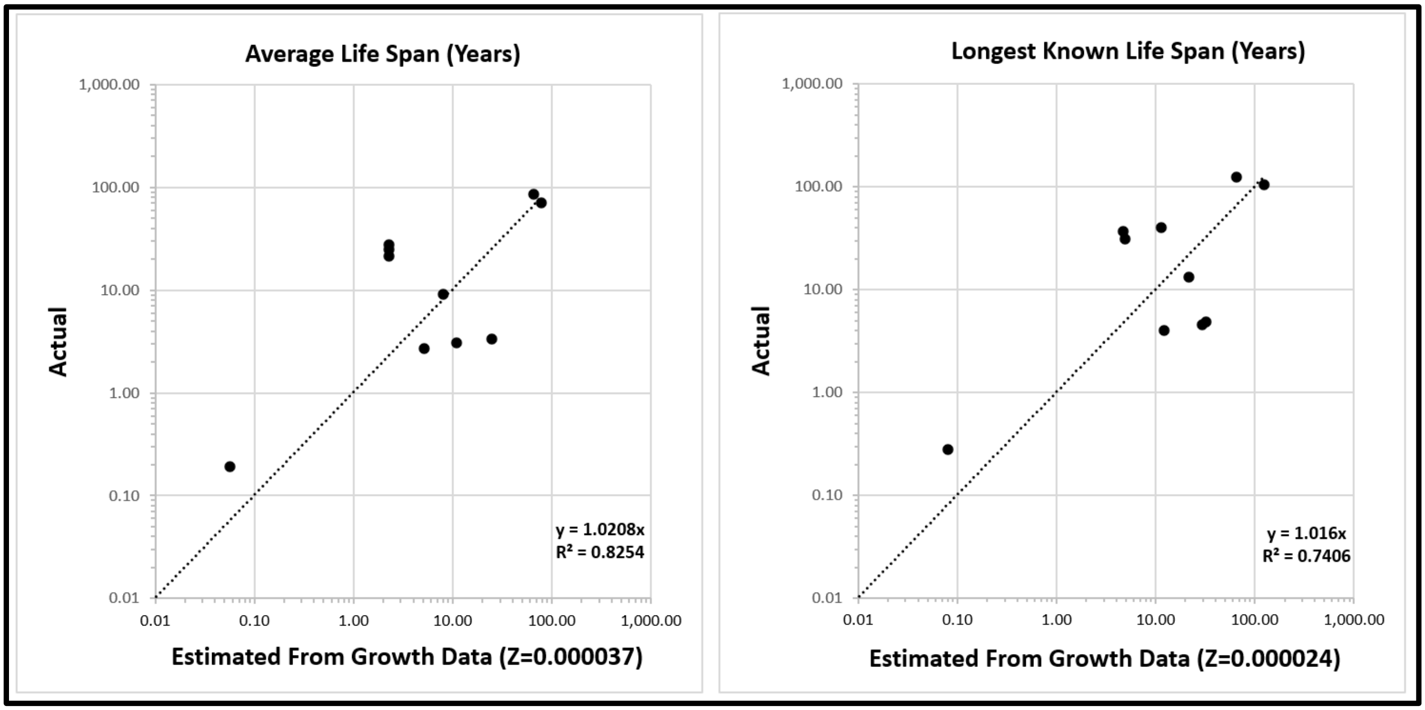


**<u>FIGURE A22</u>: Life Span - Actual vs Predicted from Growth Data** with the ***Universal Lifespan Equation*** (#6). Datapoints are for Humans, Frogs, Nematodes, Chickens, Cows, Geese, Mice, Quail, Rats, Turkeys.

##### The Pace and Shape of Lethality

While $\boldsymbol{Z}$ provides a single measure of lifespan, death occurs continuously, frequently increasing in magnitude, sometimes exponentially, as noted by Gompertz in 1825[114]. The ***Universal Mitotic Fraction in Time Equation*** also displays an accelerating decline in ***Mitotic Fraction***, although of different form than Gompertz's. Of course, mortality probably doesn't directly reflect the decrease in the ***Mitotic Fraction***, but the increase in the times since last mitoses, which can be calculated with ***Cellular Population Tree Visualization Simulation*** (Reference).

Species have lifespans that are short and long, whose risk of death may increase exponentially with age, as Gompertz showed for humans, or in other ways, or not at all, as Vaupel and colleagues made clear in the taxonomically wide survey of aging.[115]This variety has been described as the pace and shape of lethality, appearing as the "two orthogonal axes of life history".[116] Note that for the 10 species we have examined, the curves of the ***Universal Mitotic Fraction in Time Equation*** (#5) cross over each other (FIGURE A21). Such interweaving is ascribable to the capacity of the parameters $\boldsymbol{a}$ and $\boldsymbol{b}$ to twist these curves and the capacity of the parameter $\boldsymbol{c}$ to stretch or contract them. This suggests a useful place to begin to consider the possible basis for the quantitative pathways of aging and their consequences.

#### The Gompertzian Force of Mortality

Not all creatures in a species live to the same age. In 1835, Gompetz reported that the chance of death for humans, $\boldsymbol{D}$, increases exponentially with age, $\boldsymbol{t}$ (FIGURE A29):

$$D = G_H \cdot e^{G_s \cdot t} \qquad (7)$$

which is equivalent to:

$$log(D) = (G_s \cdot t) + \log(G_H) \qquad (8)$$

This expression is called the ***Gompertz Mortality Equation***, and the exponential increase in the chance of death that occurs with age that it captures is called the ***Gompertzian Force of Mortality***. One can visualize these concepts by comparing the chance of death, $\boldsymbol{D}$, against age, $\boldsymbol{t}$, on a log graph, a ***Gompertz Plot***, yielding a straight line, a ***Gompertz Line***, whose slope, $\boldsymbol{G_s}$, we call the ***Gompertzian Slope***, and whose height is seen in $\boldsymbol{G_H}$, the ***Gompertzian Height***. ***Gompertz Plots*** for humans and mice can be seen in FIGURE A29. This same log-linear ***Gompertz Line*** has also been found to capture the chance of death after exposure to a variety of pathogens, including COVID-19 (FIGURE A29). In the accompanying manuscript, we shall outline how this fit to the ***Gompertz Mortality Equation*** allows one to tease out the impact of vaccination, and other considerations, on COVID-19 lethality and transmission, and the illnesses of old age generally.[117]

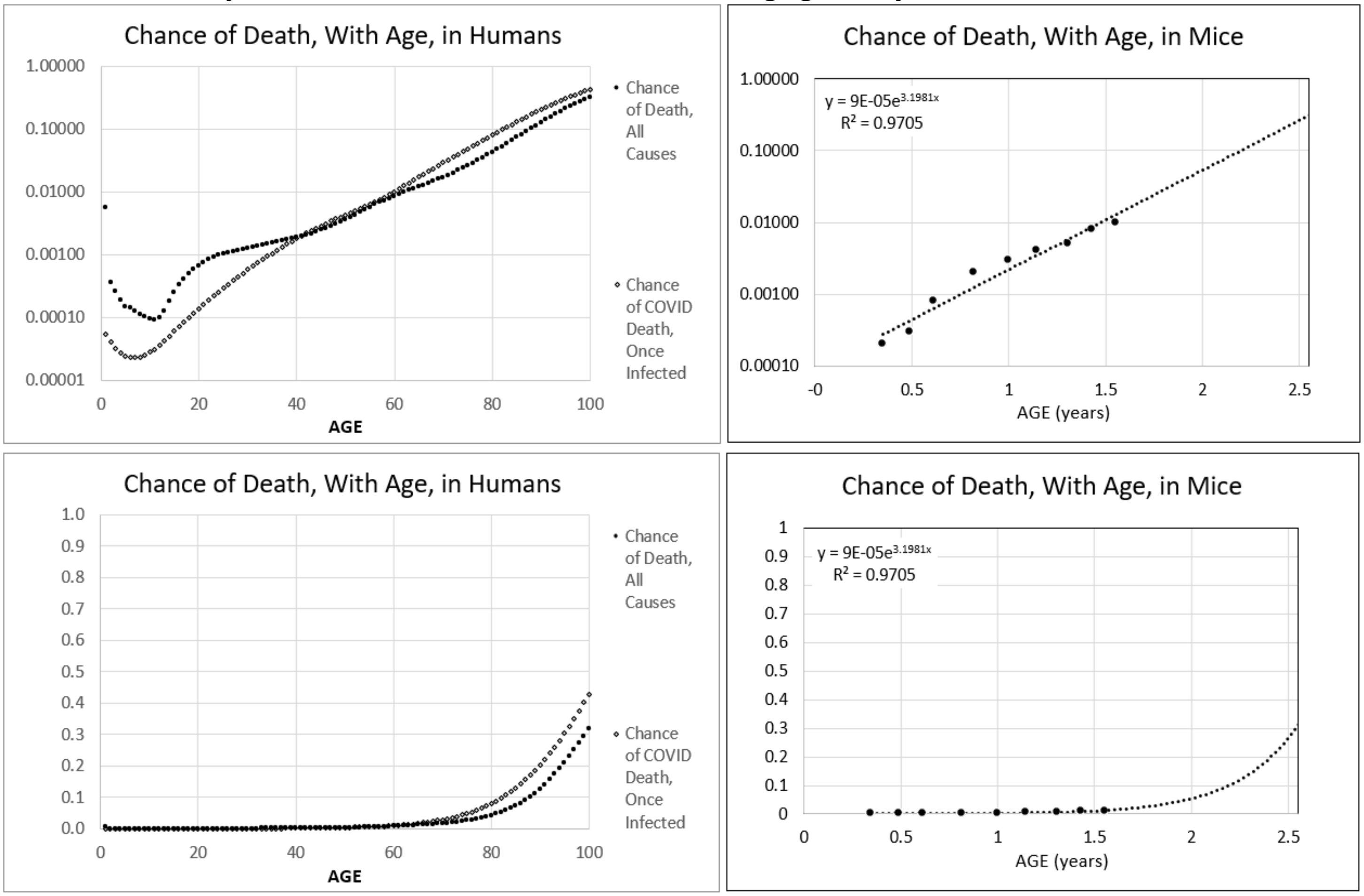


[2022 Variation in the COVID-19 infection–fatality ratio and fatality mice and human v3 jsm 8 17 22 .xlsx]VARIATION'!$AO$33

**<u>FIGURE A29</u>**

#### Growth, Decay, and Aging

As we have seen above, the examination of body size in units of numbers of cells, ***N***, reveals that the rate of animal growth declines with age by the reduction in the fraction of cells that are dividing, by the ***Mitotic Fraction in Time Equation***. This decline begins gradually early in life, gains speed early in development, and then settles to a more regular rate as one enters adulthood. For example, the ***Mitotic Fraction in Time Equation*** tell us that after the age 50, the decline in the ***Mitotic Fraction***, ***m***, appears like a slightly bent straight line (FIGURE A30), that is:

$$m \approx m_0 - (m_d) \cdot t \qquad (9)$$

Where $m_0$ is the ***Mitotic Fraction*** projected back to the time of conception (***t*=0**, not biologically relevant, because the linearity in Equation #9 does not apply to early life, but mathematically convenient for studying old age), and $m_d$ is the approximate rate of decline in the ***Mitotic Fraction*** in old age.

Consider that once a cell has divided, it will remain functional for ***F*** days, unless it divides again. It follows that a part of the body with $N_p$ cells will contain $N_{pf}$ functional cells:

$$N_{pf} = N_p \cdot F \cdot m \qquad (10)$$

Combining Equations #9 and #10:

$$N_{pf} \approx (m_0 - (m_d \cdot t)) \cdot N_p \cdot m \qquad (11)$$

Body parts have specialized cells, doing different things. For example: in the pancreas, some cells make insulin, while others make digestive enzymes; in the heart some cells are muscle, others gristle; in the liver, some cells make blood proteins, others modify bile; etc.; etc. The immune system embraces this specialization with enthusiasm, with lymphocytes of many specialized types, starting from T-cells and B-cells, then subtypes, then yet more specialized cells as the results of massive cut-and-paste randomization of antibody and T-cell receptor genes, and then, somatic mutation, just to name a few. Generally, each cell of a part of the body has a probability, ***p***, of being able to do a certain task, where:

***p*** = (number of specialized cells in the body part) / (number of all cells in the body part) (12)

Let us consider the special case of $p_a$ as the chance of any single lymphocyte making an antibody to the COVID-19 virus spike protein. The body can make that antibody as long as it has at least one such functional lymphocyte with its genes arrayed to make that antibody. However, the ***Mitotic Fraction***, ***m***, declines as we age, as seen in equation #9, the number of capable lymphocytes also declines, as seen in equation #11. Indeed, the chance of the body NOT having even one such capable lymphocytes, $D_L$, is:

$$D_L = (1 - p_a)^{N_{pf}} \qquad (13)$$

By plugging in Equation #e, which captures the approximately linear decline in ***Mitotic Fraction***, ***m***, that occurs in late adult life, we arrive at:

$$D_L = (1 - p_a)\text{^}[(m_0 - m_d * t) * N_p * F] \qquad (14a)$$

$$D_L = (1 - p_a)\text{^}(m_0 * N_p * F - m_d * t * N_p * F) \qquad (14b)$$

$$D_L = (1 - p_a)\text{^}(m_0 * N_p * F) * (1 - p_a)\text{^}(- m_d * N_p * F * t) \qquad (14c)$$

$$D_L = (1 - p_a)\text{^}(m_0 * N_p * F) * e\text{^}(- \log(1 - p_a)\, m_d * N_p * F * t) \qquad (14d)$$

$$D_L = G_{HL} \cdot e^{G_{SL}} \qquad (15)$$

Which, remarkably, is the ***Gompertz Mortality Equation***, with:

$$G_{HL} = (1 - p_a)^{m_0 \cdot N_p \cdot F} \qquad (16)$$

and:

$$G_{SL} = -m_d \cdot N_p \cdot F \cdot \log(1 - p_a) \qquad (17)$$

Thus, ***Cellular Phylodynamics***, the analysis of growth in units of numbers of cells, ***N***, has allowed us to see that aging's exponential ***Gompertzian Force of Mortality*** is the natural, and direct, result of the deceleration of growth that marks animal's approach to adult size, as captured by the ***Universal Growth Equation***.

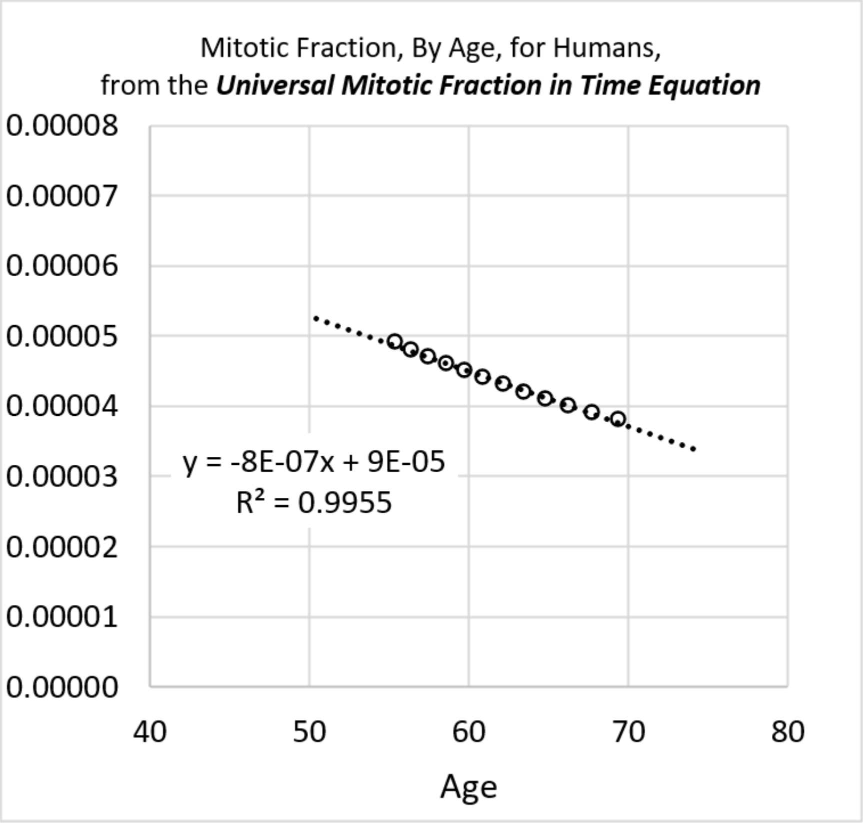
Mitotic Fraction, By Age, for Humans,
from the *Universal Mitotic Fraction in Time Equation*
0.00008
0.00007
0.00006
0.00005
0.00004
0.00003
0.00002
0.00001
0.00000
40
50
60
70
80
Age
y = -8E-07x + 9E-05
R² = 0.9955

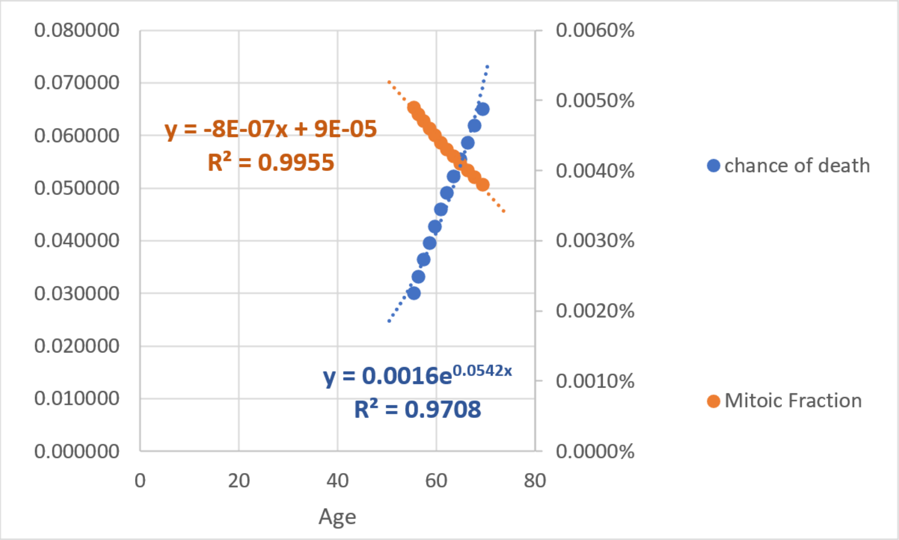
0.080000
0.070000
0.060000
0.050000
0.040000
0.030000
0.020000
0.010000
0.000000
0
20
40
60
80
Age
0.0060%
0.0050%
0.0040%
0.0030%
0.0020%
0.0010%
0.0000%
y = -8E-07x + 9E-05
R² = 0.9955
y = 0.0016e0.0542x
R² = 0.9708
chance of death
Mitoic Fraction

**FIGURE A30**

Bear in mind that we have used lymphocytes making anti-COVID-19 antibodies to illustrate how the ***Gompertzian Force of Mortality*** will arise as a result of the decline in growth. However, this finding applies to lethality arising by failure of any type of specialized cell, in any of the body's organs and tissues.

It may seem a bit surprising that a linear reduction in the fraction of functional cells that occurs in the ***Death Zone*** will lead to an exponential decline in the chance of having at least one such functional cell. However, this is a curious consequence of the subtle fact that cells are discrete things. We can picture what's going in by considering a game played with dice, in which a player is allowed to continue, as long as at least one ***Snake Eye*** appears. However, with each sequential throw, one of the dice will disappear. Let's imagine that the player begins with a ten dice, and thus the odds of continuing are rather good at 1-(5/6)^10 = 83%. At nine dice, the odds are slightly less favorable, with a 1-(5/6)^9 = 80% chance of continuing. With four dice, the odds have dropped to roughly 52%; three dice, the odds are 42.1%; with two dice, it becomes 30.6%; and with one die, there is only a 16.7% chance. As we can see, while the number of dice goes down linearly with time, the chance of having at least one ***Snake Eye*** goes down exponentially (FIGURE A31). In a similar fashion, the chance of having at least one lymphocyte with the capacity to keep a person alive after being exposed to a potentially lethal agent such as COVID also declines exponentially as we age. Thus, we have seen in general terms that the roughly linear decline in the ***Mitotic Fraction***, ***m***, by the ***Universal Mitotic Fraction Equation***, can lead to an exponential decline in survival to COVID, and survival generally, as captured by the ***Gompertz Mortality Equation***. A much more rigorous examination of this process, yielding qualitatively the same conclusions noted above, but with greater rigor, can be seen in the APPENDIX and will be published later.

Finally, let us note that $N_p$ is the number of cells in a body part from which specialized cells may arise. The specific example noted above examined the task of any single lymphocyte making an antibody to the COVID virus spike protein. As we shall show in the accompanying paper, immunization appears to decrease the chance of death after COVID infection through lowering the ***Gompertzian Height***, $G_{HL}$. Equation #g shows that this may occur by increasing the number of functional cells that might make such an antibody in an unimmunized individual, or $N_{pf}$. Specifically, equation #h shows that the ***Gompertzian Height***, $G_{HL}$, may be lowered by increasing $m_0$, or the mitotic fraction projected back to the time of conception. However, immunization appears to not affect the ***Gompertzian Slope***, $G_{SL}$. Equation #i shows this may occur if the approximate rate of decline in the ***Mitotic Fraction*** in old age, $m_d$, is not affected by immunization. As a result, this rate of decline in the ***Mitotic Fraction*** in old age, $m_d$, may be intrinsic.

Immunization is the process which increases the number of such cells, and vaccination is the method of immunization done before infection occurs. Since mice, like humans, display the log-linear ***Gompertzian Force of Mortality*** (FIGURE A31), and since immunology encompasses the study of the number of such cells, this give us an experimental entry into understanding how immunization may be applied to maximum effect. All one has to do is count the lymphocytes. We shall return to this opportunity below, briefly, and in greater details in the accompanying paper.

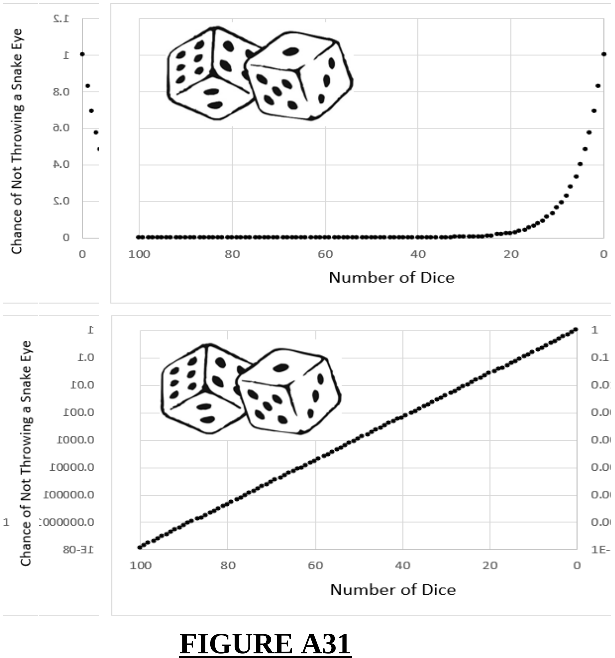


**FIGURE A31**

**BOX 1**
**TERMS, EXPRESSIONS, AND FINDINGS FOR**
**THE *CELLULAR PHYLODYNAMICS* OF ANIMAL GROWTH**

***Cellular Phylodynamics***:
The analysis of growth and development in units of integer numbers of cells, $\boldsymbol{N}$. $\boldsymbol{N_w}$ is the number of cells in an animal as a whole while $\boldsymbol{N_p}$ is the number of cells in a part of the body, such as a ***cell lineage***, ***tissue***, ***organ***, ***or anatomical structure***.

The ***Mitotic Fraction, m***:
A measure of the fraction of cells dividing, derived from growth data.

***Mitotic Fraction Method***
The ***Cellular Phylodynamic Analysis*** method, Equation #3b, which gives a measure how many cells would have to divideto account for the amount of growth that occurs over each period of time, the ***mitotic fraction***.

The ***Universal Mitotic Fraction Equation***:
An empirically derived expression, $\boldsymbol{m = a^{(N_w^b)}}$, which captures a close approximation of the decline in the ***mitotic fraction***, ***m***, that occurs as animals increase in size, $\boldsymbol{N_w}$, potentially traceable to the discrete allocation of growth factor molecules among cells.

The ***Universal Growth Equation***
An empirically derived expression for the rate of animal growth, $\boldsymbol{\frac{dN_w}{dt} = \left(\frac{\log 2}{c}\right) N_w a^{(N_w^b)}}$. ***c*** is the ***cell cycle time***, the amount of time it takes cells to divide, traceable to how much DNA the genome contains.

The Integrated Form of the ***Universal Growth Equation***
An empirically derived expression that captures a close approximation of how animals increase in size, $\boldsymbol{N_w}$, with time, ***t***, such that $\boldsymbol{N_w = \int \left(\frac{\log 2}{c}\right) N_w a^{(N_w^b)}}$ [Closed form: $\boldsymbol{t = c \cdot \left(\mathrm{Ei}\left[-N_w^b \cdot \log(a)\right] - \mathrm{Ei}[-\log(a)]\right) \frac{1}{b} \log(2)}$].

***UNI-GROWTH***.
The biological process captured by the ***Universal Mitotic Fraction Equation*** and the ***Universal Growth Equation***.

***Cellular Allometric Growth Analysis***.
The ***Cellular Phylodynamic Analysis*** of relative growth carried out by examining paired $\boldsymbol{N_p}$ / $\boldsymbol{N_w}$ values on log-log graphs. When carried out a full lineage chart for an animal, which characterizes each cell from the first fertilized egg onward, this is called ***Cellular Allometric Lineage Growth Analysis***. When carried out without lineage data, but simply with cell numbers, this is called ***Cellular Allometric Batch Growth Analysis***.

***Cellular Allometric Lineage Growth Analysis***.
The ***Cellular Phylodynamic Analysis*** of cell lineages, carried out by counting the number of cells in each ***lineage***, $\boldsymbol{N_p}$, from the first ***Founder Cell*** onward ($\boldsymbol{N_p} = 1$),

***Cellular Allometric Batch Growth Analysis***.
The ***Cellular Phylodynamic Analysis*** of ***tissues***, ***organs***, and ***anatomical structures*** based on aggregate size, which is then converted into cell number, relying on the observation that there are about $10^8$ cells in every gram of tissue[57], or by other such batch methods, such as by making cell counts microscopically, or measuring DNA content.

The ***Cellular Allometric Growth Equation***:
An empirically derived expression that captures the relationship between the number of cells in an individual part of the body, $\boldsymbol{N_p}$, with the number with the number of cells in the embryo as a whole, $\boldsymbol{N_w}$, such that $\log(N_w) = \left(\frac{1}{S_N}\right)\log(N_p) + \log(B_N)$.

The ***Cellular Allometric Birth***, $\boldsymbol{B_{N1}}$, of the ***Cellular Allometric Growth Equation***:
An empirically derived measure of the number of cells in the body as a whole, $\boldsymbol{N_w}$, at the point where the ***cellular allometric growth equation*** indicates that a body part could have been a single cell ($\boldsymbol{N_p} \approx 1$). For a cell lineage, $\boldsymbol{B_{N1}}$ is the number of cells in the body as a whole, $\boldsymbol{N_w}$, when the cell lineage is exactly 1 single cell ($\boldsymbol{N_p} = 1$).

***Cellular Allometric Slope***, $\boldsymbol{S_N}$, of the ***Cellular Allometric Growth Equation***:
An empirically derived measure of the relative growth of each part of the body, whose value can be traced to subtle ***cell-heritable*** changes in the ***cell cycle time***, potentially caused by processes such as DNA methylation.

***ALLO-GROWTH***
The biological process captured by the ***Cellular Allometric Growth Equation***.

***Cellular Population Tree Visualization Simulation***
The creation of likely approximations of cell lineage charts with the parameters of the ***Universal Growth*** and ***Cellular Allometric Growth Equations***, derived from macroscopic growth data.

#### FIGURE LEGENDS

**FIGURE A1**: **Growth, in units of numbers of cells, $N_w$, from fertilization until maturity, in units of days, *t*.** Datapoints for animal size's ($N_x$) versus time (***t***), for ***C elegans***, chickens, mice, turkeys, quail, geese, frogs, and humans, and their fit to the numerically integrated form of the ***Universal Growth Equation*** (#5b**).**

**FIGURE A2**: **Fit of human growth to the *Universal Growth Equation* (red), the *logistic equation* (blue), and the *Gompertz equation* (green).** Data values as black circles.

**FIGURE A3: Analysis of the closed form integration of the *Universal Growth Equation* (#3).**

**FIGURE A4**: **Decline in fraction of cells dividing, the *Mitotic Fraction, m*, calculated by the *Mitotic Fraction Method*, that occurs as animals increase in size, as seen by the *Universal Mitotic Fraction Equation*.** Data points for size, $N_w$, in integer units of numbers of cells, vs. the ***Mitotic Fraction***, ***m***, from fertilization, until maturity, for humans.

**FIGURE A5**: **Values of the *Mitotic Fraction*, *m*, calculated by the *Mitotic Fraction Method*, as a function of animal size, $N_w$, as shown at various scales.** Data points for animal size, $N_w$, in integer units of numbers of cells, vs. the ***Mitotic Fraction***, ***m***, from fertilization, until maturity, for humans, chickens, ***C elegans*** nematode worms, and mice.

**FIGURE A6**: **Comparison of the fit of *Mitotic Fraction, m,* as a function of size, $N_w$, with human growth data, to the *Universal Mitotic Fraction Equation* (green) and *West equation* (red).**

**FIGURE A7**: **Wave-like deviations ("*ripples*") from the *Universal Growth Equation curve*, for human growth.**

**FIGURE A8**: ***Cell lineage chart* of the *cell lineages* of the developing root knot nematode, *Meloidogyne incognita*.** Shown are the conventional and ***BinaryCellName Lineage*** naming methods. The **AB *lineage*** of nematodes forms the skin while the **E *lineage*** forms the intestine. ***Cell lineage chart*** and data from Calderón-Urrea et al[21]

**FIGURE A9**: ***Cellular Lineage Growth Analysis* of the *cell lineages* of the developing root knot nematode, *Meloidogyne incognita*.**[21]

**FIGURE A10**: ***Cellular Lineage Growth Analysis* of the *cell lineages* of the developing *C elegans* nematode**

**FIGURE A11**: ***Cellular Lineage Growth Analysis* of the *cell lineages* of the chordate tunicate *Oikopleura dioica*.** Unlike ***C elegans*** and ***M incognita***, all of the cases of ***Cellular Selection*** for this species reflect reductions in cell number, although this could be due to the limited size of the dataset rather than a biological difference. ***Cell lineage chart*** data from Stach et al[88]

**FIGURE A12**: ***Cellular Allometric Batch Growth Analysis* of *tissues*, *organs*,** and ***anatomical structures* of the chick embryo.**
Growth of the parts of the chick embryonic gizzard, liver, heart, and kidney, in units of numbers of cells, in comparison to the number of cells in the embryo as a whole shown on log-log graphs, showing how the fraction of cells comprising in the gizzard and liver increases as the number of cells in the embryo increase with development and how the fraction of cells comprising in the heart decreases as the number of cells in the embryo increase with development, and how the fraction of cells comprising in the kidney remains roughly constant as the number of cells in the embryo increases with development,

**<u>FIGURE A13</u>: *Cellular Allometric Batch Growth Analysis* of tissues, organs, and anatomical structures of the rat embryo**.
Growth of body parts of the rat embryonic liver, brain, kidney and foreleg, in units of numbers of cells, in comparison to the number of cells in the embryo as a whole, are shown on log-log graphs. Note how the fraction of cells comprising in the liver kidney and forleag increases as the number of cells in the embryo increase with development, while the fraction of cells comprising the brain decreases.

**<u>FIGURE A14</u>: *Cellular Allometric Batch Growth Analysis* of tissues, organs, and anatomical structures of the mouse embryo**.
Growth of body parts of the mouse embryonic liver, brain, kidney and foreleg, in units of numbers of cells, $N_p$, in comparison to the number of cells in the embryo as a whole, $N_w$, shown on log-log graphs. Note how the fraction of cells comprising in the liver kidney and foreleg increases as the number of cells in the embryo increase with development, while the fraction of cells comprising the brain decreases.

**<u>FIGURE A15</u>: *Cellular Allometric Batch Growth Analysis* of tissues, organs, and anatomical structures of the developing human fetuses, from autopsy data, of individual fetuses**.
For additional human relative growth data, see FIGURE A16

**<u>FIGURE A16</u>: *Cellular Allometric Batch Growth Analysis* of tissues, organs, and anatomical structures of the developing human fetuses, from autopsy data, averaged values.**[118]
For additional human relative growth data, see FIGURE A15

**<u>FIGURE A17</u>: *Cellular Allometric Batch Growth Analysis* of the Zebrafish Lens.<u>FIGURE A18</u>: The *Cellular Allometric Growth Equation*, $\log(N_w) = (1/S_N)\log(N_p) + \log(B_{N1})$.**

**<u>FIGURE A19</u>: Calculations for an idealized part of the body that is made from 3 *Founder Cells*, arising early in development**

**<u>FIGURE A20</u>: The *Universal Mitotic Fraction in Time Equation*** (#5) **captures change in *Mitotic Fraction*, *m*, with *Age*, *t*.**

$$t(m) = \frac{c}{b \cdot log(2)} \cdot \quad [\mathrm{Ei}(-log(m)) - \mathrm{Ei}(-log(a))] \qquad (4)$$

The graph on the right side shows the bottom 0.2% of the Y-axis of the graph on the left side: the ***Death Zone*** occurs when fewer the 0.03% of the cells of the body are dividing. Each curve ends at the age of the oldest known member of its species.

**<u>FIGURE A21</u>: The inverse of the *Universal Mitotic Fraction in Time Equation*** (#5) **visualizes the change in the fraction of cell not dividing with age.**

**<u>FIGURE A22</u>: Life Span - Actual vs Predicted from Growth Data** with the ***Universal Lifespan Equation*** (#6). Datapoints are for Humans, Frogs, Nematodes, Chickens, Cows, Geese, Mice, Quail, Rats, Turkeys..

#### TABLE A1
#### FILES WITH GROWTH DATA, FOR DISTRIBUTION TO INTERESTED READERS

| Species | Genus Species | Basic Data Files | Calculations Files |
|---|---|---|---|
| Human Males | *Homo sapiens* | Boys Basic Growth Data jsm 2 12 2013.xlsx | 140330 Human males.xlsx |
| Frogs | *Rana pipiens* | Rana Basic Growth Data jsm 2 15b 2013.xlsx | 140330 Rana Pipiens.xlsx |
| Nematodes | *C. elegans* | BASIC FORM Celegans SingleSheet jsm 2 13 13 LogMethod.xlsx | 140426 C elegans.xlsx |
| Chickens | *Gallus gallus* | CHikcens Aggrey and Byerley and cleavage Data 2 14 2013.xlsx | 140426 Chickens.xlsx |
| Cows | *Bos taurus* | COW Basic Growth Data jsm 1 27 2013.xlsx | 140426 Cows.xlsx |
| Geese | *Anser anser* | BASIC FORM Geese SingleSheet jsm 2 14 2013 Log Method.xlsx | 140426 Geese.xlsx |
| Mice | *Mus musculus* | BASIC FORM MicePoiley M AKR jsm 1 28 2013 Log&LinearMethods.xlsx | 140426 Mice.xlsx |
| Quail | *Colinus virgianus* | BASIC FORM Quail SingleSheet jsm 2 16 2013.xlsx | 140426 Quail.xlsx |
| Rats | *Rattus norvegicus* | RAT Basic Growth Data jsm 2 14 2013.xlsx | 140426 Rats.xlsx |
| Turkey | *Meleagris gallopavo* | BASIC FORM TURKEYwithOvaposition SingleSheet jsm 2 8 2013.xlsx | 140426 Turkey.xlsx |
| Clams | *Merceneria mercenaria* | BASIC FORM LOG METHOD mercenaria 1 11 2013 v2 from Boys SingleSheet jsm 2 12 2013 | 140705 Mercenaria.xlsx |
| LOCATION: | | C:_0\My Papers\Growth_DRAFTS__final Excell Files_Basic Data 7 7 14 | C:_0\My Papers\Growth_DRAFTS__final Excell Files_Phils Analysis 7 7 14 |

#### TABLE A2
##### *Universal Growth Equation*.
Parameters and goodness-of-fit metrics

| Organism | Genus species | *Fit of the data to the rate of growth* $g=N*a^N^b$ ($c$=*Cell Cycle Time*, in days) | | | | | *Fit of the data to growth* $N=\int[\log(2)/c]*N*a^N^b$ |
|---|---|---|---|---|---|---|---|
| | | c | a | b | R^2 (linear fit) | R^2 (log fit) | R^2 (log fit) |
| Humans (♂) | *Homo sapiens* | 1.016 | 0.943 | 0.169 | 89.9% | 95.0% | 99.5% |
| Frogs | *Rana pipiens* | 0.070 | 0.925 | 0.269 | 57.0% | 82.3% | 97.3% |
| Nematodes | *C. elegans* | 0.011 | 0.946 | 0.680 | 66.7% | 91.6% | 99.8% |
| Chickens | *Gallus gallus* | 0.127 | 0.906 | 0.166 | 80.8% | 97.8% | 99.8% |
| Cows | *Bos taurus* | 0.921 | 0.882 | 0.128 | 76.8% | 95.1% | 100.0% |
| Geese | *Anser anser* | 0.048 | 0.898 | 0.170 | 77.0% | 92.9% | 97.1% |
| Mice | *Mus musculus* | 0.508 | 0.990 | 0.281 | 65.0% | 96.2% | 99.7% |
| Quail | *Colinus virgianus* | 0.292 | 0.795 | 0.126 | 76.5% | 95.3% | 94.2% |
| Rats | *Rattus norvegicus* | 0.417 | 0.950 | 0.197 | 66.5% | 90.6% | 99.5% |
| Clams | *M. mercenaria* | 0.033 | 0.962 | 0.275 | 79% | 91% | 99.5% |
| Turkeys | *Meleagris gallopavo* | 0.028 | 0.757 | 0.122 | 65.3% | 92.2% | 99.7% |

Zebrafish (*Danio rerio*) and European sea bass (*Dicentrarchus labrax*) also show these features of growth (data not shown)
